# Prognostic factors for a change in eye health or vision: A rapid review

**DOI:** 10.1101/2024.01.18.24301468

**Authors:** Greg M. Hammond, Antonia Needham-Taylor, Nathan Bromham, Elizabeth Gillen, Lydia Searchfield, Ruth Lewis, Alison Cooper, Adrian Edwards, Rhiannon Tudor Edwards, Jacob Davies

## Abstract

The general public are advised to have regular routine eye examinations to check their vision and ocular health; however current UK guidance on how often to have eye examinations is not evidence-based and was issued in 2002.This Rapid Review aims to provide an evidence base that stakeholders can use to form updated guidance for Wales by asking the question ‘What are the prognostic factors for a change in ocular status in the general population attending routine eye examinations?’

The review included evidence available from January 2009 up until August 2023. Evidence was included from 2011 up until 2023. 19 studies were included: two systematic reviews; nine prospective cohort studies; three retrospective cohort studies; two longitudinal studies; two case-control studies; and one cross-sectional study were included.

**Research Implications and Evidence Gaps:** Future research to inform appropriate eye examination intervals should be narrower in focus to ensure as much relevant and useful evidence as possible is gathered. There are large amounts of evidence on prevalence and prognostic factors for prevalent conditions, which did not meet the inclusion criteria of this rapid review which looks at incident or changing conditions.

**Policy and Practice Implications:** Caution should be taken if using this review for decision making on appropriate eye examination intervals due to low certainty and generalisability. This review should be used to identify key prognostic factors and suggesting these for further targeted research and evidence synthesis.

**Economic considerations:** Sight loss costs the UK economy 25 billion pounds per annum, with more than 2 million people in the UK currently living with sight loss. The economic implications of appropriate or inappropriate testing intervals for different causes of vision loss will be different. When captured at a population wide scale, the earlier detection of conditions through examination can result in significant economic savings.

**Funding statement:** The Centre for Health Economics and Medicines Evaluation, the Bangor Institute for Medical and Health Research, and the Swansea Centre for Health Economics were funded for this work by the Health and Care Research Wales Evidence Centre, itself funded by Health and Care Research Wales on behalf of Welsh Government.

## EXECUTIVE SUMMARY

### What is a Rapid Review?

Our rapid reviews (RR) use a variation of the systematic review approach, abbreviating or omitting some components to generate the evidence to inform stakeholders promptly whilst maintaining attention to bias.

### Who is this summary for?

This Rapid Review is intended for use by clinical leaders and decision makers in Wales’ primary eye care services. The evidence in this Review is intended to be used to examine the risk of a person experiencing a change in their ocular health, vision, or systemic health that affects their eyes so that guidance can be produced on how often people should attend for routine eye examinations based on their individual risk factors.

It is also intended to identify gaps in the evidence to determine where further research is required for certain risk factors or patient groups.

### Background / Aim of Rapid Review

The general public are advised to have regular routine eye examinations to check their vision and ocular health; however current UK guidance on how often to have eye examinations is not evidence-based and was issued in 2002.

This Rapid Review aims to provide an evidence base that stakeholders can use to form updated guidance for Wales by asking the question “What are the prognostic factors for a change in ocular status in the general population attending routine eye examinations?”

### Results

#### Recency of the evidence base

- The review included evidence available from January 2009 up until August 2023. Evidence was included from 2011 up until 2023.

#### Extent of the evidence base

- 19 studies were included: two systematic reviews; nine prospective cohort studies; three retrospective cohort studies; two longitudinal studies; two case-control studies; and one cross-sectional study were included.

#### Key findings and certainty of the evidence

- Demographic prognostic factors: age, sex, ethnicity, and household net worth are potential prognostic factors for a change in ocular health or vision.
- Ocular prognostic factors: intraocular pressure, family history of glaucoma, visual acuity, visual field mean deviation, spherical equivalent refraction, high myopia, age-related macular degeneration, glaucoma, and cataract are potential prognostic factors for a change in ocular health or vision.
- Lifestyle/behaviour prognostic factors: diet, alcohol intake, smoking, time spent outdoors, and time spent reading are potential prognostic factors for a change in ocular health or vision.
- Systemic health prognostic factors: hypertension, heart disease, cholesterol, diabetes, peripheral arterial disease, hypercoagulable state, stroke, pregnancy, age at menarche, oral contraceptive use, and atopy are potential prognostic factors for a change in ocular health or vision.
- Increasing length of time between eye examinations is a potential prognostic factor for a change in ocular health or vision.
- The level of certainty for all prognostic factors is low as there was generally only one study reporting for each individual outcome.
- Studies were often performed in specific populations, meaning the results cannot be applied to the general population, particularly due to low study numbers per outcome.

### Research Implications and Evidence Gaps

- Future research to inform appropriate eye examination intervals should be narrower in focus to ensure as much relevant and useful evidence as possible is gathered. Prognostic factors or specific ocular conditions of interest potentially need to be investigated individually for their effect on a change in ocular status.
- There are large amounts of evidence on prevalence and prognostic factors for prevalent conditions, which did not meet the inclusion criteria of this rapid review which looks at incident or changing conditions. Further evidence generation could be conducted in this area.
- Very little evidence was identified in a UK setting, more primary evidence generation may be required.
- There is a notable lack of evidence in younger adults aged under 40 years.

### Policy and Practice Implications

- Caution should be taken if using this review for decision making on appropriate eye examination intervals due to low certainty and generalisability.
- This review should be used to identify key prognostic factors and suggesting these for further targeted research and evidence synthesis.

### Economic considerations

- Sight loss costs the UK economy £25 billion per annum, with more than 2 million people in the UK currently living with sight loss.
- The economic implications of appropriate or inappropriate testing intervals for different causes of vision loss will be different.
- A new case of age-related macular degeneration (AMD) in an adult aged 50 or over, costs the UK economy £73,350 over the person’s lifetime. Lifetime costs to the UK economy for a person diagnosed with glaucoma are approximately £49,800 per person. Reducing the prevalence of these conditions by just 14 or 20 cases respectively could save the UK economy £1 million in lifetime costs.
- On economic grounds, early detection of AMD in eye care services and the eye care pathway may be of benefit due to the high level of prevalence and associated long term costs to the NHS as the condition causes irreversible, life limiting damage.
- When captured at a population wide scale, the earlier detection of conditions through examination can result in significant economic savings.

**Disclaimer:** The views expressed in this publication are those of the authors, not necessarily Health and Care Research Wales. The Health and Care Research Wales Evidence Centre and authors of this work declare that they have no conflict of interest.

## 1. BACKGROUND

### 1.1 Who is this review for?

This Rapid Review was conducted as part of the Health and Care Research Wales Evidence Centre Work Programme. The original question was suggested by the National Clinical Leads for Wales General Ophthalmic Services and the Optometry and Audiology Policy Branch, Welsh Government. Working with these stakeholders, the question was then amended to the one mentioned above.

This Rapid Review is intended for use by clinical leaders and decision makers in Wales’ primary eye care services. The evidence in this Review is intended to be used to examine the risk of a person experiencing a change in their ocular health, vision, or systemic health that affects their eyes so that guidance can be produced on how often people should attend for routine eye examinations based on their individual risk factors.

It is also intended to identify gaps in the evidence to determine where further research is required for certain risk factors or patient groups.

### 1.2 Background and purpose of this review

The general public are advised to attend routine eye examinations to regularly check their visual acuity, provide any necessary vision correction, and identify ocular health problems. Current guidance in the UK was issued in 2002 and is based on a consensus decision regarding minimum re-examination intervals, with no evidence base. The currently recommended minimum intervals are:

- Under 16 years in the absence of any binocular vision anomaly - 1 year
- Under 7 years with binocular vision anomaly or corrected refractive error - 6 months
- 7 years and over and under 16 with binocular vision anomaly or rapidly progressing myopia - 6 months
- 16 years and over and under 70 years - 2 years
- 70 years and over - 1 year
- 40 years and over with a family history of glaucoma or with ocular hypertension and not in a monitoring scheme - 1 year
- Diabetic patients - 1 year

With significant reform of Wales General Ophthalmic Services underway, it is pertinent to review the evidence that is available that may be able to inform recommendations on the frequency of routine eye examinations in Wales.

The evidence identified in this review will be used by stakeholders to help answer questions similar to the below:

- What is the risk of an asymptomatic person attending for a routine eye examination having experienced a change in ocular status?
- Is there evidence to suggest that this risk may vary between different groups?
- Can the evidence regarding this risk be used to inform appropriate time intervals between routine eye examinations?

## 2. RESULTS

### 2.1 Overview of the Evidence Base

Nineteen studies were identified that met the inclusion criteria of this rapid review. Two systematic reviews and 17 primary studies were included, all of which are observational studies. The study designs varied, with nine prospective cohort studies; three retrospective cohort studies; two longitudinal studies; two case-control studies; and one cross-sectional study included. Sample sizes were also very different across studies, with some having only a few hundred participants whilst others had more than 400,000. Full details of the eligibility criteria are presented in Section 5.1, Table 7. Full details of the included studies and the extracted data can be found in Section 6.2, Tables 8 and 9.

The results of this rapid review have been categorised into common prognostic factors. These factors are demographic, ocular, lifestyle/behaviour, and systemic health related. The factors are then further categorised into specific prognostic factors or similar categories of prognostic factors.

As the scope of this prognostic factor review is broad, and the review conducted with exploratory aims, essential restrictions were put in place to make sure the review remained tenable within the limits of rapid review methodology. Included studies were therefore limited to a pre-determined list of countries (Section 5.1, Table 7) that were determined by the review team to have similar demographics and eye care systems to the UK. Studies were also included only if they had presented their findings as odds ratios, risk ratios/relative risk, or hazard ratios – an approach that is in line with other prognostic factor reviews (Riley et al. 2019) – and had used multivariate analyses to determine these. The use of multivariate analyses means, where an association has been identified, the reported prognostic factors have an effect on the outcome that is independent of the other factors controlled for. Factors controlled for in each study are included in Table 8; there was considerable variation in the types of factors and the number of factors controlled for in each study. Owing to this being a rapid review, it was not feasible to convert other types of outcomes into ratios as is sometimes done in prognostic factor reviews. This is discussed in the limitations of this review.

Using the Quality in Prognostic factor Studies (QUIPS) tool, all but one of the primary evidence sources were determined to be of low or moderate risk of bias across all six domains of the tool. Ten of the 17 studies were assessed as moderate risk of bias for prognostic factor or outcome measurement, with eight of these relying on self-reporting, leading to increased risk of recall bias. Similarly, there were concerns regarding loss to follow-up or study attrition in seven studies and it was unclear whether the strategy for model building was appropriate and based on a conceptual framework or model in four studies. One study (Barsam et al. 2017) was determined to be at high risk of bias due to its case-control design and only including a small number of the cases in multivariable modelling.

Using the Risk of Bias in Systematic reviews (ROBIS) tool, both systematic reviews included in this rapid review were deemed to have either low risk of bias (Kessel et al. 2015) or an unclear risk of bias (Dinu et al. 2019). For the systematic review deemed unclear, issues were centred around the failure to address heterogeneity, and a lack of clarity on whether subgroup analyses were pre-specified. Both studies included meta-analyses and the results of these were extracted for this rapid review. None of the identified primary evidence sources were included in either of the systematic reviews.

### 2.2 Demographic prognostic factors

Results for this section are summarised in Table 1 with comprehensive details available in Section 6.2, Tables 8 and 9.

**Table 1:**
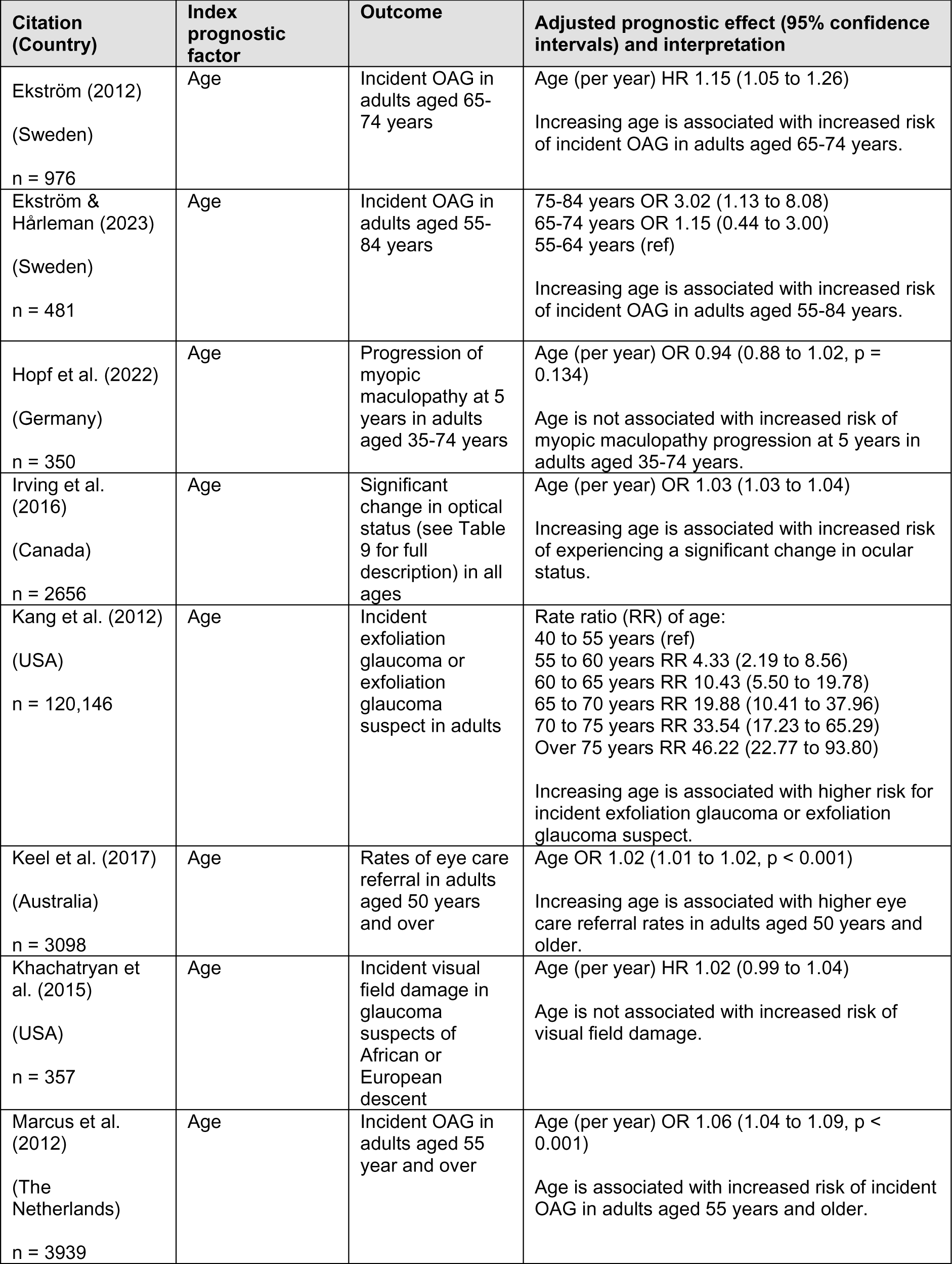
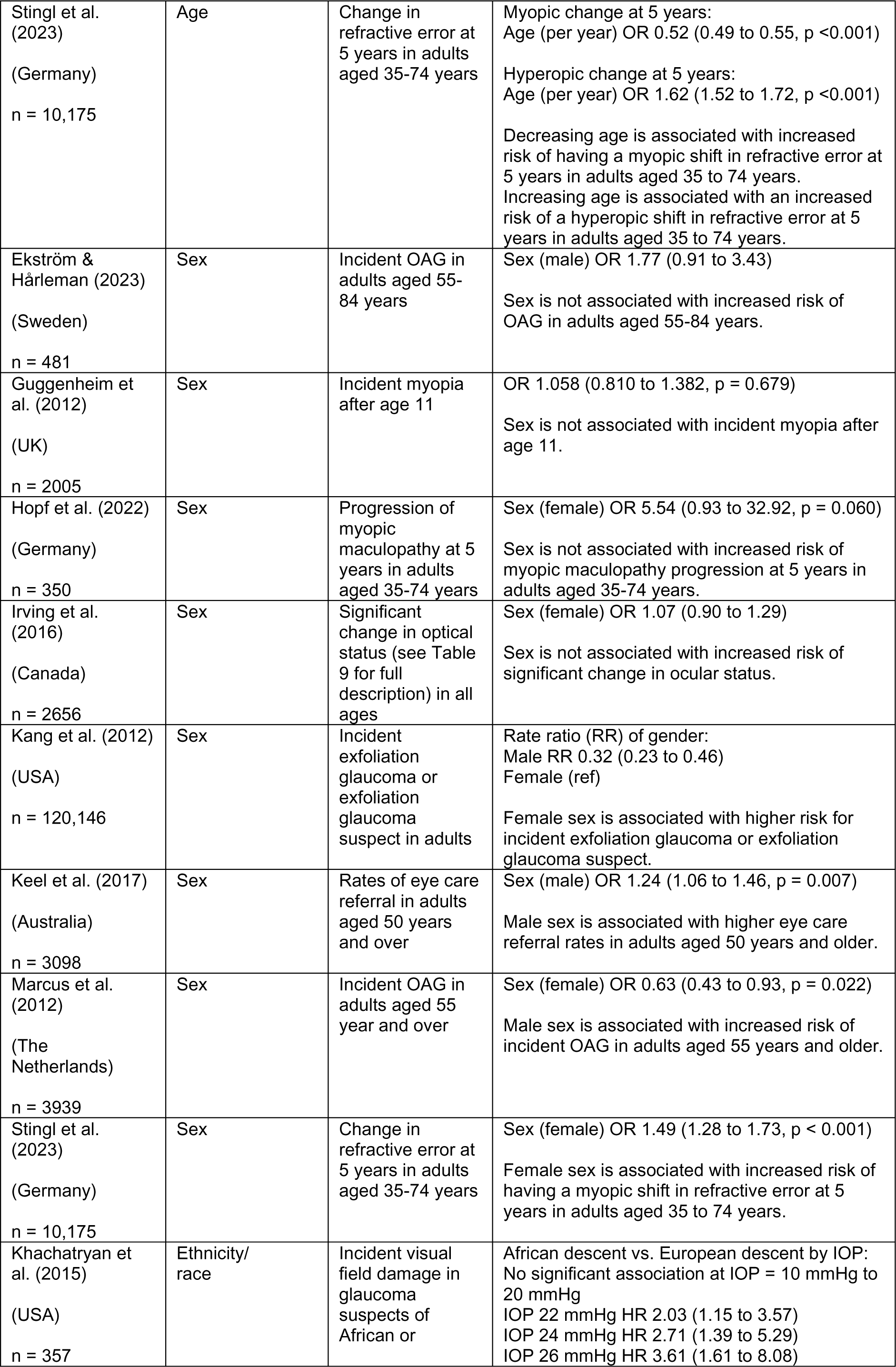
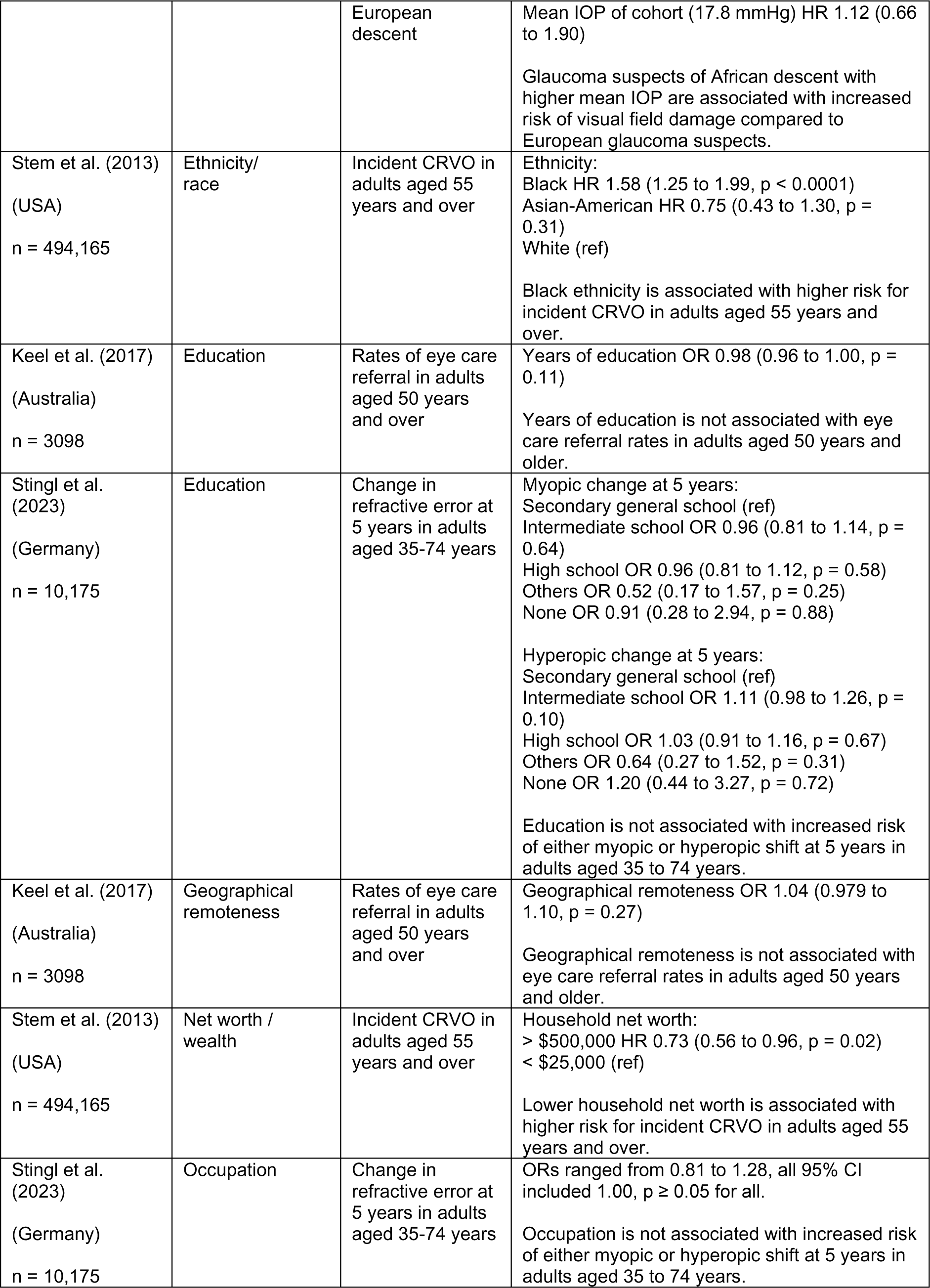
Summary of demographic prognostic factors.

| Citation (Country) | Index prognostic factor | Outcome | Adjusted prognostic effect (95% confidence intervals) and interpretation |
| --- | --- | --- | --- |
| Ekström (2012)<br>(Sweden)<br>n = 976 | Age | Incident OAG in adults aged 65-74 years | Age (per year) HR 1.15 (1.05 to 1.26)<br>Increasing age is associated with increased risk of incident OAG in adults aged 65-74 years. |
| Ekström & Hårleman (2023)<br>(Sweden)<br>n = 481 | Age | Incident OAG in adults aged 55-84 years | 75-84 years OR 3.02 (1.13 to 8.08)<br>65-74 years OR 1.15 (0.44 to 3.00)<br>55-64 years (ref)<br>Increasing age is associated with increased risk of incident OAG in adults aged 55-84 years. |
| Hopf et al. (2022)<br>(Germany)<br>n = 350 | Age | Progression of myopic maculopathy at 5 years in adults aged 35-74 years | Age (per year) OR 0.94 (0.88 to 1.02, p = 0.134)<br>Age is not associated with increased risk of myopic maculopathy progression at 5 years in adults aged 35-74 years. |
| Irving et al. (2016)<br>(Canada)<br>n = 2656 | Age | Significant change in optical status (see Table 9 for full description) in all ages | Age (per year) OR 1.03 (1.03 to 1.04)<br>Increasing age is associated with increased risk of experiencing a significant change in ocular status. |
| Kang et al. (2012)<br>(USA)<br>n = 120,146 | Age | Incident exfoliation glaucoma or exfoliation glaucoma suspect in adults | Rate ratio (RR) of age:<br>40 to 55 years (ref)<br>55 to 60 years RR 4.33 (2.19 to 8.56)<br>60 to 65 years RR 10.43 (5.50 to 19.78)<br>65 to 70 years RR 19.88 (10.41 to 37.96)<br>70 to 75 years RR 33.54 (17.23 to 65.29)<br>Over 75 years RR 46.22 (22.77 to 93.80)<br>Increasing age is associated with higher risk for incident exfoliation glaucoma or exfoliation glaucoma suspect. |
| Keel et al. (2017)<br>(Australia)<br>n = 3098 | Age | Rates of eye care referral in adults aged 50 years and over | Age OR 1.02 (1.01 to 1.02, p < 0.001)<br>Increasing age is associated with higher eye care referral rates in adults aged 50 years and older. |
| Khachatryan et al. (2015)<br>(USA)<br>n = 357 | Age | Incident visual field damage in glaucoma suspects of African or European descent | Age (per year) HR 1.02 (0.99 to 1.04)<br>Age is not associated with increased risk of visual field damage. |
| Marcus et al. (2012)<br>(The Netherlands)<br>n = 3939 | Age | Incident OAG in adults aged 55 year and over | Age (per year) OR 1.06 (1.04 to 1.09, p < 0.001)<br>Age is associated with increased risk of incident OAG in adults aged 55 years and older. |
| Stingl et al. (2023)<br>(Germany)<br>n = 10,175 | Age | Change in refractive error at 5 years in adults aged 35-74 years | Myopic change at 5 years:<br>Age (per year) OR 0.52 (0.49 to 0.55, p <0.001)<br>Hyperopic change at 5 years:<br>Age (per year) OR 1.62 (1.52 to 1.72, p <0.001)<br>Decreasing age is associated with increased risk of having a myopic shift in refractive error at 5 years in adults aged 35 to 74 years.<br>Increasing age is associated with an increased risk of a hyperopic shift in refractive error at 5 years in adults aged 35 to 74 years. |
| Ekström & Hårleman (2023)<br>(Sweden)<br>n = 481 | Sex | Incident OAG in adults aged 55-84 years | Sex (male) OR 1.77 (0.91 to 3.43)<br>Sex is not associated with increased risk of OAG in adults aged 55-84 years. |
| Guggenheim et al. (2012)<br>(UK)<br>n = 2005 | Sex | Incident myopia after age 11 | OR 1.058 (0.810 to 1.382, p = 0.679)<br>Sex is not associated with incident myopia after age 11. |
| Hopf et al. (2022)<br>(Germany)<br>n = 350 | Sex | Progression of myopic maculopathy at 5 years in adults aged 35-74 years | Sex (female) OR 5.54 (0.93 to 32.92, p = 0.060)<br>Sex is not associated with increased risk of myopic maculopathy progression at 5 years in adults aged 35-74 years. |
| Irving et al. (2016)<br>(Canada)<br>n = 2656 | Sex | Significant change in optical status (see Table 9 for full description) in all ages | Sex (female) OR 1.07 (0.90 to 1.29)<br>Sex is not associated with increased risk of significant change in ocular status. |
| Kang et al. (2012)<br>(USA)<br>n = 120,146 | Sex | Incident exfoliation glaucoma or exfoliation glaucoma suspect in adults | Rate ratio (RR) of gender:<br>Male RR 0.32 (0.23 to 0.46)<br>Female (ref)<br>Female sex is associated with higher risk for incident exfoliation glaucoma or exfoliation glaucoma suspect. |
| Keel et al. (2017)<br>(Australia)<br>n = 3098 | Sex | Rates of eye care referral in adults aged 50 years and over | Sex (male) OR 1.24 (1.06 to 1.46, p = 0.007)<br>Male sex is associated with higher eye care referral rates in adults aged 50 years and older. |
| Marcus et al. (2012)<br>(The Netherlands)<br>n = 3939 | Sex | Incident OAG in adults aged 55 year and over | Sex (female) OR 0.63 (0.43 to 0.93, p = 0.022)<br>Male sex is associated with increased risk of incident OAG in adults aged 55 years and older. |
| Stingl et al. (2023)<br>(Germany)<br>n = 10,175 | Sex | Change in refractive error at 5 years in adults aged 35-74 years | Sex (female) OR 1.49 (1.28 to 1.73, p < 0.001)<br>Female sex is associated with increased risk of having a myopic shift in refractive error at 5 years in adults aged 35 to 74 years. |
| Khachatryan et al. (2015)<br>(USA)<br>n = 357 | Ethnicity/<br>race | Incident visual field damage in glaucoma suspects of African or | African descent vs. European descent by IOP:<br>No significant association at IOP = 10 mmHg to 20 mmHg<br>IOP 22 mmHg HR 2.03 (1.15 to 3.57)<br>IOP 24 mmHg HR 2.71 (1.39 to 5.29)<br>IOP 26 mmHg HR 3.61 (1.61 to 8.08) |
|  |  | European descent | Mean IOP of cohort (17.8 mmHg) HR 1.12 (0.66 to 1.90)<br>Glaucoma suspects of African descent with higher mean IOP are associated with increased risk of visual field damage compared to European glaucoma suspects. |
| Stem et al. (2013)<br>(USA)<br>n = 494,165 | Ethnicity/<br>race | Incident CRVO in adults aged 55 years and over | Ethnicity:<br>Black HR 1.58 (1.25 to 1.99, p < 0.0001)<br>Asian-American HR 0.75 (0.43 to 1.30, p = 0.31)<br>White (ref)<br>Black ethnicity is associated with higher risk for incident CRVO in adults aged 55 years and over. |
| Keel et al. (2017)<br>(Australia)<br>n = 3098 | Education | Rates of eye care referral in adults aged 50 years and over | Years of education OR 0.98 (0.96 to 1.00, p = 0.11)<br>Years of education is not associated with eye care referral rates in adults aged 50 years and older. |
| Stingl et al. (2023)<br>(Germany)<br>n = 10,175 | Education | Change in refractive error at 5 years in adults aged 35-74 years | Myopic change at 5 years:<br>Secondary general school (ref)<br>Intermediate school OR 0.96 (0.81 to 1.14, p = 0.64)<br>High school OR 0.96 (0.81 to 1.12, p = 0.58)<br>Others OR 0.52 (0.17 to 1.57, p = 0.25)<br>None OR 0.91 (0.28 to 2.94, p = 0.88)<br>Hyperopic change at 5 years:<br>Secondary general school (ref)<br>Intermediate school OR 1.11 (0.98 to 1.26, p = 0.10)<br>High school OR 1.03 (0.91 to 1.16, p = 0.67)<br>Others OR 0.64 (0.27 to 1.52, p = 0.31)<br>None OR 1.20 (0.44 to 3.27, p = 0.72)<br>Education is not associated with increased risk of either myopic or hyperopic shift at 5 years in adults aged 35 to 74 years. |
| Keel et al. (2017)<br>(Australia)<br>n = 3098 | Geographical remoteness | Rates of eye care referral in adults aged 50 years and over | Geographical remoteness OR 1.04 (0.979 to 1.10, p = 0.27)<br>Geographical remoteness is not associated with eye care referral rates in adults aged 50 years and older. |
| Stem et al. (2013)<br>(USA)<br>n = 494,165 | Net worth / wealth | Incident CRVO in adults aged 55 years and over | Household net worth:<br>> \$500,000 HR 0.73 (0.56 to 0.96, p = 0.02)<br>< \$25,000 (ref)<br>Lower household net worth is associated with higher risk for incident CRVO in adults aged 55 years and over. |
| Stingl et al. (2023)<br>(Germany)<br>n = 10,175 | Occupation | Change in refractive error at 5 years in adults aged 35-74 years | ORs ranged from 0.81 to 1.28, all 95% CI included 1.00, p ≥ 0.05 for all.<br>Occupation is not associated with increased risk of either myopic or hyperopic shift at 5 years in adults aged 35 to 74 years. |

#### 2.2.1 Age

Nine identified studies examined the association of age with various ocular/vision conditions, including five prospective cohort analyses, one retrospective cohort analysis, one population-based longitudinal study, one cross-sectional study, and one case-control study. All of the included observational studies were of low to moderate risk of bias, with three studies rated as moderate due to some measures being self-reported and lack of clarity whether the strategy for multivariate model building was appropriate.

Only three studies reported on the same outcome, and found that aging is associated with an increased risk of developing open-angle glaucoma in adults aged 65 to 74 years, 55 to 84 years, and 55 years or over, respectively (Ekström 2012, Ekström & Hårleman 2023, Marcus et al. 2012). The two studies by Ekström were, however, identified by the review team as having a high risk of double reporting. Kang et al. (2012) found that increasing age is also a risk factor for exfoliation glaucoma or being an exfoliation glaucoma suspect. When compared to 40 to 55 year olds, the rate ratio increased for every 5-year bracket, with 55 to 60 year olds having approximately four times (rate ratio 4.33) the risk of developing exfoliation glaucoma or being a suspect case and those over 75 years old having approximately 46 times the risk (rate ratio 46.22).

Aging is also associated with increased odds of needing an eye care referral in adults over 50 years of age (Keel et al. 2017) or experiencing any kind of change in ocular status (Irving et al. 2016). For every one-year increase in age, the odds of having a change in ocular status increased by 3%.

Aging is associated with a reduced risk of experiencing a myopic change in refractive error at five years in adults aged 35 to 74 years, by 48% per year (Stingl et al. 2023). At the same time, it is associated with increased risk of experiencing a hyperopic change in refractive error over five years in adults aged 35 to 74 years, with 62% increased risk per year (Stingl et al. 2023).

Age was found not to be associated with the risk of progression of myopic maculopathy in high myopes (people with extreme or severe near-sightedness) aged between 35 and 74 (Hopf et al. 2022) or with developing visual field damage in glaucoma suspects of African or European descent (Khachatryan et al. 2015).

#### 2.2.2 Sex

Eight studies were identified that examined the effect of sex as a prognostic factor for a change in ocular status. These included four prospective cohort studies, one retrospective cohort analysis, one longitudinal study, one cross-sectional study and one case-control study. All included studies were identified as having low to moderate risk of bias, with concerns around the use of self-reported measures in four studies.

Male sex was found to be a risk factor for requiring an eye care referral in adults over 50 years of age, with 24% higher odds than females (Keel et al. 2017). Males were also found to be at higher risk of developing open-angle glaucoma in adults aged 55 years and older, with 37% higher risk (Marcus et al. 2012). However, females are more likely to be diagnosed with exfoliation glaucoma or a suspect case of this, with males having less (32%) chance than females do (Kang et al. 2012). Females are also nearly 50% more likely than males to experience a myopic change in refractive error over five years in adults aged 35 to 74 years (Stingl et al. 2023).

Sex was not found to be associated with the risk of developing myopia in children (Guggenheim et al. 2012), or open-angle glaucoma in adults aged 55 to 84 years (Ekström & Hårleman 2023), progression of myopic maculopathy in high myopes aged 35 to 74 years (Hopf et al. 2022), or experiencing a change in ocular status (Irving et al. 2016).

#### 2.2.3 Ethnicity/race

Two identified studies looked into the effect of ethnicity/race in relation to different eye conditions. This included a prospective cohort analysis and a retrospective cohort analysis. Both observational studies were judged to be at low risk of bias.

Glaucoma suspects, defined as eyes with a history of elevated intraocular pressure (IOP) and/or an optic disc appearance suspicious of glaucoma but normal visual fields at baseline in this study, of African descent were at higher risk of developing visual field damage than those of European descent if their mean IOP was 22 mmHg or higher with the hazard ratio increasing as IOP increased (Khachatryan et al. 2015). The study found that those with a mean IOP of 22 mmHg had double the risk of their European counterparts, whilst the risk was more than 3.5 times greater with a mean IOP of 26 mmHg. The study found that there was no significant association with race at IOPs of 10 to 20 mmHg.

Black ethnicity was also associated with increased risk of developing central retinal vein occlusion (CRVO) compared to White ethnicity in adults aged 55 years and over (hazard ratio 1.58 [95% confidence interval (CI) 1.25 to 1.99]) (Stem et al. 2013). Asian-American ethnicity was not deemed to be a risk factor according to this study.

#### 2.2.4 Socioeconomic characteristics

Various socioeconomic factors were examined as potential prognostic factors in three identified studies. These were a prospective cohort study, a retrospective cohort study, and a cross-sectional study. All three studies were rated as low or low to moderate risk of bias.

Geographical remoteness and years of education were not found to be associated with the risk of eye care referral in adults over 50 years of age by Keel et al. (2017). This study was conducted in Australia and, thus there are concerns about the generalisability of this evidence to Wales due to much greater remoteness and distances to major urban settlements in Australia.

Education was found to not be associated with change in refractive error at five years in adults aged 35 to 74 years (Stingl et al. 2023). This same study also found that occupation is not associated with change in refractive error.

Lower household net worth was found to be associated with increased risk of developing CRVO in adults 55 years and older in an American study (Stem et al. 2013). Those with a household net worth of greater than US$500,000 had 27% lower risk of developing CRVO than those with a net worth less than US$25,000 (hazard ratio 0.73 [95% CI 0.56 to 0.96]).

#### 2.2.5 Bottom line results for demographic prognostic factors

The evidence identifies suggests that age, sex, ethnicity, and household net worth are potential risk factors for changes in vision or ocular health. Aging and increasing age is associated with a general increased risk of change in ocular status, while sex, ethnicity and household net worth are dependent on the outcome examined. No studies were identified that examined the use of index of deprivation as a prognostic factor.

Across all studies in this review the certainty of the evidence is low due to the paucity of evidence for each outcome – with only one study identified in many cases. Though the evidence was deemed to be at low or moderate risk of bias, further research is necessary to inform any decision making in this area.

### 2.3 Ocular prognostic factors

Results for this section are summarised in Table 2 with comprehensive details available in Section 6.2, Tables 8 and 9.

**Table 2:**
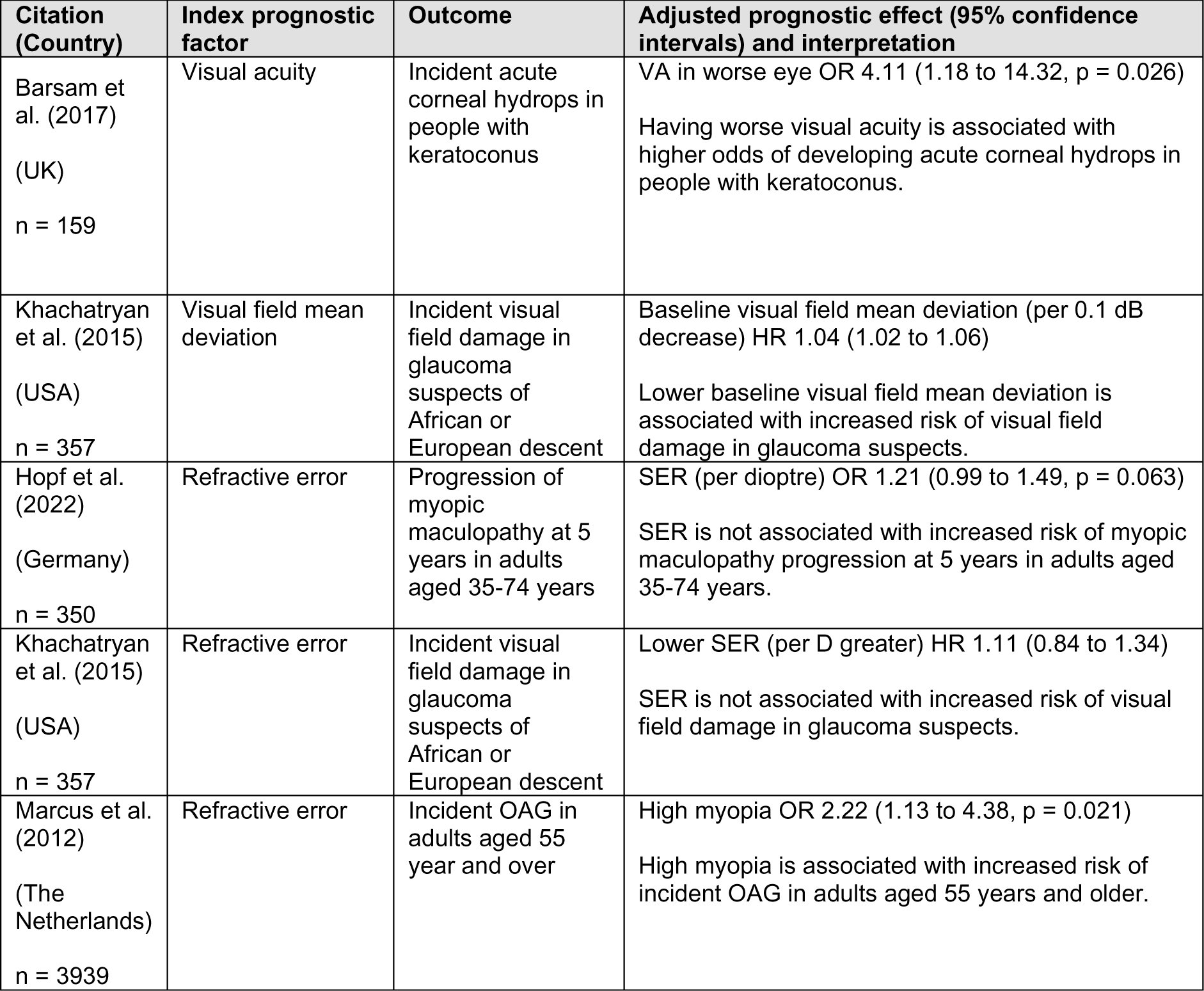
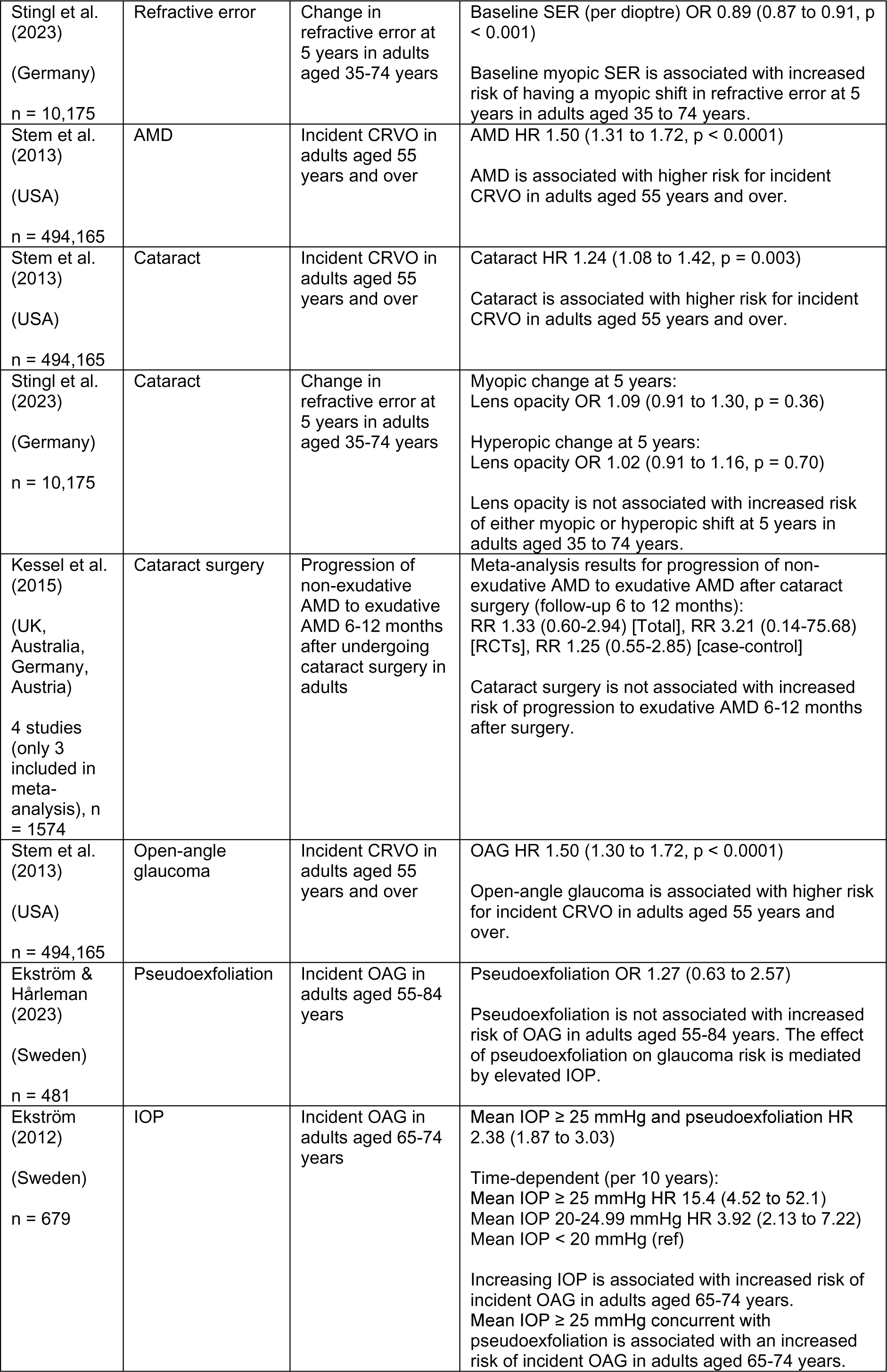
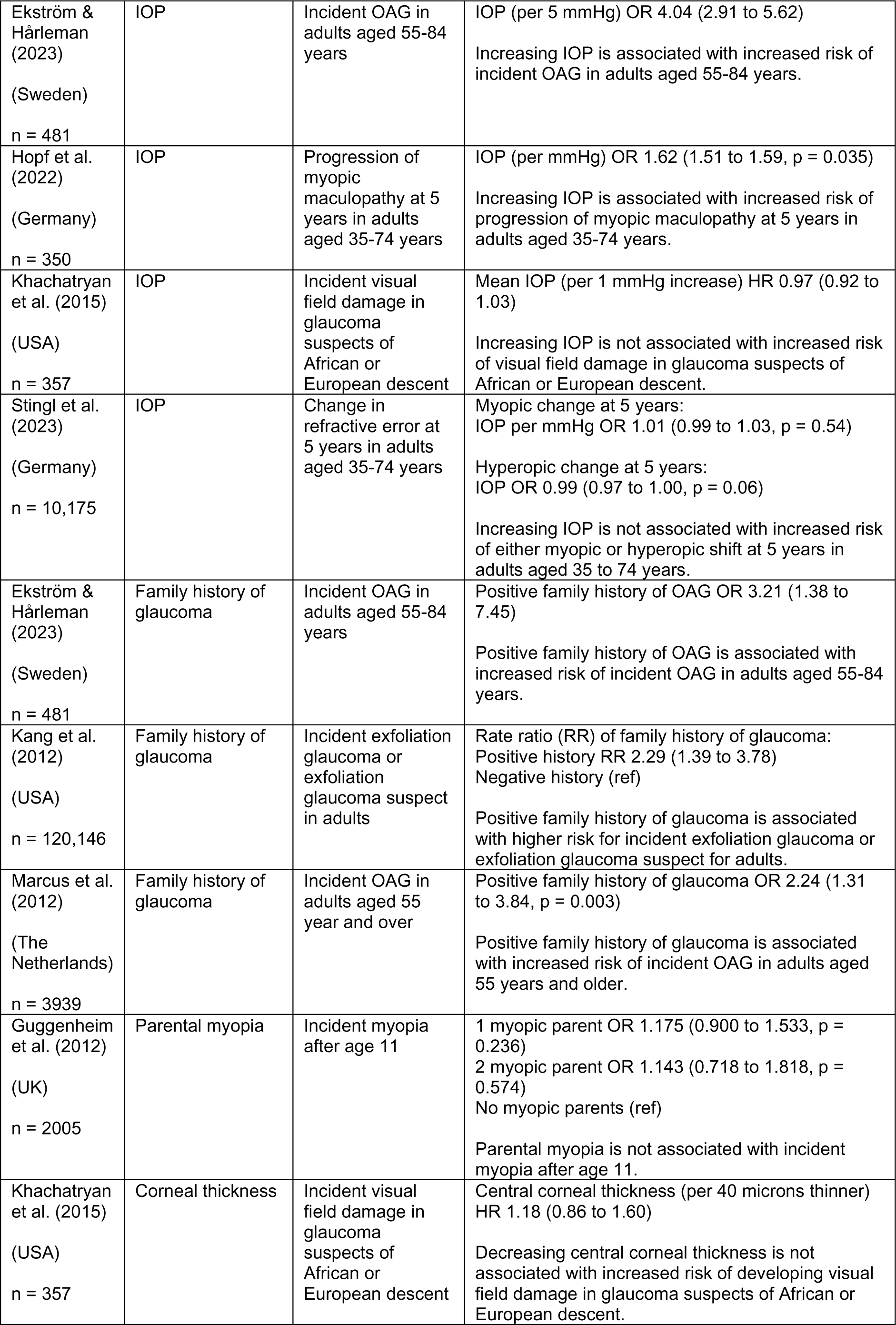
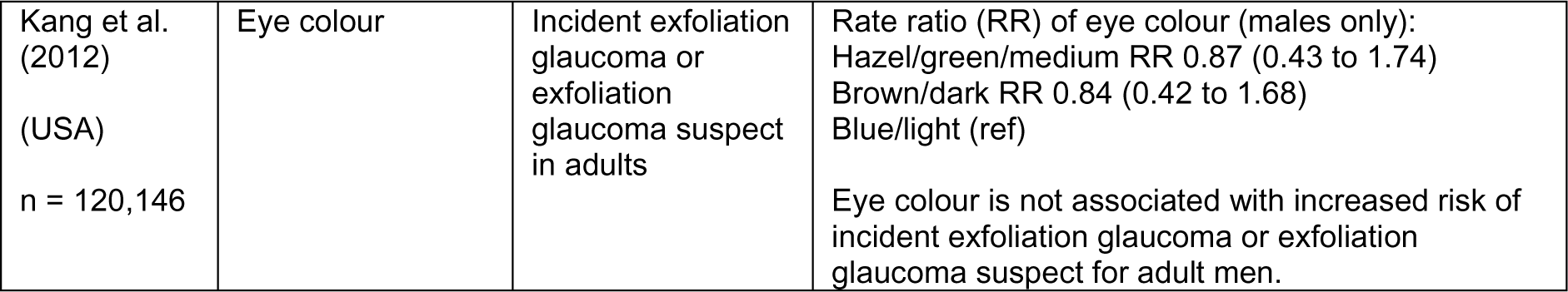
Summary of ocular prognostic factors.

#### 2.3.1 Vision-related

Five identified studies looked at vision-related characteristics as prognostic factors for changes in ocular status. Four were prospective cohort studies and one was a case-control study. Three of the studies were rated as low to moderate risk of bias, with one (Barsam et al. 2017) deemed high risk of bias, due to the case-control study design meaning that the prognostic data was collected after the outcome was known and the low number of cases (21%) included in the multivariate model.

In a case-control study comparing people with keratoconus, a progressive condition causing thinning and irregular curvature of the cornea, who developed acute corneal hydrops (a sight-threatening complication of keratoconus that can leaving scarring) to those who did not, Barsam et al. (2017) found that worse visual acuity (VA) was associated with increased odds of developing corneal hydrops.

A prospective cohort study by Khachatryan et al. (2015) of glaucoma suspects found that a worse result on visual field assessment (mean deviation) at baseline was a risk factor for developing visual field damage (hazard ratio 1.04 (95% CI 1.02 to 1.06) per 0.1 dB decrease).

In a study by Stingl et al. (2023), baseline spherical equivalent refraction (SER) was found to be a risk factor for having a myopic shift in refractive error at five years in adults aged 35 to 74 years, with increasing myopia being a greater risk factor for a myopic shift (11% per dioptre more myopic). However, SER was not found to be associated with the risk of developing visual field damage in glaucoma suspects (Khachatryan et al. 2015) or for the progression of myopic maculopathy in high myopes aged 35 to 74 years (Hopf et al. 2022). High myopia was found to be associated with increased risk of incident open-angle glaucoma in adults aged 55 years and over (Marcus et al. 2012).

#### 2.3.2 Ocular pathology

Four studies identified looked at various ocular pathologies as prognostic factors for other ocular pathology or changes in refractive error. This included one systematic review and meta-analysis, one prospective cohort analysis, one retrospective cohort analysis, and one case-control study. The meta-analysis was judged to have low concerns for risk of bias and the three observational studies were judged to be of low to moderate risk of bias, with two studies unclear whether the strategy for model building was appropriate.

Stem et al. (2013) investigated whether age-related macular degeneration (AMD), open-angle glaucoma, and cataract were prognostic factors for developing CRVO in adults of 55 years of age and older. They found that all three were associated with increased risk of developing CRVO with increased risks of 50%, 50% and 24% respectively compared to those without these conditions.

It was found that the presence of cataract is not associated with increased risk of a change in refractive error in adults aged 35 to 74 years (Stingl et al. 2023), and a meta-analysis of four studies found that undergoing cataract surgery was not associated with increased risk of progression to wet AMD in people with dry AMD 6 to 12 months after surgery (Kessel et al. 2015). Stingl et al. note that their findings are contrary to other cohort studies which report an association between nuclear cataract and a myopic shift in refractive error. They suggest this may be explained by the lack of differentiation of nuclear and cortical cataract in their study.

The presence of pseudoexfoliation was found not to be a risk factor for developing open-angle glaucoma in adults aged 55 to 84 years; odds ratio 1.27 (95% CI 0.63 to 2.57) (Ekström & Hårleman 2023). This is thought to be due to the strong interaction between increased IOP and the presence of pseudoexfoliation (see Section 2.3.3) and, therefore, pseudoexfoliation is not independently associated with increased risk of open-angle glaucoma.

#### 2.3.3 Intraocular pressure

Five identified studies investigated IOP as a prognostic factor for ocular pathology or a change in refractive error. There were three prospective cohort analyses, one longitudinal study, and a case-control study. All studies were assessed as low to moderate risk of bias.

Ekström (2012) and Ekström & Hårleman (2023) confirmed that increasing IOP is associated with increased risk of open-angle glaucoma, something which is well established as stated in the studies. While pseudoexfoliation is not independently associated with glaucoma, mean IOP greater than or equal to 25 mmHg concurrent with pseudoexfoliation is associated with greater risk of incident open-angle glaucoma in 65 to 74 year olds (Ekström 2012).

A study of glaucoma suspects by Khachatryan et al. (2015) found that increasing IOP was not independently associated with increased risk of developing visual field damage in suspects of African or European descent; HR 0.97 per 1 mmHg increase (95% CI 0.92 to 1.03). However, as stated in Section 2.2, the risk of visual field damage in glaucoma suspects of African descent compared to European descent did show a positive correlation with mean IOP increase.

Hopf et al. (2022) found that increasing IOP increased the risk of progression of myopic maculopathy in high myopes aged 35 to 74 years by 62% per mmHg at 5 years. Whilst another study found that IOP is not associated with increased risk of a change in refractive error at 5 years in adults aged 35 to 74 years (Stingl et al. 2023).

#### 2.3.4 Family history

Four studies were identified that investigated family history as a potential prognostic factor. These include two prospective cohort studies, a longitudinal study, and a case-control study. All three studies were judged as low to moderate risk of bias, with self-reporting of measures being a factor.

Three of the studies investigated positive family history of glaucoma as a risk factor for different types of glaucoma. Positive family history of glaucoma was found to be associated with over double (rate ratio 2.29) the risk of developing exfoliation glaucoma or becoming an exfoliation glaucoma suspect in adults (Kang et al. 2012). Positive history was associated with more than double (OR 2.24) the risk of developing open-angle glaucoma in adults aged 55 years and over in a study from The Netherlands (Marcus et al. 2012) and more than three times (OR 3.21) increased risk in adults aged 55 to 84 years (Ekström & Hårleman 2023).

Positive parental history of myopia was not associated with increased risk of children becoming myopic after age 11 years (Guggenheim et al. 2012). This was true for children with only one myopic parent or both parents.

#### 2.3.5 Ocular parameters

Two prospective cohort studies were identified that examined other ocular parameters as potential prognostic factors. One study was rated as low risk of bias, whilst Kang et al. (2012) was rated low to moderate risk of bias due to self-reporting of both outcomes and prognostic factors.

It was found that central corneal thickness is not associated with risk of developing visual field damage in glaucoma suspects of African or European descent (Khachatryan et al. 2015). Eye colour was also found not to be associated with the risk of developing exfoliation glaucoma or becoming a exfoliation glaucoma suspect in adult men (Kang et al. 2012).

#### 2.3.6 Bottom line results for ocular prognostic factors

The evidence identified shows that various ocular prognostic factors, which could be identified during a routine eye examination, were identified as risk factors for the development other ocular pathology or changes in vision or refractive error. This includes VA, visual field mean deviation, SER, various ocular pathologies, IOP, and family history of glaucoma.

As with Section 2.2, it is pertinent to note that the evidence summarised in this section is specific to the outcomes mentioned above, and cannot be extrapolated to cover all eye conditions. In many cases, a lack of evidence means that there is often only one study per prognostic factor / outcome pairing, and therefore the certainty of the evidence is low. There were two studies that reported positive family history of glaucoma as a risk factor for incident open-angle glaucoma and the confidence in this finding is also higher. Most included primary studies were of low or moderate risk of bias, with one at high risk of bias, whilst the included meta-analysis states that the studies it included were of moderate or very low quality.

### 2.4 Interval between eye examinations

Results for this section are summarised in Table 3 with comprehensive details available in Section 6.2, Tables 8 and 9.

**Table 3:** Summary of studies examining interval between eye examinations as a prognostic factor.

| Citation (Country) | Index prognostic factor | Outcome | Adjusted prognostic effect (95% confidence intervals) and interpretation |
| --- | --- | --- | --- |
| Keel et al. (2017)<br>(Australia)<br>n = 3098 | Interval between eye examinations | Rates of eye care referral in adults aged 50 years and over | Time since last eye examination OR 1.15 per year (1.12 to 1.19, p < 0.001)<br>Longer time period since last eye examination is associated with higher eye care referral rates in adults aged 50 years and older. |
| Irving et al. (2016)<br>(Canada)<br>n = 2656 | Interval between eye examinations | Significant change in optical status (see Table 9 for full description) in all ages | Interval between eye examinations (per year) OR 1.06 (1.02 to 1.11)<br>Increasing length of time between eye examinations is associated with increased risk of experiencing a significant change in ocular status. |
| Wright et al. (2020)<br>(UK)<br>n = 132,046 | Interval between eye examinations | Referral to a GP in adults aged 60 years and over | Aged 60-69:<br>Delayed eye exam attendance OR 1.30 (1.04 to 1.61)<br>Early eye exam attendance OR 2.86 (2.36 to 3.46)<br>Aged ≥ 70:<br>Delayed eye exam attendance OR 1.07 (1.01 to 1.13)<br>Early eye exam attendance OR 2.72 (2.58 to 2.87)<br>Delayed attendance for eye examinations is associated with increased risk of requiring a GP referral for adults aged 60 years and older. Early attendance is also associated with increased risk of referral for adults aged 60 years or older, though this is driven by early attendance usually being due to symptomatic problems or early recall suggested by the optometrist. |

Three of the identified studies investigated whether the length of time between eye examinations is a prognostic factor for changes in ocular health. This included two retrospective cohort analyses and a cross-sectional study. All three studies were of low or low to moderate risk of bias.

Keel et al. (2017) and Wright et al. (2020) examined whether increased time between eye examinations affected eye care referral rates and rates of referral to a general practitioner (GP), respectively. The odds of requiring an eye care referral increased 15% per year since last examination in the study by Keel et al. (2017) in adults aged 50 years and older. Delayed attendance at eye examinations was associated with 30% increased odds of requiring referral to a GP for 60 to 69 year olds and a 7% increase for those aged 70 years or over (Wright et al. 2020). Early attendance for an eye examination was associated with nearly three times increased odds of requiring GP referral in both age groups. However, this was believed to be due to early attendance usually being in response to the patient noticing symptomatic problems or because the optometrist recommended early assessment at the last examination.

The relationship between the time elapsed since previous eye examination and the odds of experiencing a change in ocular status (such as a change in vision or glasses prescription, emergence of new pathology, requiring a referral) was investigated by Irving et al. (2016). The study found that increasing the length of time elapsed between examinations is associated with an increased risk of experiencing a significant change in ocular status, with a 6% increase in risk per year.

#### 2.4.1 Bottom line results for intervals between eye examinations

Increasing length of time between eye examinations is associated with increased risk of experiencing a change in ocular status, including requiring onward referral. The confidence in this finding is high as three studies reported this as a prognostic factor for similar outcomes. Due to the study designs of Keel et al. (2017) and Wright et al. (2020), it is possible that some of the participants in these studies were experiencing symptomatic eye issues.

### 2.5 Lifestyle/behaviour prognostic factors

Results for this section are summarised in Table 4 with comprehensive details available in Section 6.2, Tables 8 and 9.

**Table 4:**
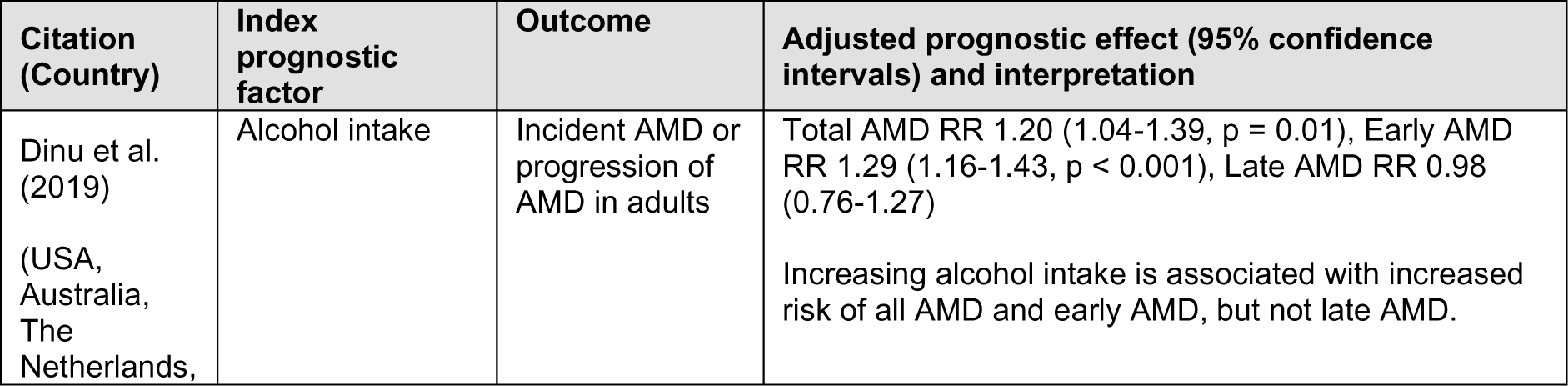
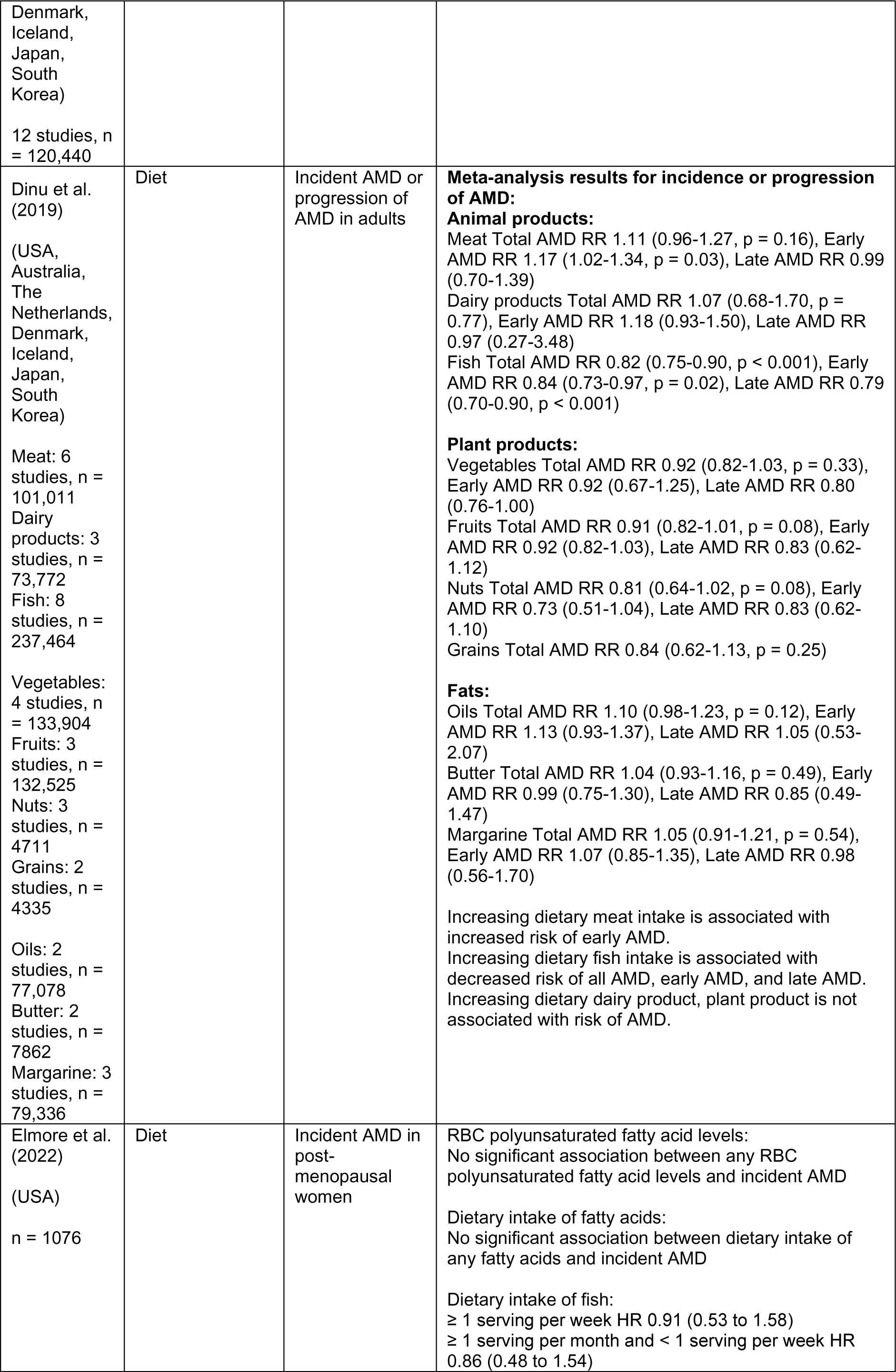
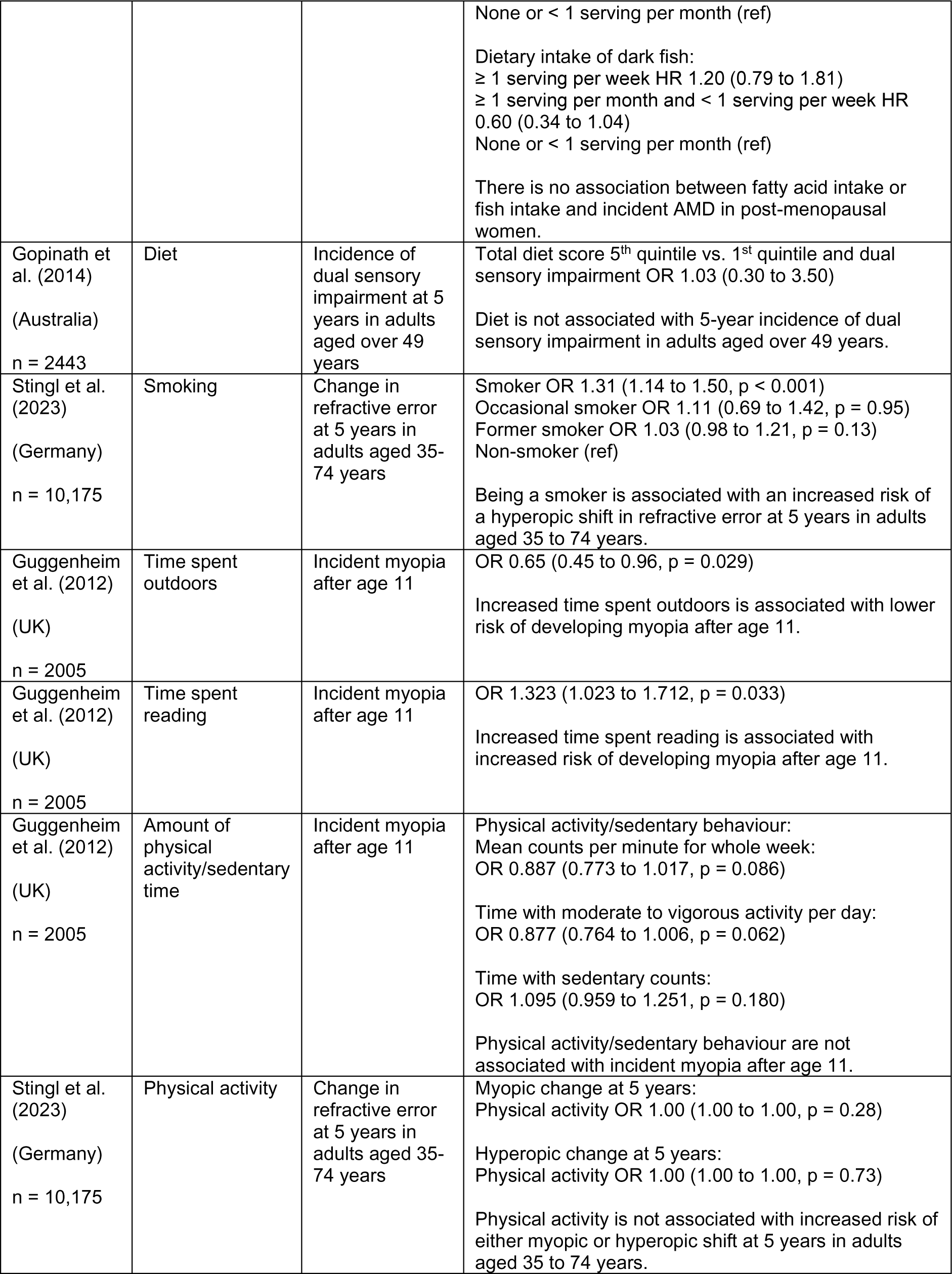
Summary of lifestyle/behaviour prognostic factors.

#### 2.5.1 Diet

Three of the identified studies investigated diet or alcohol intake as prognostic factors for changes in ocular status. This included one systematic review and meta-analysis, one prospective cohort study, and one longitudinal population-based cohort study. The two observational studies were rated as moderate risk of bias due to uncertainties around study attrition and some self-reporting of measures. The meta-analysis was rated as having unclear concerns of risk of bias due to heterogeneity in the studies not being addressed and a lack of clarity as to whether subgroup analyses were pre-specified.

In a meta-analysis of 26 studies, Dinu et al. (2019) found that higher meat intake is associated with higher risk of the occurrence or progression of early AMD, increasing the risk by 17%. Increasing alcohol intake was found to be associated with increased risk of all AMD and early AMD (20% and 29% increased risk respectively) but not with late AMD. Increased dietary intake of fish was found to have a protective effect against AMD with the risk of early AMD being reduced by 16%, late AMD by 21%, and all AMD by 18%. An increased intake of dairy products, plant products, and fats was not found to be associated with risk of AMD.

In a study on post-menopausal women, Elmore et al. (2022) found no association between dietary intake of fish or fatty acids and incident AMD. The study also investigated red blood cell fatty acid levels as a longer-term biomarker of fatty acid intake and still found no association between levels of any red blood cell polyunsaturated fatty acid levels and incident AMD.

Gopinath et al. (2014) investigated the effect of diet on the 5-year incidence of dual sensory impairment (concurrent visual and hearing impairment). The study found no association between having a higher total diet score (healthier diet) and the incidence of dual sensory impairment in adults aged over 49 years.

#### 2.5.2 Smoking

One identified study investigated smoking as a prognostic factor. This was a prospective cohort study. The study was rated as low to moderate risk of bias due to issues with study attrition and lack of clarity on whether the model building strategy was appropriate.

Smoking is associated with increased risk of experiencing a hyperopic change in refractive error at five years in adults aged 35 to 74 years (Stingl et al. 2023), with the risk increasing by 31%. However, the study found that being an occasional smoker or former smoker does not increase the risk of having a hyperopic change in prescription compared to non-smokers.

#### 2.5.3 Activity-related

Two studies were identified that investigated whether activity levels, or the types of activities done, are prognostic factors for refractive changes. One was a prospective cohort analysis and the other was a longitudinal study. Both observational studies were of low to moderate risk of bias, with study attrition being a common issue.

Guggenheim et al. (2012) examined the role of time spent outdoors and physical activity on myopia prevalence and progression. The study found that increased time spent outdoors was independently associated with lower risk of developing myopia after the age of 11 years, with the risk reducing by 35%. It also found that increased time spent reading was associated with a 32% increased risk of developing myopia after age 11, but that the amount of physical activity done/amount of sedentary time was not independently associated with the risk of developing myopia.

Stingl et al. (2023) also found that the amount of physical activity was not associated with increased risk of a change in refractive error at five years in adults aged 35 to 74 years.

#### 2.5.4 Bottom line results for lifestyle/behaviour prognostic factors

Dietary intake of meat, fish, and alcohol are prognostic factors for incident AMD. However, there is discrepancy in the literature on the protective effect of fish intake in the sub-group of post-menopausal women.

Smoking is potentially a risk factor for hyperopic changes in refractive error. Increased time spent outdoors may protect against children developing myopia, whilst reading may be a risk factor for it. However, the amount of time being physically active does not appear to be associated with changes in refractive error.

As before, though all the studies included in this section of the rapid review are of moderate or low risk of bias, the findings are of low certainty due to the fact that there is generally only one study for each prognostic factor/outcome pairing. The findings therefore cannot be generalised beyond the outcome or specific populations that are presented in this section.

### 2.6 Systemic health prognostic factors

Results for this section are summarised in Table 5 with comprehensive details available in Section 6.2, Tables 8 and 9.

**Table 5:** Summary of systemic health prognostic factors.

| Citation (Country) | Index prognostic factor | Outcome | Adjusted prognostic effect (95% confidence intervals) and interpretation |
| --- | --- | --- | --- |
| Ekström & Hårleman (2023)<br>(Sweden)<br>n = 481 | Hypertension | Incident OAG in adults aged 55-84 years | Treated hypertension OR 0.58 (0.29 to 1.15)<br>Treated hypertension is not associated with risk of incident OAG in adults aged 55 to 84 years. |
| Khachatryan et al. (2015)<br>(USA)<br>n = 357 | Hypertension | Incident visual field damage in glaucoma suspects of African or European descent | Mean arterial pressure (per 1 mmHg increase) HR 1.03 (1.00 to 1.06)<br>Increasing mean arterial pressure is associated with increased risk of visual field damage in glaucoma suspects of African or European descent. |
| Stem et al. (2013)<br>(USA)<br>n = 494,165 | Hypertension / high cholesterol / diabetes | Incident CRVO in adults aged 55 years and over | Hypertension HR 1.66 (1.14 to 2.42, p = 0.01)<br>Hypertension and diabetes HR 1.82 (1.15 to 2.89, p = 0.01)<br>Hypertension and hyperlipidaemia HR 1.46 (1.04 to 2.05, p = 0.03)<br>Hypertension, hyperlipidaemia, and diabetes HR 1.58 (1.11 to 2.23, p = 0.01)<br>No diabetes, hypertension, or hyperlipidaemia (ref)<br>Hypertension (alone or in combination with diabetes, hyperlipidaemia, or both) is associated with higher risk for incident CRVO in adults aged 55 years and over. |
| Stem et al. (2013)<br>(USA)<br>n = 494,165 | Peripheral artery disease | Incident CRVO in adults aged 55 years and over | HR 1.15 (1.00 to 1.33, p = 0.05)<br>Peripheral artery disease is associated with higher risk for incident CRVO in adults aged 55 years and over. |
| Stem et al. (2013)<br>(USA)<br>n = 494,165 | Hypercoagulable state | Incident CRVO in adults aged 55 years and over | HR 2.45 (1.40 to 4.28, p = 0.002)<br>Hypercoagulable state is associated with higher risk for incident CRVO in adults aged 55 years and over. |
| Stem et al. (2013)<br>(USA)<br>n = 494,165 | Stroke | Incident CRVO in adults aged 55 years and over | HR 1.44 (1.23 to 1.68, p < 0.0001)<br>Stroke is associated with higher risk for incident CRVO in adults aged 55 years and over. |
| Keel et al. (2017)<br>(Australia)<br>n = 3098 | Stroke | Rates of eye care referral in adults aged 50 years and over | OR 1.00 (0.99 to 1.00, p = 0.64)<br>Previous stroke is not associated with eye care referral rates in adults aged 50 years and older. |
| Stem et al. (2013)<br>(USA) | Myocardial infarction | Incident CRVO in adults aged 55 years and over | HR 0.72 (0.57 to 0.92, p = 0.01)<br>Previous myocardial infarction is associated with lower risk of incident CRVO in adults aged 55 years and over. |
| n = 494,165 |  |  |  |
| Ekström & Hårleman (2023)<br>(Sweden)<br>n = 481 | Ischaemic heart disease | Incident OAG in adults aged 55-84 years | OR 2.41 (1.15 to 5.06)<br>Ischaemic heart disease is associated with increased risk of incident OAG in adults aged 55-84 years. |
| Stingl et al. (2023)<br>(Germany)<br>n = 10,175 | Cardiovascular parameters | Change in refractive error at 5 years in adults aged 35-74 years | ORs ranged from 0.98 to 1.07, all 95% CI included 1.00, $p > 0.05$ for all.<br>Cardiovascular parameters are not associated with change in refractive error at 5 years in adults aged 35 to 74 years. |
| Keel et al. (2017)<br>(Australia)<br>n = 3098 | Diabetes | Rates of eye care referral in adults aged 50 years and over | OR 0.83 (0.67 to 1.04, $p = 0.11$)<br>History of diabetes is not associated with eye care referral rates in adults aged 50 years and older. |
| Fernández-Montero et al. (2017)<br>(Spain)<br>n = 10,401 | Pregnancy | Incident myopia or progression of myopia in women aged 20-50 years | HR 0.58 (0.49 to 0.69, $p < 0.001$)<br>Pregnancy is associated with a decreased risk of developing myopia or progression of existing myopia in women aged 20-50 years. |
| Pasquale & Kang (2011)<br>(USA)<br>n = 79,440 | Age at menarche | Incident OAG in women aged 40 years and over | > 13 years and normal tension glaucoma RR 1.47 (1.01 to 2.13)<br>< 12 years (ref)<br>Age at menarche older than 13 years is associated with increased risk of normal tension glaucoma in women aged 40 years or older. |
| Pasquale & Kang (2011)<br>(USA)<br>n = 79,440 | Reproductive duration | Incident OAG in women aged 40 years and over | < 36 years RR 0.93 (0.71 to 1.22)<br>36-38 years RR 0.94 (0.73 to 1.21)<br>39-40 years (ref)<br>≥ 41 years RR 0.96 (0.73 to 1.27)<br>Reproductive duration is not associated with OAG in women aged 40 years or older. |
| Pasquale & Kang (2011)<br>(USA)<br>n = 79,440 | Oral contraceptives, duration of use | Incident OAG in women aged 40 years and over | Ever used RR 1.14 (0.98 to 1.34)<br>< 2 years RR 1.10 (0.89 to 1.36)<br>2-4 years RR 1.04 (0.81 to 1.35)<br>5+ years RR 1.25 (1.02 to 1.53)<br>Never used (ref)<br>Five years or greater use of oral contraceptives is associated with increased risk of OAG in women aged 40 years or older. |
| Pasquale & Kang (2011)<br>(USA)<br>n = 79,440 | Oral contraceptives, time since discontinuing use | Incident OAG in women aged 40 years and over | ≥ 25 years RR 1.13 (0.91 to 1.40)<br>20-24 years RR 1.06 (0.82 to 1.38)<br>15-19 years RR 1.20 (0.91 to 1.59)<br>< 10 years RR 1.39 (1.01 to 1.91)<br>Never used (ref)<br>Time since discontinuing use of oral contraceptives less than 10 years is associated with increased risk of OAG in women aged 40 years or older. |
| Pasquale & Kang (2011)<br>(USA) | Parity | Incident OAG in women aged 40 years and over | No children RR 0.85 (0.60 to 1.21)<br>1-2 children (ref)<br>3 children RR 1.08 (0.90 to 1.29)<br>4+ children RR 1.00 (0.84 to 1.19) |
| n = 79,440 |  |  | Parity is not associated with OAG in women aged 40 years or older. |
| Barsam et al. (2017)<br>(UK)<br>n = 159 | Atopy | Incident acute corneal hydrops in people with keratoconus | Vernal conjunctivitis OR 15.00 (1.30 to 173.70, p = 0.026)<br>Asthma OR 4.92 (1.22 to 19.78, p = 0.025)<br>History of vernal conjunctivitis and asthma are associated with higher odds of developing acute corneal hydrops in people with keratoconus. |
| Marcus et al. (2012)<br>(The Netherlands)<br>n = 3939 | Corticosteroid use | Incident OAG in adults aged 55 year and over | Ophthalmic steroid use OR 1.04 (0.66 to 1.65, p = 0.86)<br>Inhaled steroid use OR 0.79 (0.42 to 1.48, p = 0.46)<br>Nasal steroid use OR 1.26 (0.74 to 2.13, p = 0.40)<br>Oral steroid use OR 1.03 (0.65 to 1.64, p = 0.89)<br>Ointment steroid use OR 0.70 (0.47 to 1.05, p = 0.086)<br>Corticosteroid use is not associated with OAG incidence in adults aged 55 years and older. |

#### 2.6.1 Cardiovascular/vascular issues

Five of the identified studies examined cardiovascular or vascular issues as prognostic factors for ocular changes. This included two prospective cohort studies, one retrospective cohort study, one cross-sectional study, and one case-control study. All the studies were of low or low to moderate risk of bias.

Treated hypertension was found not to be associated with increased risk of incident open-angle glaucoma in adults aged 55 to 84 years, however the effect of untreated hypertension was not investigated (Ekström & Hårleman 2023). However, increasing mean arterial pressure was associated with increased risk of visual field damage in glaucoma suspects of African or European descent, with a 3% increased risk per 1 mmHg (Khachatryan et al. 2015). Stem et al. (2013) found that hypertension is associated with increased risk of incident CRVO in adults aged 55 years and over. The risk increased by two-thirds compared to those without hypertension. Those with hypertension and high cholesterol still had increased risk of CRVO compared to those without these conditions but lower increased risk than hypertension alone (46% compared with 66%). Having hypertension, diabetes, and high cholesterol lead to CRVO risk increasing by 58%.

Peripheral arterial disease and hypercoagulable state were both found to be associated with higher risk of CRVO, increasing the risk 1.15 and 2.45 times respectively (Stem et al. 2013). Previous stroke was also associated with higher risk of CRVO (HR 1.44 [95% CI 1.23 to 1.68, p < 0.0001). However, self-reported previous stroke was not associated with higher risk of requiring eye care referral in adults aged 50 years or over (Keel et al. 2017).

Previous myocardial infarction was associated with a lower risk of incident CRVO in adults aged 50 years and over (Stem et al. 2013). The chance of developing CRVO was reduced by 28%; this may be due to treatments given after the myocardial infarction has happened. Ischaemic heart disease was also found to be associated with higher risk of incident open-angle glaucoma in adults aged 55 to 84 years, more than doubling the risk (Ekström & Hårleman 2023).

Stingl et al. (2023) found that a range of cardiovascular parameters were not associated with change in refractive error at five years in adults aged 35 to 74 years.

#### 2.6.2 Diabetes

Two of the identified studies examined diabetes as a prognostic factor. These were a retrospective cohort analysis and a cross-sectional study. The retrospective cohort analysis was rated as low risk of bias, whilst the cross-sectional study was rated low to moderate risk due to self-reporting of some prognostic factors and only including covariates that were significant in univariate analysis in the multivariable model.

Having hypertension and diabetes was associated with higher risk of developing CRVO in adults aged 55 years and over, increasing the risk by 82% (Stem et al. 2013). As mentioned in Section 2.5.1, diabetes alongside hypertension and high cholesterol increased the risk of incident CRVO in this population by 58%.

Self-reported history of diabetes was not found to be associated with eye care referral rates in adults aged 50 years and over (Keel et al. 2017).

#### 2.6.3 Women’s health

Two studies were identified that examined various aspects of women’s health/reproductive health as prognostic factors for eye conditions. Both were prospective cohort analyses. Fernández-Montero et al. (2017) was rated as moderate risk of bias due to self-reporting of outcomes and prognostic factors and due to the loss to follow-up rate. Pasquale & Kang (2011) was also rated as moderate risk for similar reasons.

Fernández-Montero et al. (2017) found that pregnancy was associated with lower risk of developing myopia or progression of existing myopia in women aged 20 to 50 years. This risk was decreased by 42% compared to women who were not pregnant and was proposed to be due to increased time spent outdoors during periods of maternity leave.

For women aged 40 and over, age at menarche was associated with increased risk of incident normal tension glaucoma (Pasquale & Kang 2011). Those with age at menarche older than 13 had 47% increased risk compared to those with age at menarche under 12 years. The length of time between menarche and menopause (reproductive duration) was not associated with risk of incident open-angle glaucoma.

Pasquale & Kang (2011) also found that oral contraceptive use was associated with the risk of incident open-angle glaucoma in women 40 years of age and over. Five years or greater use of oral contraceptives and time since discontinuing use of oral contraceptives less than 10 years were associated with increased risk of open-angle glaucoma. The risk was increased by 25% and 39% respectively compared to those who had never used oral contraceptives. Whether a woman had had children or not, and the number of children they have had, was not associated with incident open-angle glaucoma.

#### 2.6.4 Other systemic health issues

In a case-control study examining risk factors for acute corneal hydrops in keratoconics, two atopic conditions were found to be associated with increased risk of developing corneal hydrops (Barsam et al. 2017). History of vernal conjunctivitis increased the risk by 15 times and asthma increased the risk by nearly five times. However, both had very wide 95% CI, suggesting these results are not very precise, and the multivariable model only included 15 cases and 144 controls out of the samples of 64 and 1794 respectively. This study was rated as high risk of bias due to the case-control design and the prognostic data being collected after the outcome was known and the low number of cases included in the multivariable model.

When investigating adults over 55 years of age, Marcus et al. (2012) found that use of any type of corticosteroid medications was not associated with increased risk of incident open-angle glaucoma. They acknowledged that this is contradictory to many other studies’ findings, but also noted studies with consistent findings that often found that corticosteroid use is associated with increased IOP but not necessarily with a diagnosis of glaucoma. This study was rated as moderate risk of bias due to a fairly low number of participants having follow-up data and some measures being self-reported.

#### 2.6.5 Bottom line results for systemic health prognostic factors

There are many systemic health conditions that can be risk factors or protective factors for ocular pathology, refractive error, or the need for eye care referral. These include hypertension, high cholesterol, diabetes, heart disease, peripheral artery disease, stroke, hypercoagulable state, and atopy. However, this list is not exhaustive. Many of these were associated with CRVO.

Factors related to reproductive health, such as current pregnancy, use of oral contraceptives, and age at menarche, are also linked to glaucoma and myopia. Use of corticosteroids was not identified as a risk factor for glaucoma.

All findings for this section are of low certainty due to the lack of evidence available. Further research is necessary to understand the relationship between systemic health factors and visual health. These findings cannot be extrapolated beyond the outcomes or populations explored in this section.

## 3. DISCUSSION

### 3.1 Summary of the findings

The evidence included in this rapid review suggests that increasing age, sex, Black/African ethnicity, increasing IOP, positive family history of glaucoma, increasing length of time between eye examinations, hypertension, and heart disease are potential prognostic factors for a change in ocular health or vision. These were prognostic factors that were investigated in multiple studies; however, certainty in the evidence is low due to the majority of outcomes only being evidenced by one study. Similarly, the majority of studies were undertaken in specific populations, meaning that the association between these prognostic factors and the individual outcomes remains unclear in the general population. Single studies suggest that lower household net worth, worse VA, worse visual field mean deviation, SER, high myopia, AMD, glaucoma, cataract, diet, increasing alcohol intake, smoking, time spent outdoors, time spent reading, cholesterol, diabetes, peripheral arterial disease, hypercoagulable state, stroke, pregnancy, age at menarche, oral contraceptive use, and atopy are potential prognostic factors for a change in ocular status. This is summarised in Table 6.

**Table 6:**
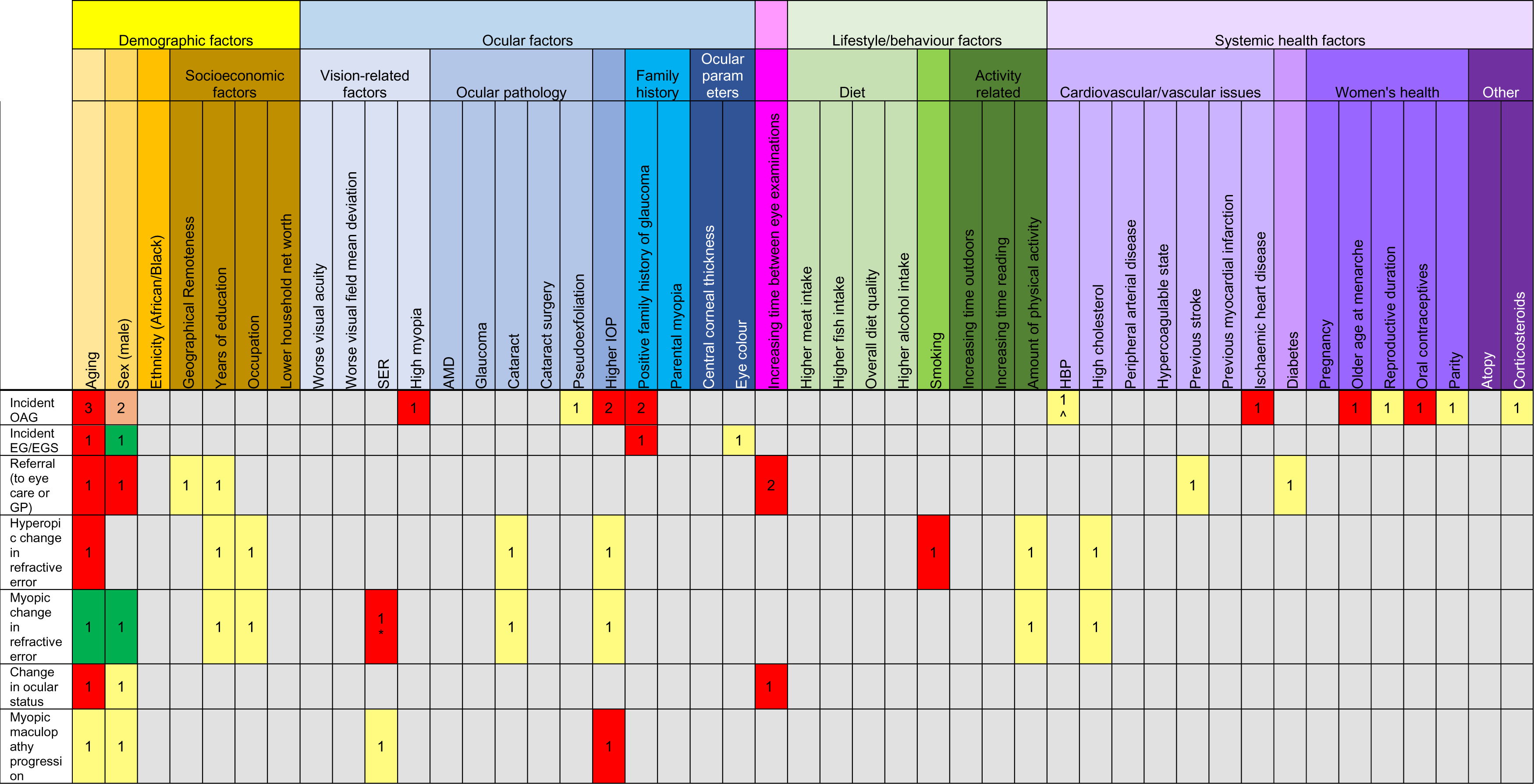
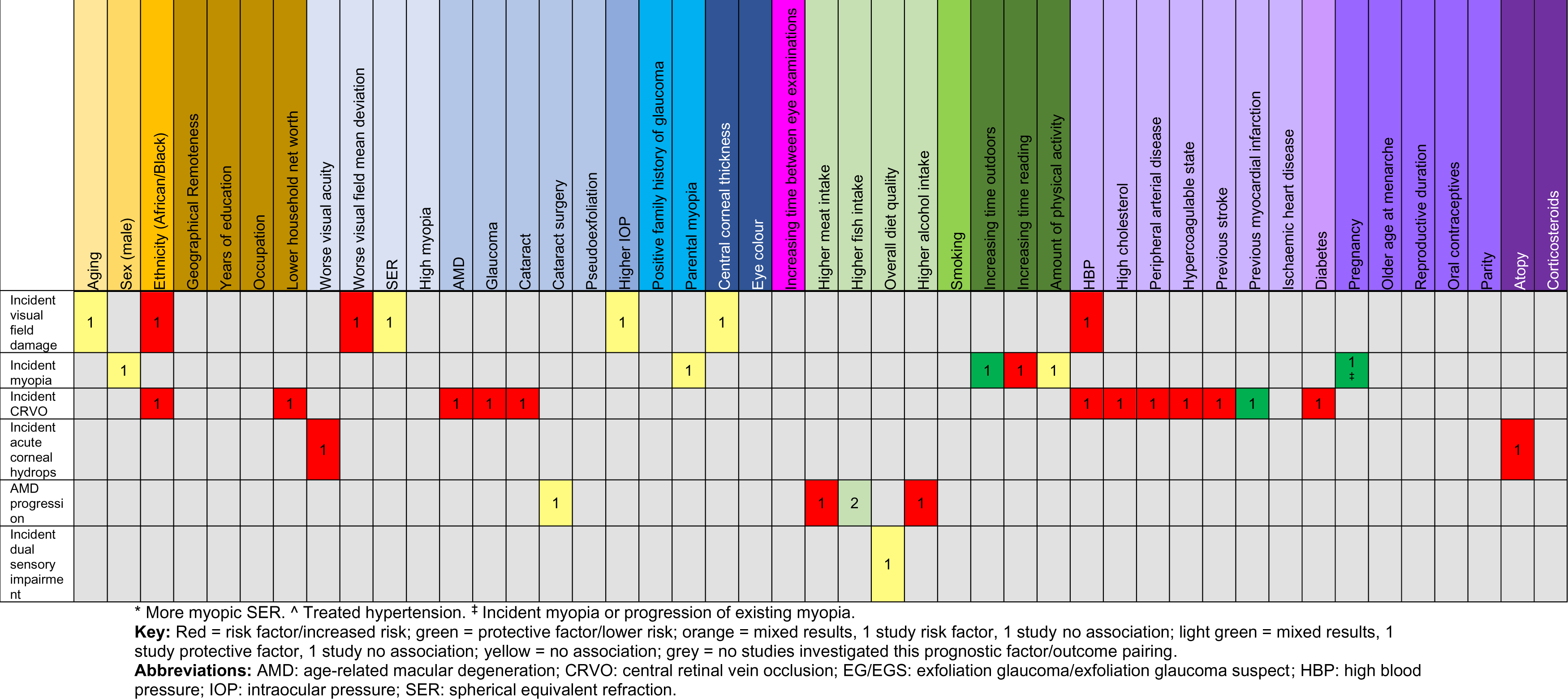
Summary of findings and numbers of studies reporting each finding.

Limited confidence in the results of this rapid review mean that these prognostic effects have limited applicability to the general population, owing to the specificity of the studies. Caution should therefore be taken when drawing from this rapid review, and further research is necessary to inform policy and practice.

It should also be noted that these findings are not specific to risk factors, with some studies also identifying protective factors. Each prognostic factor should be considered in relation to specific outcomes, rather than in relation to the overall category of a change in ocular status.

### 3.2 Strengths and limitations of the available evidence

One of the strengths of the available evidence is that all results were derived from multivariate analyses, presenting adjusting odds / risk / hazard ratios. This demonstrates that the prognostic factor of interest has an independent effect on the outcome. However, this is limited to only being independent of the other covariates included in the multivariate model, and these are not exhaustive. There is notable variation between studies in how many covariates they included in models, with some studies including as few as three covariates and others more than a dozen. Some studies did not clearly state what their adjustment factors were, or how they had decided which factors to include.

Limitations to the available evidence include only identifying two relevant studies that were carried out in the UK. Though search limits and eligibility criteria were applied to ensure only evidence from countries sufficiently similar to the UK were included in the review, the generalisability of any of these studies to Wales remains uncertain. Sample sizes of the studies also varied considerably with some being quite small, only several hundred participants. The sample populations were also often quite specific, such as post-menopausal women, female graduates, or glaucoma suspects, meaning that it is difficult or not possible to apply these findings to the broader population. There was comparatively less evidence identified that examined prognostic factors for conditions that would not require onward referral and could be managed by an optometrist, potentially influencing their decision on frequency of eye examinations, than conditions that would be referred to be managed by other health professionals.

There was a lack of relevant systematic reviews and meta-analyses identified for this review, which would have helped increase the certainty of the evidence by collating more studies. There were also no RCTs identified. However, the types of primary evidence included in the review were of appropriate design for a prognostic factor review. Observational studies, such as cohort studies and case-control studies, can be useful to these types of review and variation in study designs is to be expected (Riley et al. 2019). The included studies were of reasonable quality, with risk of bias ranging from low to moderate as judged using the QUIPS and ROBIS tools. Only one primary study was rated as high risk of bias. However, the data in a number of studies were subject to bias as it was self-reported from participants or their carers. Questionnaires were used in several studies to collect both prognostic and outcome data, and this may lead to high risk of recall bias.

Finally, there was a distinct lack of evidence identified in children and younger adults. Only three studies included children in their study population, and only one of these reported results specifically for children. Similarly, only a small number of studies included adults aged 18-39 years and no studies reported specifically on younger adults. Further research is needed in these populations.

### 3.3 Strengths and limitations of this Rapid Review

This rapid review was strictly limited to the included studies that were deemed to align with the research question and protocol, the scope of which was broad owing to the exploratory nature of this review. Controls intended to manage the amount of retrieved evidence were used, such as strict exclusion criteria and date limits and as such the methods used in this rapid review have been robust and pragmatic. However, it is crucial to note that due to the nature of rapid review methodology there remains the possibility that additional relevant studies were not identified. Additionally, some identified studies that reported findings relevant to the review question were excluded due to not reporting multivariate results or because the findings were not presented as odds/hazard/risk ratios. Sometimes these results can be converted to ratios, however, due to the nature of rapid review, it was not feasible to conduct this and include these studies. As such, the review team were reliant on interpreting the results of studies that have differing levels of quality and their own limitations. This therefore impacts the confidence of this review’s conclusions.

### 3.4 Implications for policy and practice

The low certainty of the evidence in this review means caution should be taken should this review be used for decision making on appropriate eye examination intervals. Additionally, there is very little data from the UK and, thus, the generalisability of the findings to the Welsh population is uncertain.

This review should be used to identify what are thought to be the key prognostic factors and patient characteristics that could be used when an optometrist is determining an individual patient’s risk of a change in ocular status, and therefore the appropriate interval until their next eye examination and suggesting these for further targeted research and evidence synthesis. The chosen factors or characteristics should be specific and narrow in scope, so that the limitations discussed above are mitigated. Alternatively, further research could be conducted looking at prognostic factors for specific ocular outcomes instead. The implications for future research are discussed in more detail below.

### 3.5 Implications for future research

Any further research undertaken to inform guidance on appropriate eye examination re-assessment intervals should be much narrower in focus to ensure as much relevant and useful evidence as possible is gathered. Prognostic factors or specific ocular conditions of interest potentially need to be investigated individually for their effect on a change in ocular status.

This rapid review focused on incident conditions or progression of existing conditions. Prevalence data and prognostic factors for prevalent eye conditions or vision problems may also be useful for decision makers producing guidance on eye examination intervals and further evidence review could be performed in this area.

It has been noted in previous evidence-based guidelines on eye examination frequency that there is a lack of data in younger adults aged under 40 years (Robinson et al. 2012). This was also found to be the case during this review. Therefore, further research is required in this demographic. This is a demographic that is assumed to be at lower risk of ocular issues, but published evidence is lacking to support this claim and research should be conducted to determine if this is the case.

This review has identified a significant lack of evidence that would be needed to make confident conclusions to the research question and represents the findings of evidence that may not be generalisable to Wales, limiting the validity of this review’s conclusions. More high-quality research must be undertaken in the populations of interest in order to inform and guide policy.

### 3.6 Economic considerations*

- Sight loss costs the UK economy £25 billion per annum (RNIB 2021).
- Over 2 million people in the UK are currently living with sight loss (NHS 2021).
- The economic implications of appropriate or inappropriate testing intervals for different causes of vision loss will be different.
- A new case of age-related macular degeneration (AMD) in an adult aged 50 or over, costs the UK economy £73,350 over the person’s lifetime. Lifetime costs to the UK economy for a person diagnosed with glaucoma are approximately £49,800 per person. Reducing the prevalence of these conditions by just 14 or 20 cases respectively could save the UK economy £1 million in lifetime costs (Fight for Sight 2020).
- On economic grounds, early detection of AMD in eye care services and the eye care pathway may be of benefit due to the level of prevalence and associated long term costs to the NHS as the condition causes irreversible, life limiting damage (Stahl 2020, Pezzullo et al. 2018).
- Draft National Institute for Health and Care Excellence (NICE) Guidance for Diabetic Retinopathy (DR) examinations suggest the use of ultrawide-field imaging for diagnosing and monitoring progression. The new guidance publishes in early 2024. DR costs the UK economy £80 million per annum when adjusted to October 2023 prices** (Hex et al. 2012).
- The earlier detection of eye conditions through regular screening can identify conditions before severely impactful symptoms manifest. When captured at a population wide scale, this can result in significant economic savings (Fight for Sight 2020).

*\*This section has been completed by the Centre for Health Economics & Medicines Evaluation (CHEME), Bangor University*.

*\*\* Prices adjusted using Bank of England Inflation Calculator*.

## Data Availability

All data produced in the present study are available upon reasonable request to the authors

## Abbreviations

AMD: Age-related macular degeneration
CI: Confidence interval
CRVO: Central retinal vein occlusion
HR: Hazard ratio
IOP: Intraocular pressure
OR: Odds ratio
QUIPS: Quality in Prognostic factor Studies
RCT: Randomised controlled trial
ROBIS: Risk of Bias in Systematic reviews
RR: Risk ratio / relative risk
SER: Spherical equivalent refraction
VA: Visual acuity

## 5. RAPID REVIEW METHODS

### 5.1 Eligibility criteria

**Table 7:** Eligibility criteria.

|  | Inclusion criteria | Exclusion criteria |
| --- | --- | --- |
| **Population** | Asymptomatic adults and children attending for routine eye examinations, including those without refractive error or pre-existing ocular pathology, as well as those with pre-existing, managed ocular conditions. | People attending for eye examinations due to new symptoms. |
| **Index prognostic factor** | Prognostic factors that are available to optometrists during a routine eye examination will be examined in this review, with appropriate sub-group analyses performed.<br>Specific prognostic factors of interest will include, but not be restricted to, age, sex, ethnicity, systemic health conditions (such as diabetes, hypertension), family history of eye disease, health behaviours (such as smoking, display screen use). Other prognostic factors/sub-groups that are identified during the evidence sift and data extraction stages will also be analysed. |  |
| **Comparison** | Not applicable. |  |
| **Outcomes** | Primary outcomes:<br>A change in ocular status:<br>• change in refractive error<br>• change in visual acuity<br>• emergence of new ocular pathology, e.g. glaucoma, cataracts, macular degeneration<br>• ocular signs of new systemic pathology, e.g. Diabetes, hypertension<br>• change in existing ocular pathology<br>• ocular signs suggesting a change in existing systemic pathology<br>• new referral to general practitioner or secondary care<br>Secondary outcomes:<br>• Prevalence of ocular pathology or refractive errors<br>• Rates of disease/condition progression |  |
| **Timing** | Prognostic factor and outcomes measured at baseline and outcomes measured at any follow-up period up to 5 years.<br>This time horizon was chosen as it was considered an appropriate length of time beyond the current most used interval between eye examinations in Wales of two years. |  |
| **Setting** | To be used during primary care eye examinations to determine an individual patient's risk of experiencing a change in ocular status (as detailed in 'Outcome measures') with this risk being used to create dynamic re-examination intervals. |  |
| **Study design** | Evidence-based clinical guidelines, systematic and rapid reviews, controlled trials, cohort analyses or population-based studies. |  |
| **Countries** | We will prioritise studies from the UK and will not look at evidence from other countries where there is thought to be sufficient evidence from the UK.<br>Where more evidence is required, studies from other countries, where optometry services are similarly comparable to Wales, will be prioritised in the following order: Ireland, Australia, New Zealand, Norway, Canada, Sweden, USA, Malta, Austria, Finland, Germany, Spain, The Netherlands, Switzerland.<sup>a</sup> |  |
| **Language of publication** | English. |  |
| **Publication date** | January 2009 to present. |  |
| **Publication type** | Published and preprint. |  |
| **Other factors** | We will include evidence on both prevalence and progression rates of ocular conditions where this is an outcome reported in relevant studies.<br>We will report relevant recommendations for the frequency of eye examinations made by any evidence-based guidelines identified. |  |

<sup>a</sup>Prioritisation is based on similarity to UK optometry services and inclusion of countries in clinical guidelines by Robinson et al. (2012), and on data from the European Council of Optometry and Optics Blue Book \(2020\)

**Definitions: Refractive error** – A common eye disorder when the eye does not clearly focus images, which can usually be corrected by spectacles or contact lenses. The most common types of refractive error are myopia (shortsightedness), hypermetropia/hyperopia (longsightedness), astigmatism and presbyopia (reduced ability to focus on near objects); **Visual acuity** – A person’s ability to recognise small details with precision, also referred to as clarity of vision.

### 5.2 Literature search

Prior to planning this review, a preliminary search for existing reviews was undertaken of Cochrane Database of Systematic Reviews, NIHR Journals Library, Trip database, KSR Evidence, Canadian Agency for Drugs and Technologies in Health, Prospero, PubMed, NICE, SIGN, Epistemonikos, Google Advanced Search, and Google Scholar using the keywords sight test, eye examination, eye test, sight examination, routine, frequency, interval, recall, and time. The findings were presented to the stakeholders and used to refine the scope of the present rapid review, and to inform the methods.

A comprehensive search was conducted to identify any additional English-language reviews from 2009 onwards. An analysis of the text words contained in the title and abstracts, and of the index terms used to describe any relevant reviews already identified were used to inform the search. The full search strategy was designed and run using Ovid Medline and then translated to all other databases:

- CINAHL via the EBSCO platform
- Embase via the Ovid platform
- Cochrane Library database
- Epistemonikos

This was followed by a thorough search for relevant English-language primary studies from 2009 onwards on the following databases:

- CINAHL via the EBSCO platform
- Medline and Embase via the Ovid platform

The full searches for English-language reviews and primary studies can be found in Appendix 1.

Grey literature sources, including websites of key third sector and government organisations, identified by the review team, or provided by Stakeholders were also searched (see Appendix 2).

### 5.3 Reference management

All citations retrieved from the database searches were imported or entered manually into EndNote^TM^ (Thomson Reuters, CA, USA) and duplicates removed by a single reviewer. The citations that remained were exported as a TXT file and imported to Rayyan^TM^ for study selection. Grey literature search results were added to an Excel spreadsheet and cross-checked against the database search results.

### 5.4 Study selection process

Two reviewers screened 20% of titles and abstracts independently. If 20% is equal to less than 200 total records, then the two reviewers will screen 200 records. After this, the level of agreement was assessed with disagreements settled by discussion and consensus. Both reviewers had to achieve at least 80% agreement on screened records before progressing to the next stage. The remaining titles and abstracts were screened by the primary reviewer alone. 20% of full texts were screened by both reviewers, with the same agreement threshold (80%) as before necessary before the remaining records could be screened by the primary reviewer alone. During independent screening, the primary reviewer consulted with the secondary reviewer in the case of any uncertainties.

### 5.5 Data extraction

Data extraction was based on the outlined eligibility criteria. We extracted details/characteristics on study country, study design, number of participants, relevant outcomes (see eligibility criteria) and study settings. The [Checklist for Critical Appraisal and Data Extraction for Systematic Reviews of Prediction Modelling Studies (CHARMS-PF)](https://www.bmj.com/content/bmj/364/bmj.k4597.full.pdf) (Riley et al. 2019) was used to guide data extraction.

Data extraction was completed by individual reviewers and checked by a second reviewer (see Section 6.2 for completed data extraction forms for all included studies). In line with other prognostic factor reviews, data were only extracted from studies that reported prognostic factors as hazard ratios, odds ratios, or risk ratios/relative risk. Only multivariate or adjusted ratios were extracted so that only factors that were independently associated with outcomes were included in the report. Studies were excluded if they only reported unadjusted/univariate results. Relevant prevalence or condition progression rates were also extracted from studies that had reported the ratios listed above.

### 5.6 Quality appraisal

Study quality was assessed using the Risk of Bias in Systematic Reviews (ROBIS) tool (Whiting et al. 2016) for included systematic reviews and using the Quality in Prognostic Factor Studies (QUIPS) tool for included primary studies (Hayden et al. 2013). Critical appraisal was completed by individual reviewers and checked by a second reviewer. Studies of all quality were included.

### 5.7 Synthesis

We undertook narrative synthesis of the evidence identified based on the selection criteria outlined above.

### 5.8 Assessment of body of evidence

All evidence selected after the sift stage was deemed fit for inclusion in the final review. Due to the scope of this review and the methodological constraints of rapid review, formal assessment of the body of evidence using GRADE was not feasible in this case. An informal assessment of the evidence has been conducted.

## 6. EVIDENCE

### 6.1 Search results and study selection

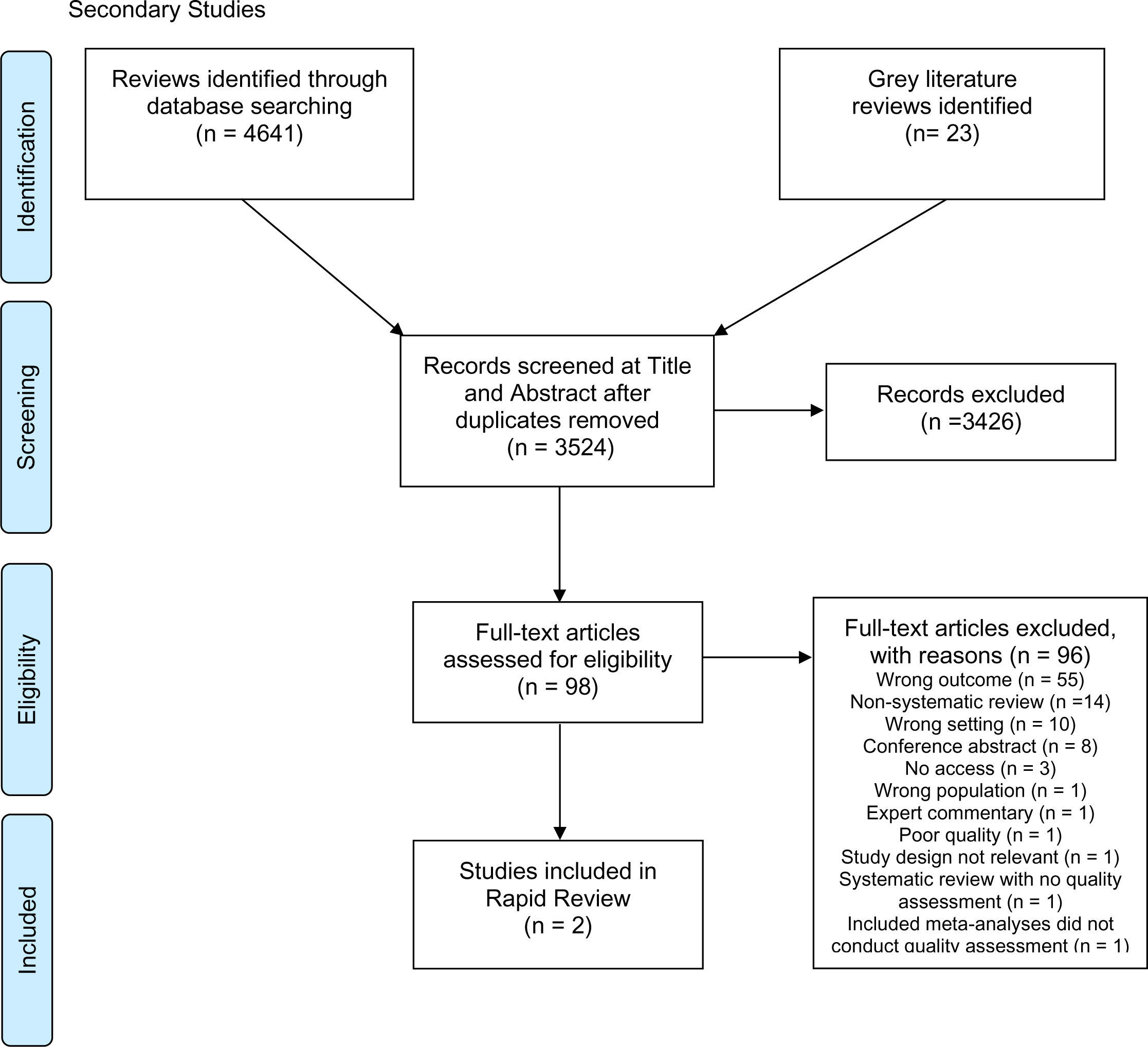

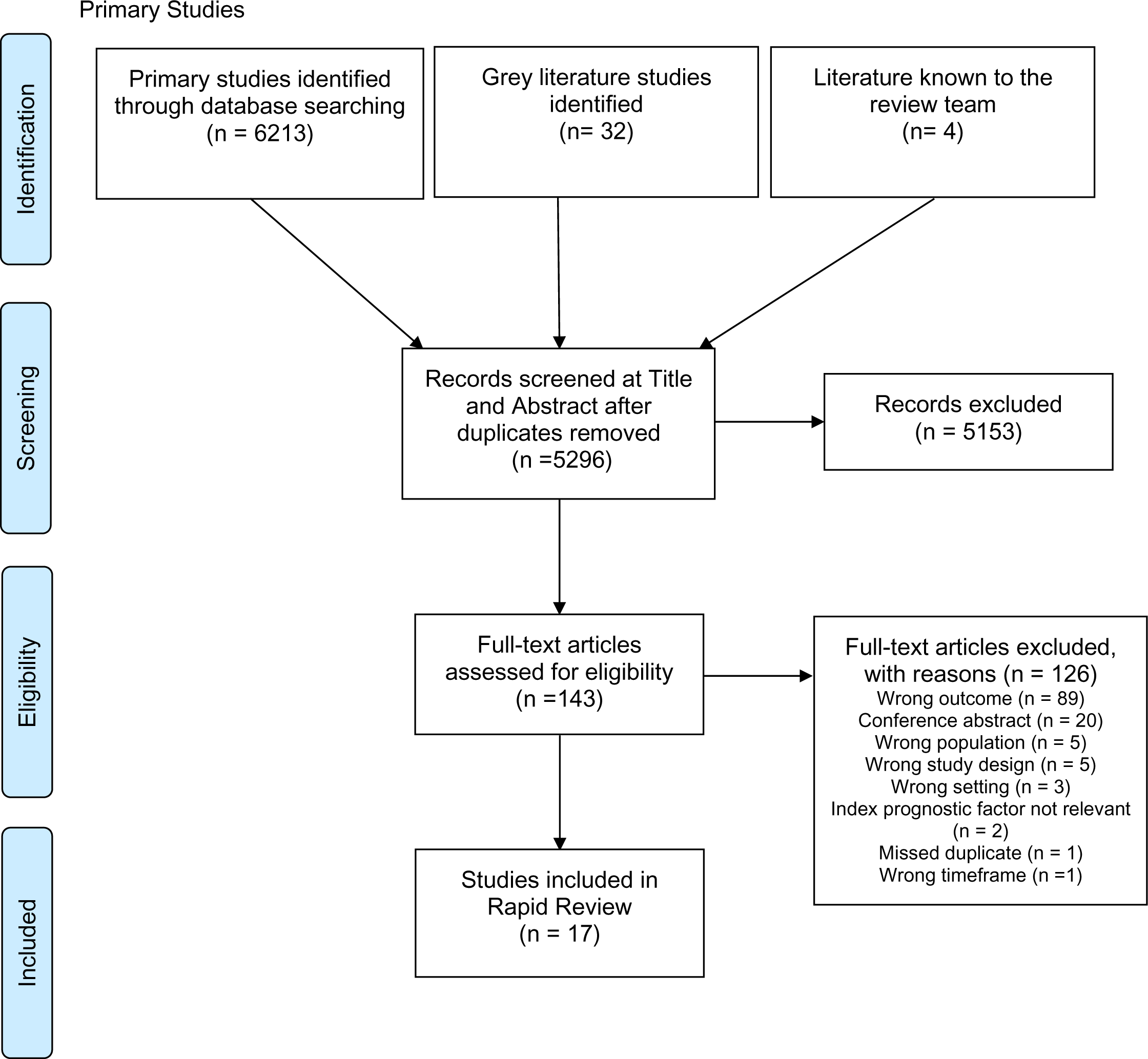

### 6.2 Data extraction

All of the studies included in the rapid review are listed here. This includes two systematic reviews (Table 8) and 17 primary studies (Table 9).

**Table 8:** Summary of included systematic reviews.

| Citation | Review details | Included studies | Quality | Key findings | Interpretation and observations |
| --- | --- | --- | --- | --- | --- |
| **Dinu et al. (2019)** | Review period: 1966 to January 2018<br>Review purpose: To evaluate the consumption of different food groups and alcohol in relation to occurrence and progression of AMD<br>Study designs: Prospective cohort studies<br>Outcome measures: Occurrence of AMD | 26 studies:<br>Meat: 6 studies, n = 101,011<br>Dairy products: 3 studies, n = 73,772<br>Fish: 8 studies, n = 237,464<br>Vegetables: 4 studies, n = 133,904<br>Fruits: 3 studies, n = 132,525<br>Nuts: 3 studies, n = 4711<br>Grains: 2 studies, n = 4335<br>Oils: 2 studies, n = 77,078<br>Butter: 2 studies, n = 7862<br>Margarine: 3 studies, n = 79,336<br>Alcohol: 12 studies, n = 120,440<br>Country: 10 USA, 10 Australia, 2 The Netherlands, 1 Denmark, 1 Iceland, 1 Japan, 1 South Korea | Methodological quality of the included studies was appraised using the Newcastle–Ottawa Scale.<br>All but 3 studies were ranked as high quality.<br>ROBIS RoB assessment:<br>1. Study eligibility criteria: Low risk of bias. Sufficient information on eligibility criteria & their justification<br>2. Identification and selection of studies: Low risk of bias. Search strategy appears appropriate. Dual sifting used<br>3. Data collection and study appraisal: Low risk of bias. Dual independent data extraction. Sufficient data extracted. RoB analysed | **Animal products, RR (95% CI):**<br>Meat Total AMD RR 1.11 (0.96-1.27, p = 0.16), Early AMD RR 1.17 (1.02-1.34, p = 0.03), Late AMD RR 0.99 (0.70-1.39, p NR)<br>Dairy products Total AMD RR 1.07 (0.68-1.70, p = 0.77), Early AMD RR 1.18 (0.93-1.50, p NR), Late AMD RR 0.97 (0.27-3.48, p NR)<br>Fish Total AMD RR 0.82 (0.75-0.90, p < 0.001), Early AMD RR 0.84 (0.73-0.97, p = 0.02), Late AMD RR 0.79 (0.70-0.90, p < 0.001)<br>**Alcohol:**<br>Total AMD RR 1.20 (1.04-1.39, p = 0.01), Early AMD RR 1.29 (1.16-1.43, p < 0.001), Late AMD RR 0.98 (0.76-1.27, p NR)<br>**Plant products:**<br>Vegetables Total AMD RR 0.92 (0.82-1.03, p = 0.33), Early AMD RR 0.92 (0.67-1.25, p NR), Late AMD RR 0.80 (0.76-1.00, p NR)<br>Fruits Total AMD RR 0.91 (0.82-1.01, p = 0.08), Early AMD RR 0.92 (0.82-1.03, p NR), Late AMD RR 0.83 (0.62-1.12, p NR)<br>Nuts Total AMD RR 0.81 (0.64-1.02, p = 0.08), Early AMD RR 0.73 (0.51-1.04, p NR), Late AMD RR 0.83 (0.62-1.10, p NR)<br>Grains Total AMD RR 0.84 (0.62-1.13, p = 0.25) | Increasing dietary meat intake is associated with increased risk of early AMD.<br>Increasing alcohol intake is associated with increased risk of all AMD and early AMD, but not late AMD.<br>Increasing dietary fish intake is associated with decreased risk of all AMD, early AMD, and late AMD.<br>Studies from countries included in the rapid review protocol contributed more than 85% of the weighting to all included meta-analyses. |
|  |  |  | 4. Synthesis and findings: Unclear risk of bias. Serious heterogeneity ($I^2 > 50\%$) not addressed in the synthesis. RoB not addressed for each pooled result. Unclear whether food groups or early vs late AMD subgroups were pre-specified in protocol.<br>5. Risk of bias in the review: Unclear risk of bias. Heterogeneity not addressed in review. Unclear whether subgroup analyses were prespecified | **Fats:**<br>Oils Total AMD RR 1.10 (0.98-1.23, $p = 0.12$), Early AMD RR 1.13 (0.93-1.37, $p$ NR), Late AMD RR 1.05 (0.53-2.07, $p$ NR)<br>Butter Total AMD RR 1.04 (0.93-1.16, $p = 0.49$), Early AMD RR 0.99 (0.75-1.30, $p$ NR), Late AMD RR 0.85 (0.49-1.47, $p$ NR)<br>Margarine Total AMD RR 1.05 (0.91-1.21, $p = 0.54$), Early AMD RR 1.07 (0.85-1.35, $p$ NR), Late AMD RR 0.98 (0.56-1.70, $p$ NR) |  |
| **Kessel et al. (2015)** | Review period: 1996 to August 2014<br>Review purpose: To examine whether cataract surgery increases the risk of progression of dry AMD<br>Study designs: RCTs, case-control studies<br>Outcome measures: Best corrected distance visual acuity, funduscopy signs of AMD progression at least three months after surgery | 4 studies: 2 RCTs, 2 case-control studies. $n = 1679$.<br>Country: UK, Australia, Germany, Austria | Quality was assessed using GRADE.<br>The 2 RCTs were downgraded to 'moderate' quality due to imprecision and the 2 case-control studies were rated as 'very low' due to risk of bias. | **Progression of non-exudative AMD to exudative AMD after cataract surgery (follow-up 6 to 12 months), RR (95% CI):**<br>RR 1.33 (0.60-2.94) [Total], RR 3.21 (0.14-75.68) [RCTs], RR 1.25 (0.55-2.85) [case-control] | Cataract surgery is not associated with increased risk of progression to exudative AMD 6-12 months after surgery. The event rate was low (around 2%), however, and there is uncertainty about the effect estimate.<br>Uncertain whether RRs have been adjusted for confounders. |
|  |  |  | ROBIS RoB assessment:<br>1. Study eligibility criteria: Low risk of bias. Sufficient information on eligibility criteria<br>2. Identification and selection of studies: Unclear risk of bias. Search strategy - seems to very limited - relatively few terms included. Not reported whether there was dual sifting<br>3. Data collection and study appraisal: Low risk of bias. Dual data extraction.<br>4. Synthesis and findings: Low risk of bias.<br>5. Risk of bias in the review: Low risk of bias. |  | One of the RCTs could not have a RR calculated (due to zero events) and, therefore, does not contribute to the meta-analysis. However, it is still reported as being part of the meta-analysis. Many of the excluded studies are then included in a pooled OR, but this is not mentioned in the methodology and the rationale for doing this and including these studies is not given. |

**Abbreviations:** AMD: age-related macular degeneration; CI: confidence interval; NR: not reported; RCT: randomised controlled trial; RoB: risk of bias; RR: risk ratio/relative risk

**Table 9:**
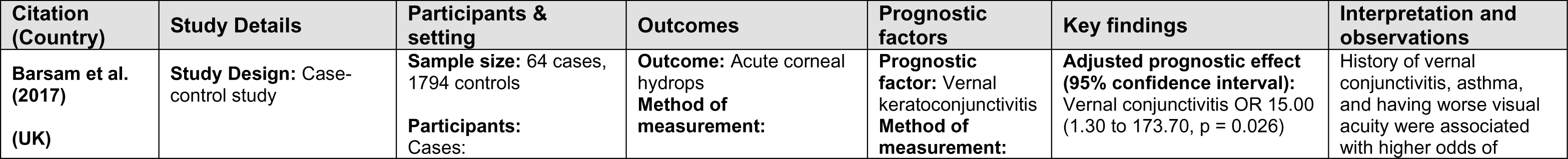
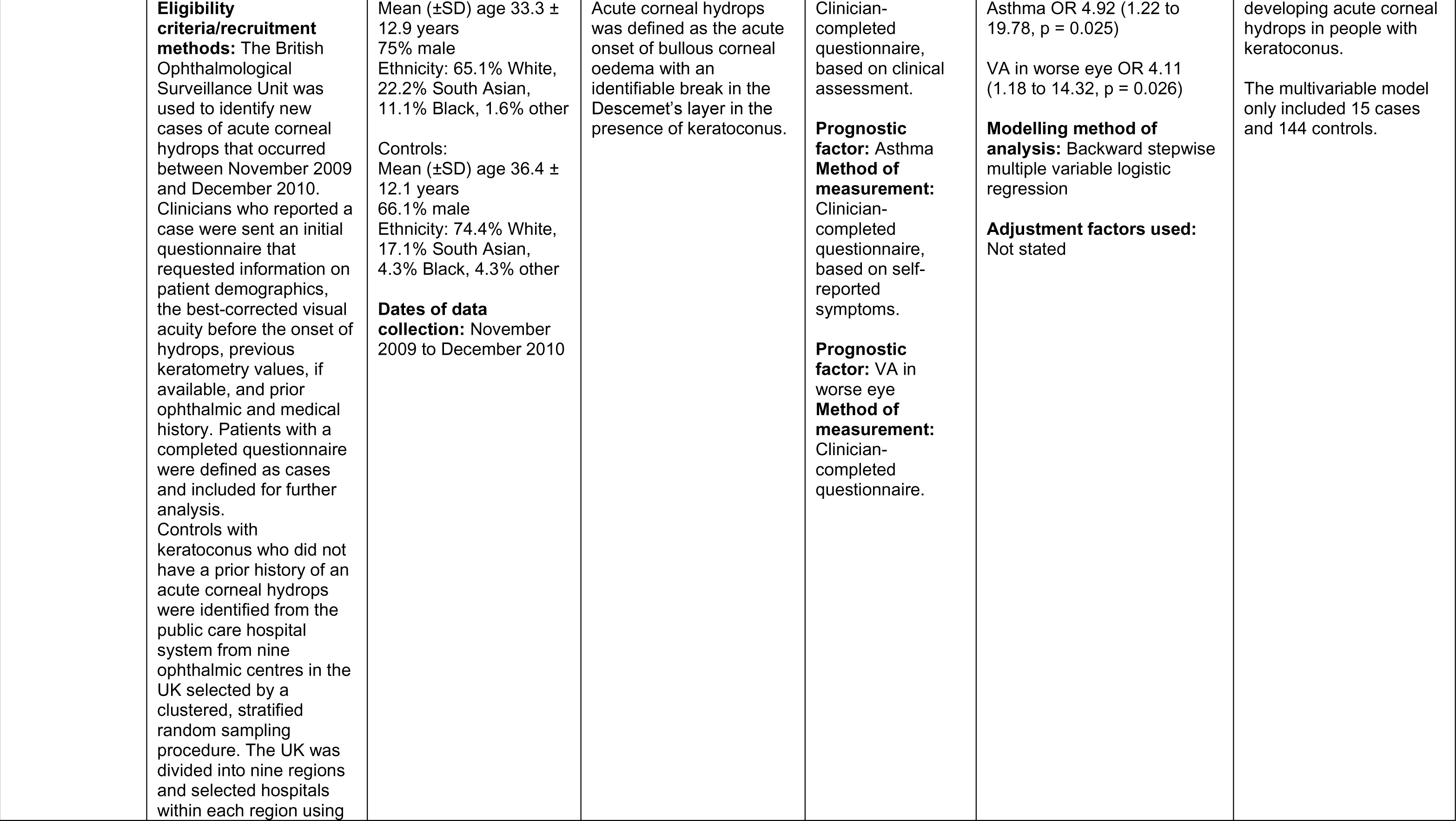
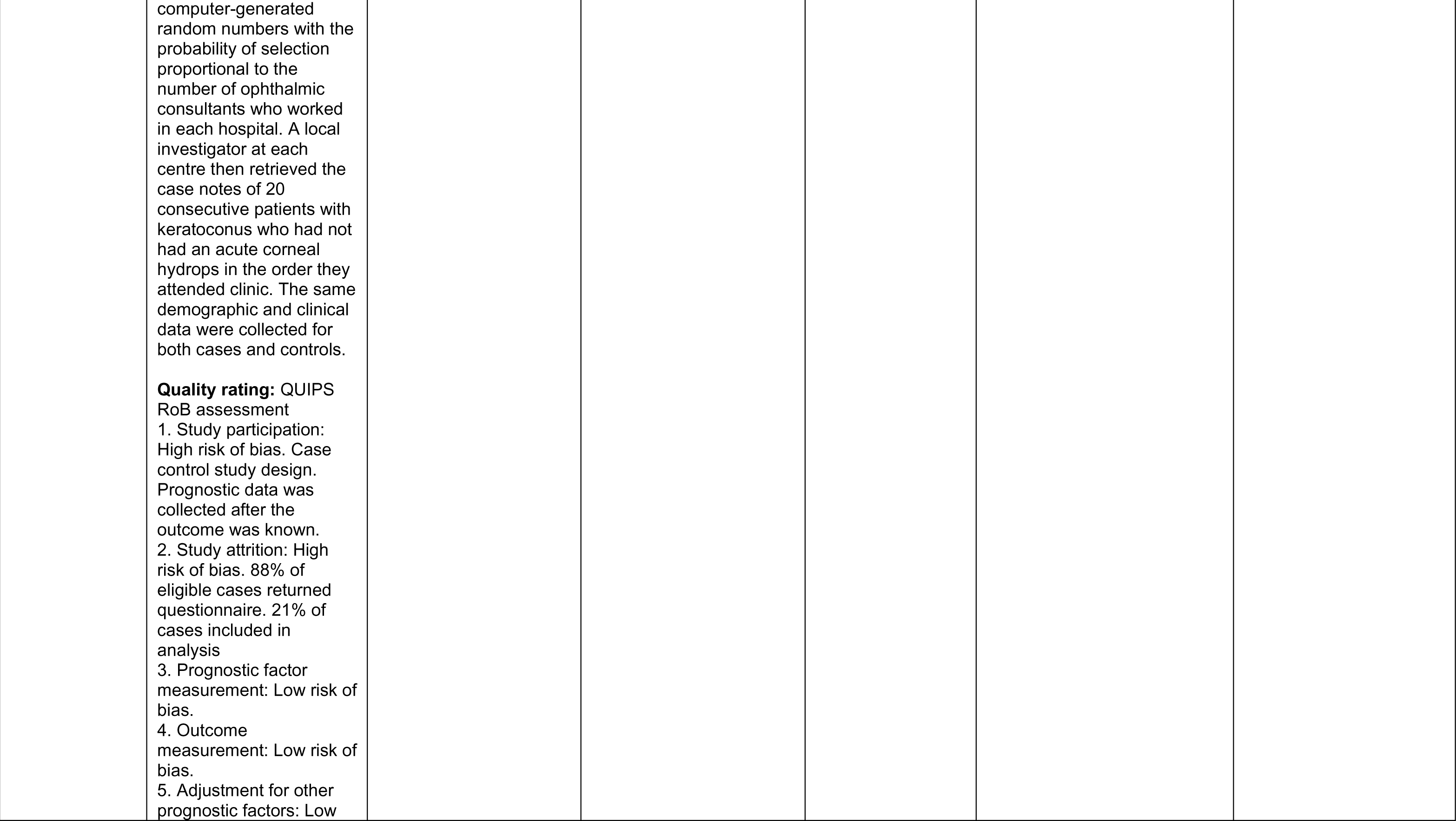
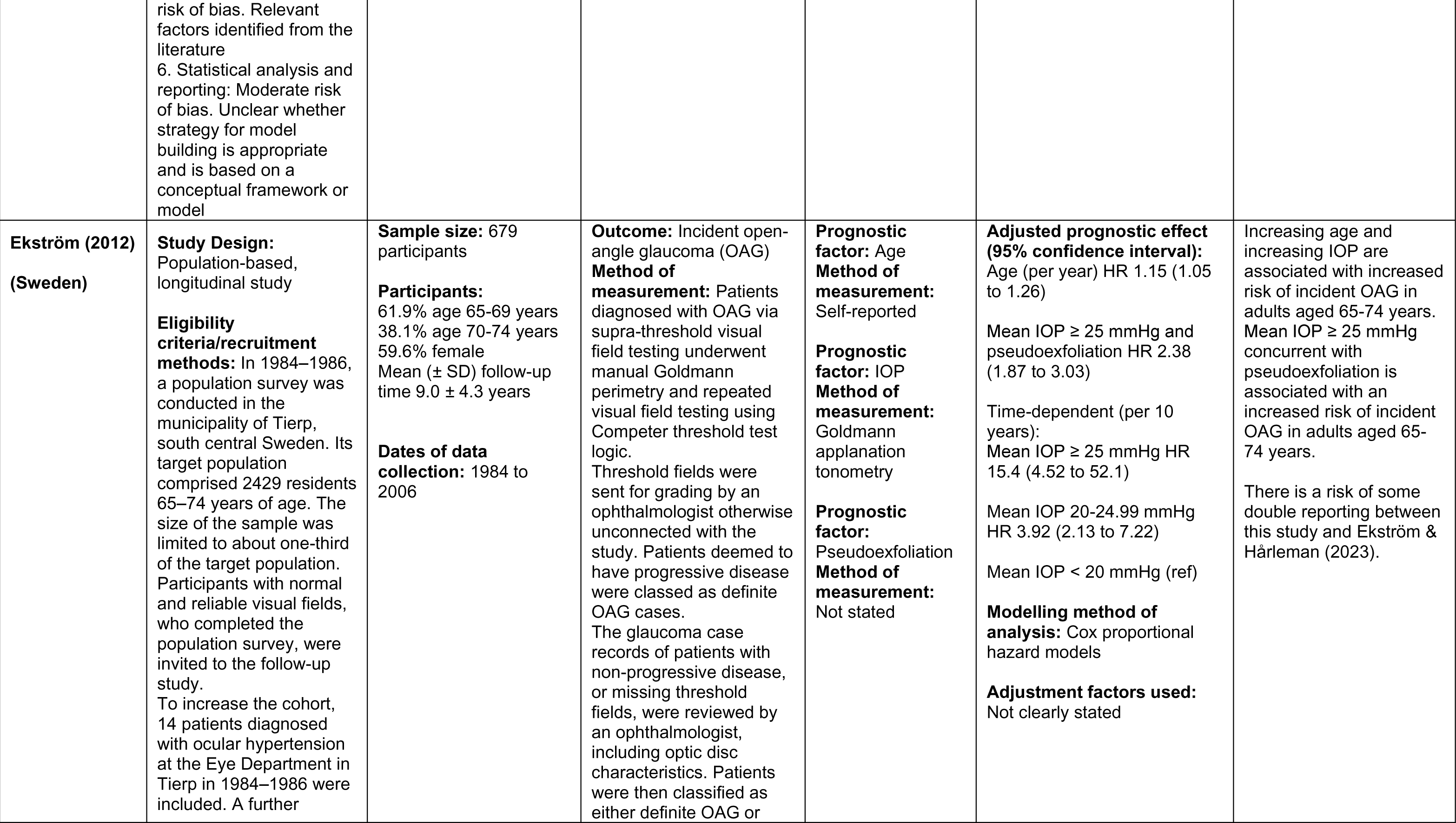
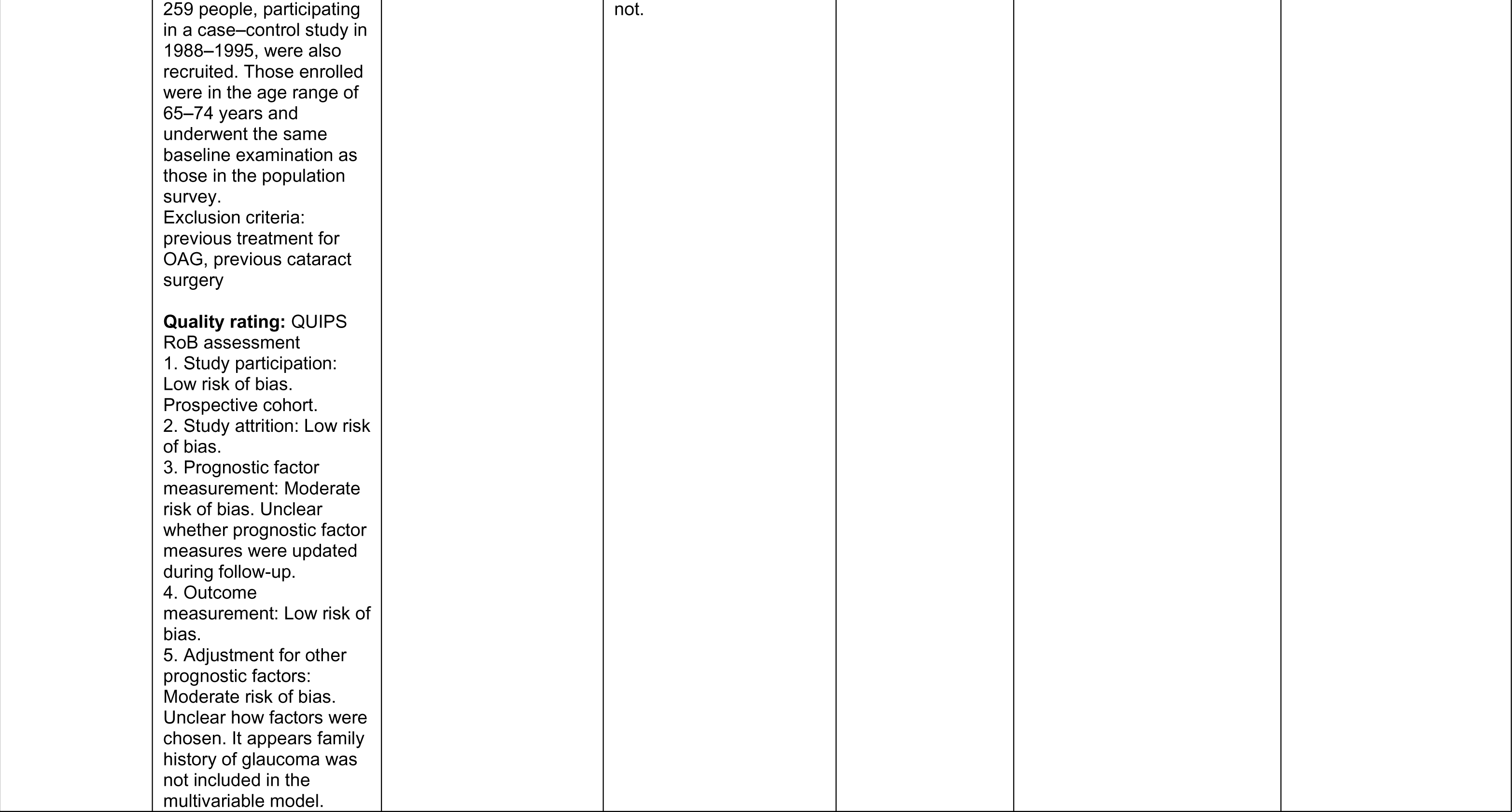
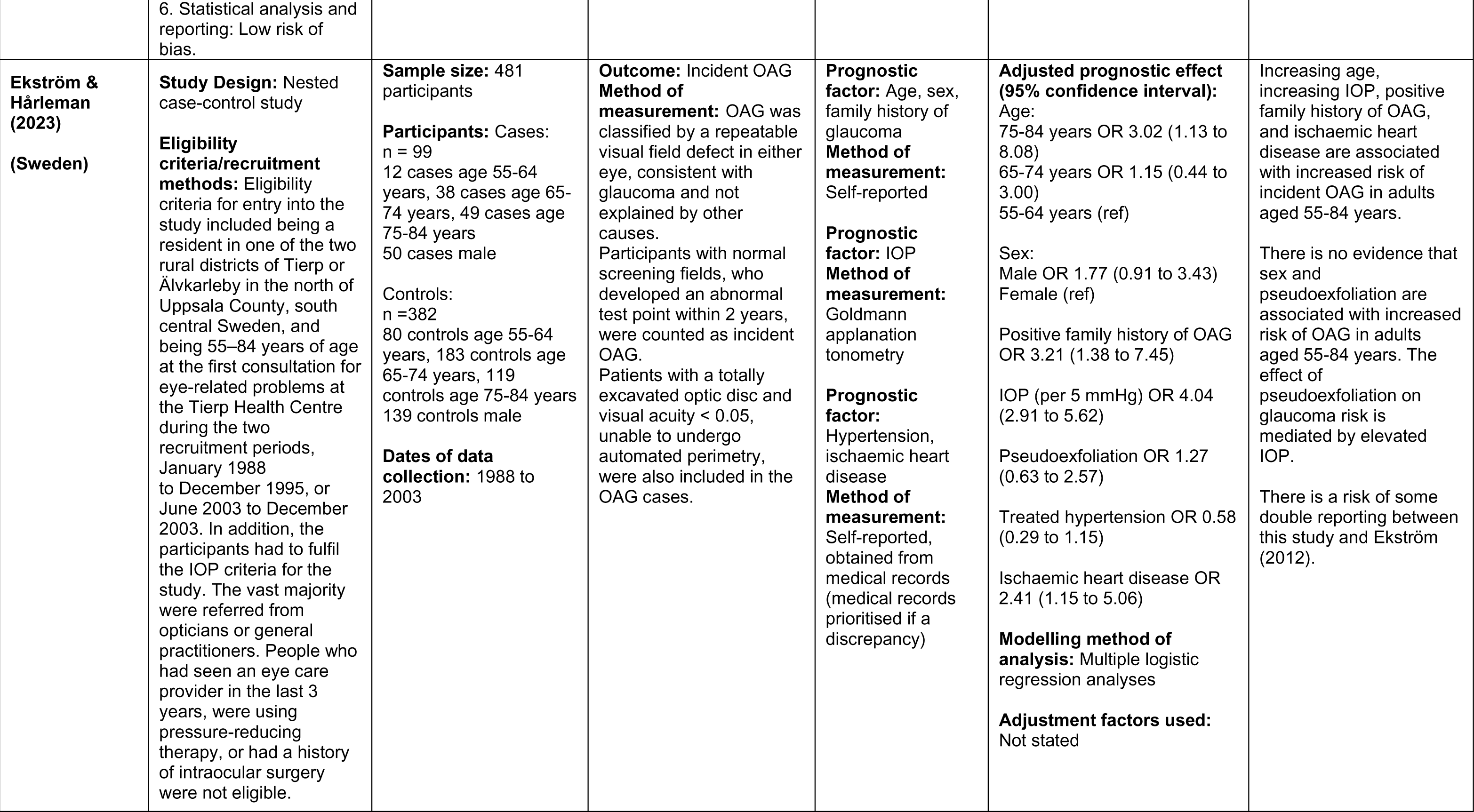
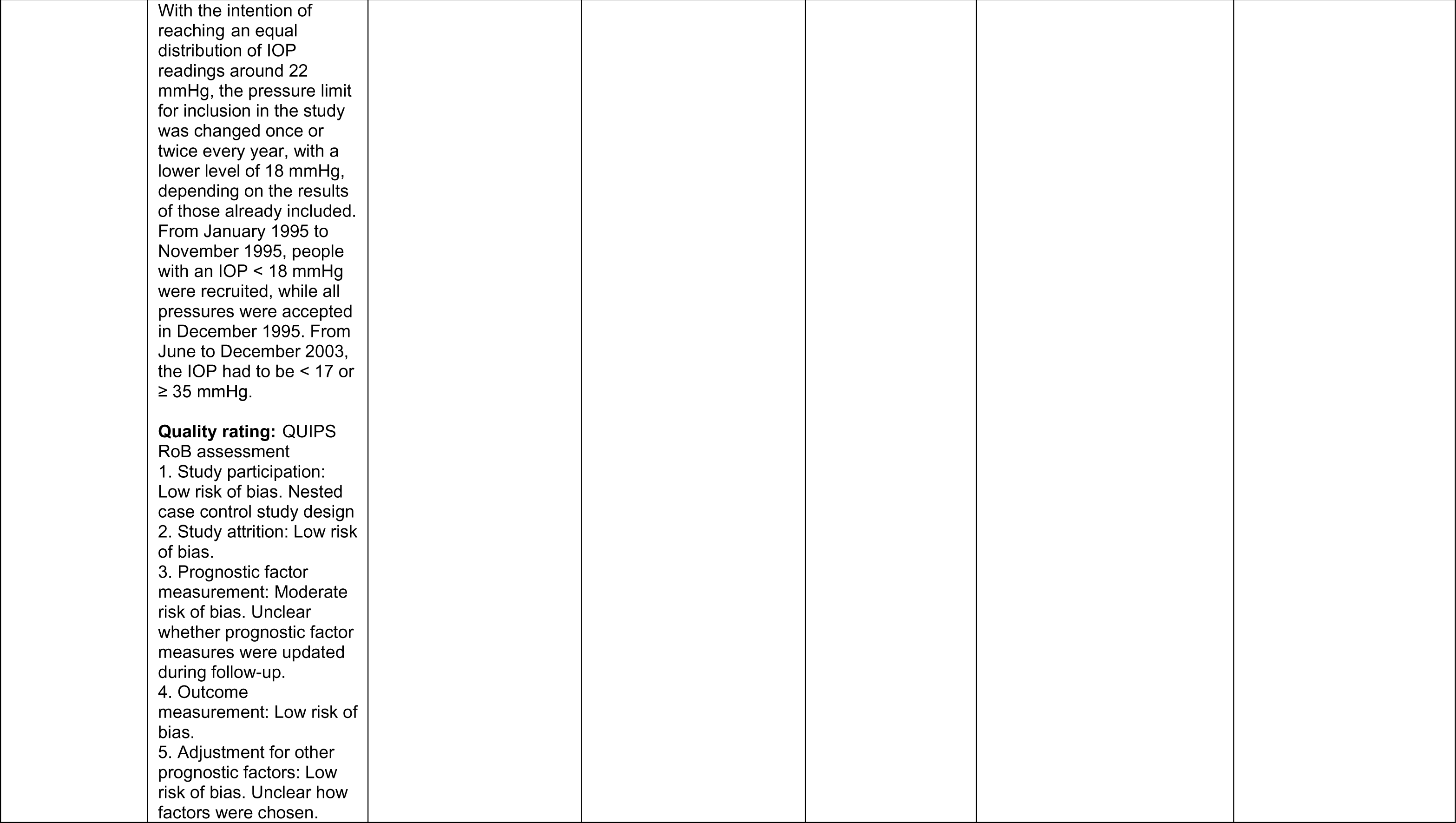
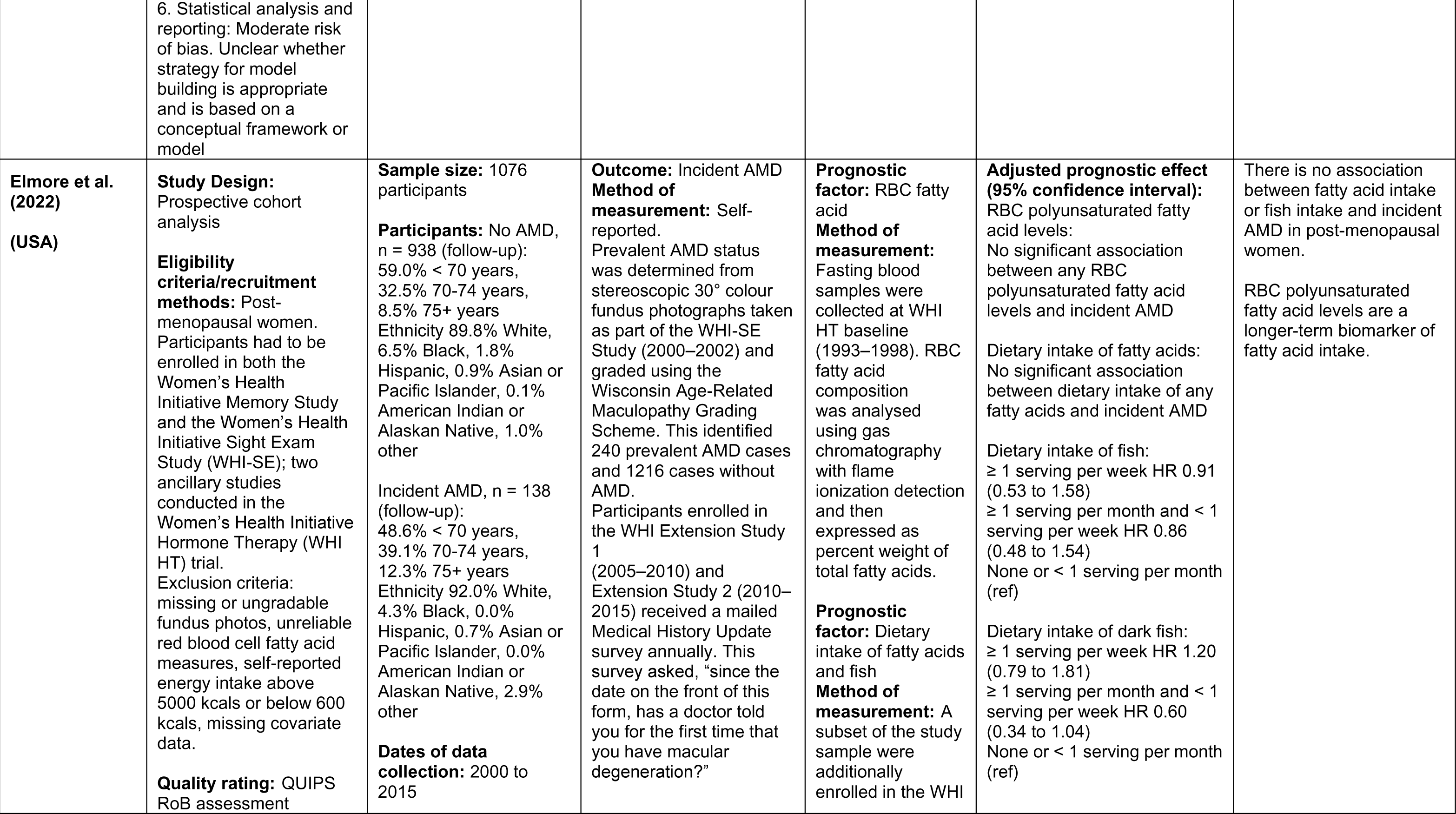
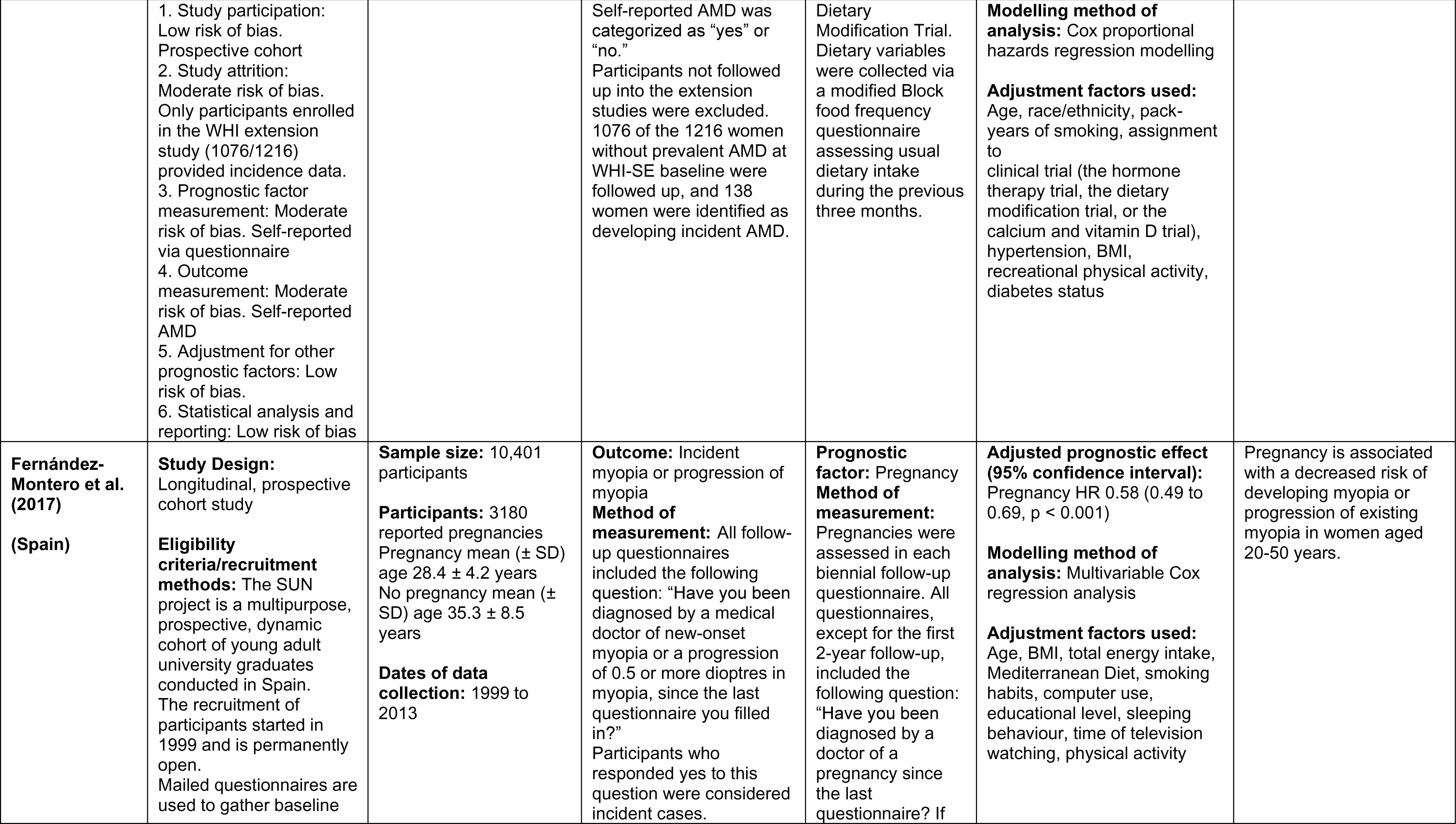
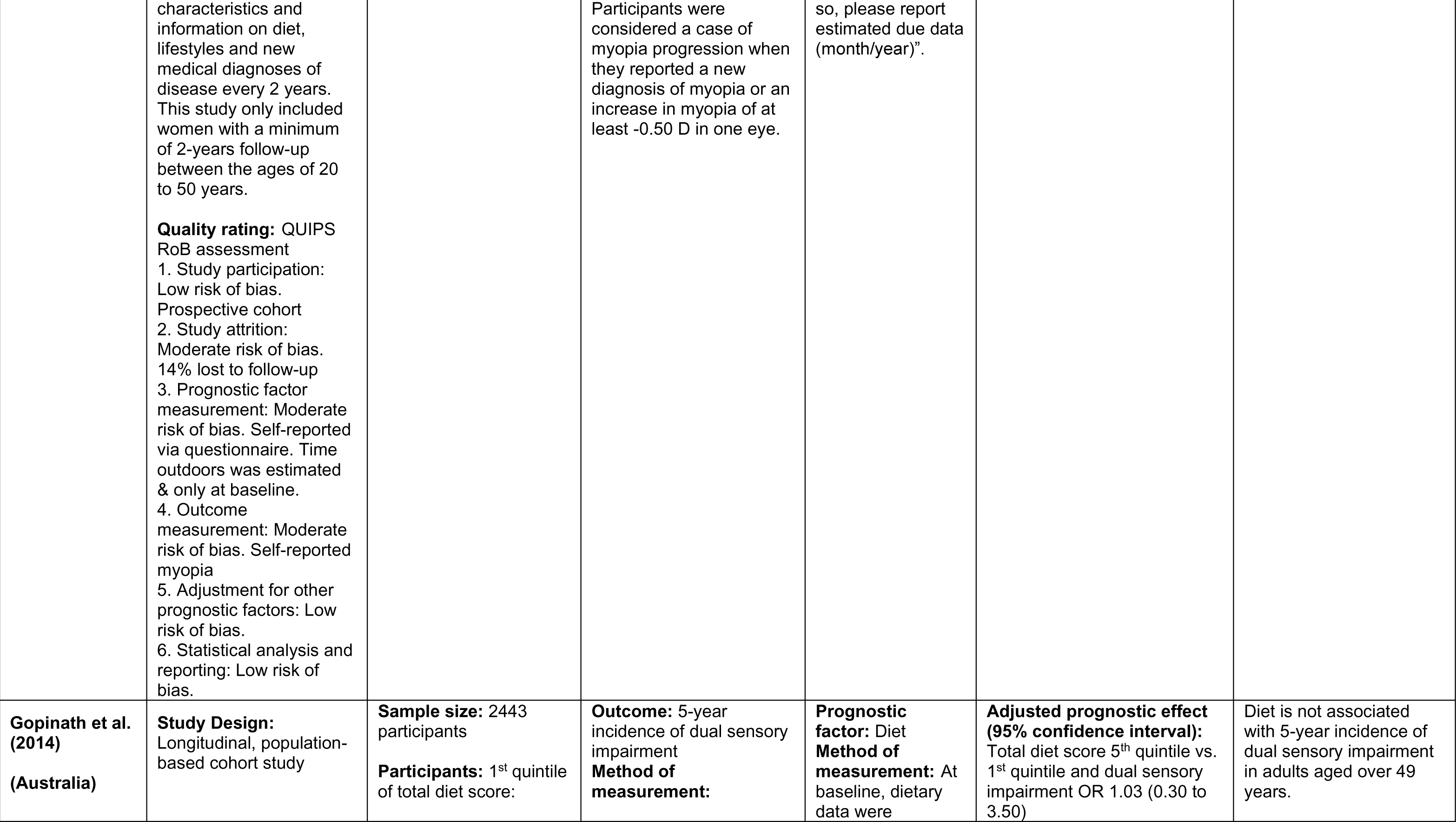
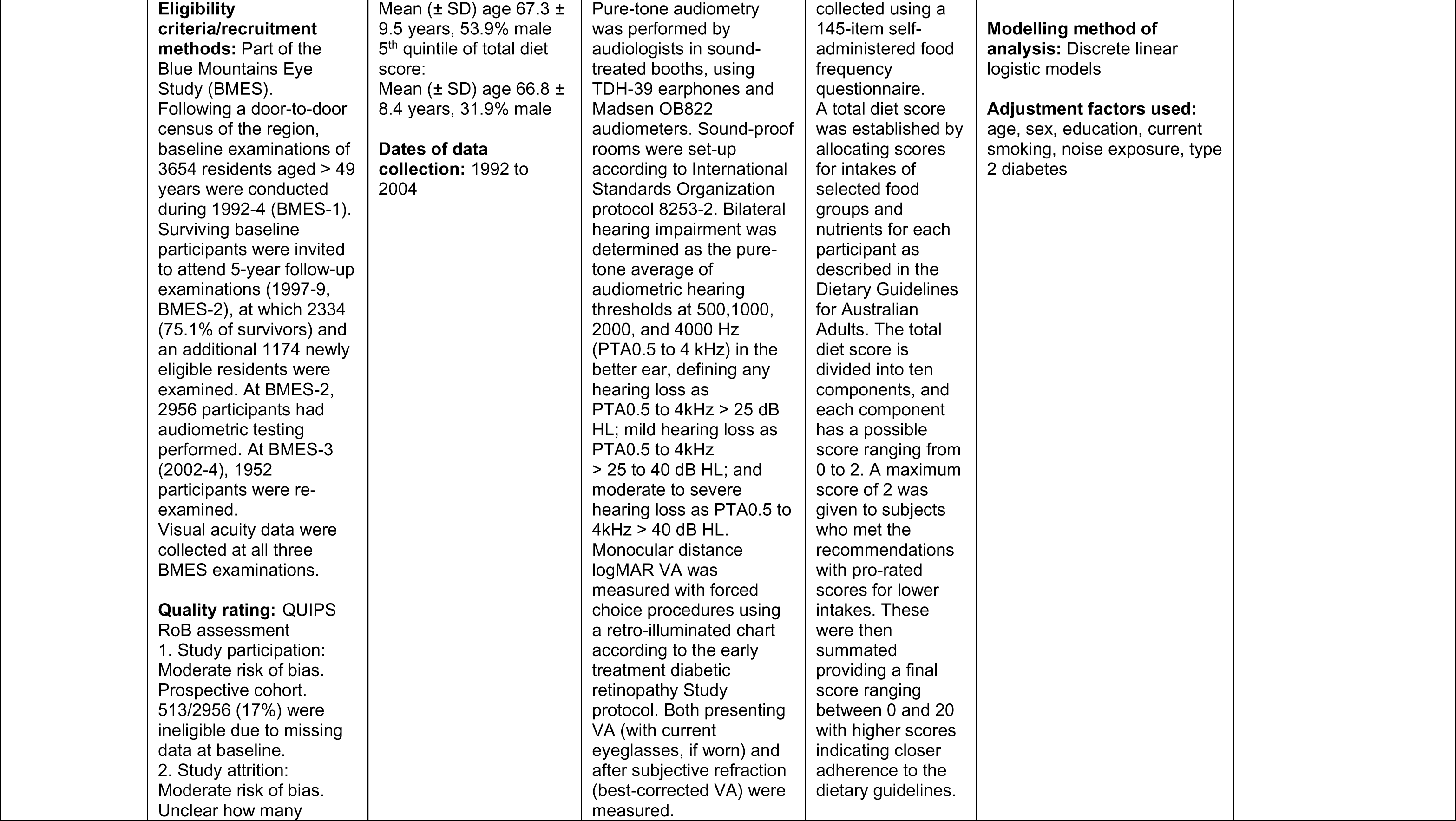
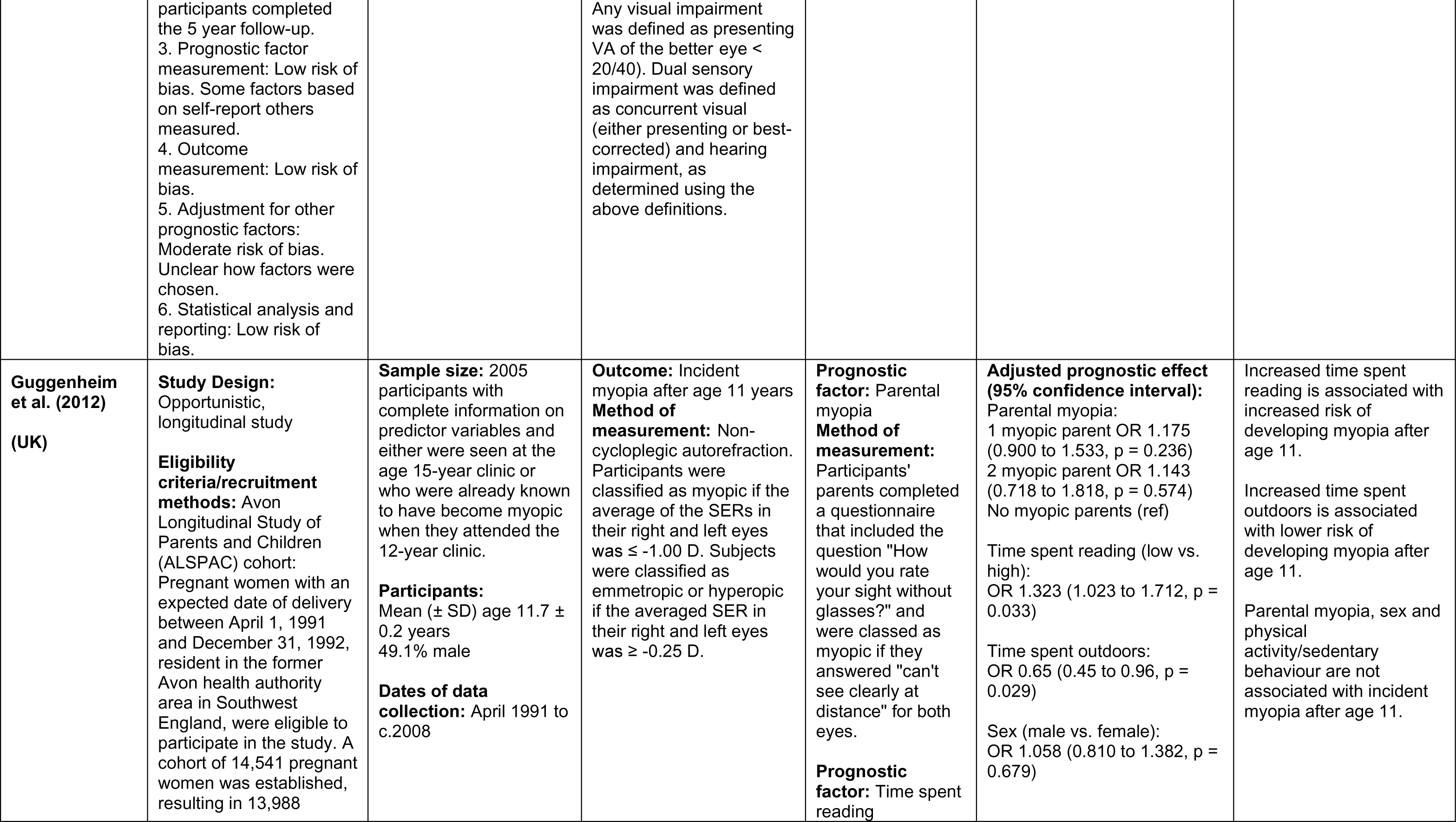
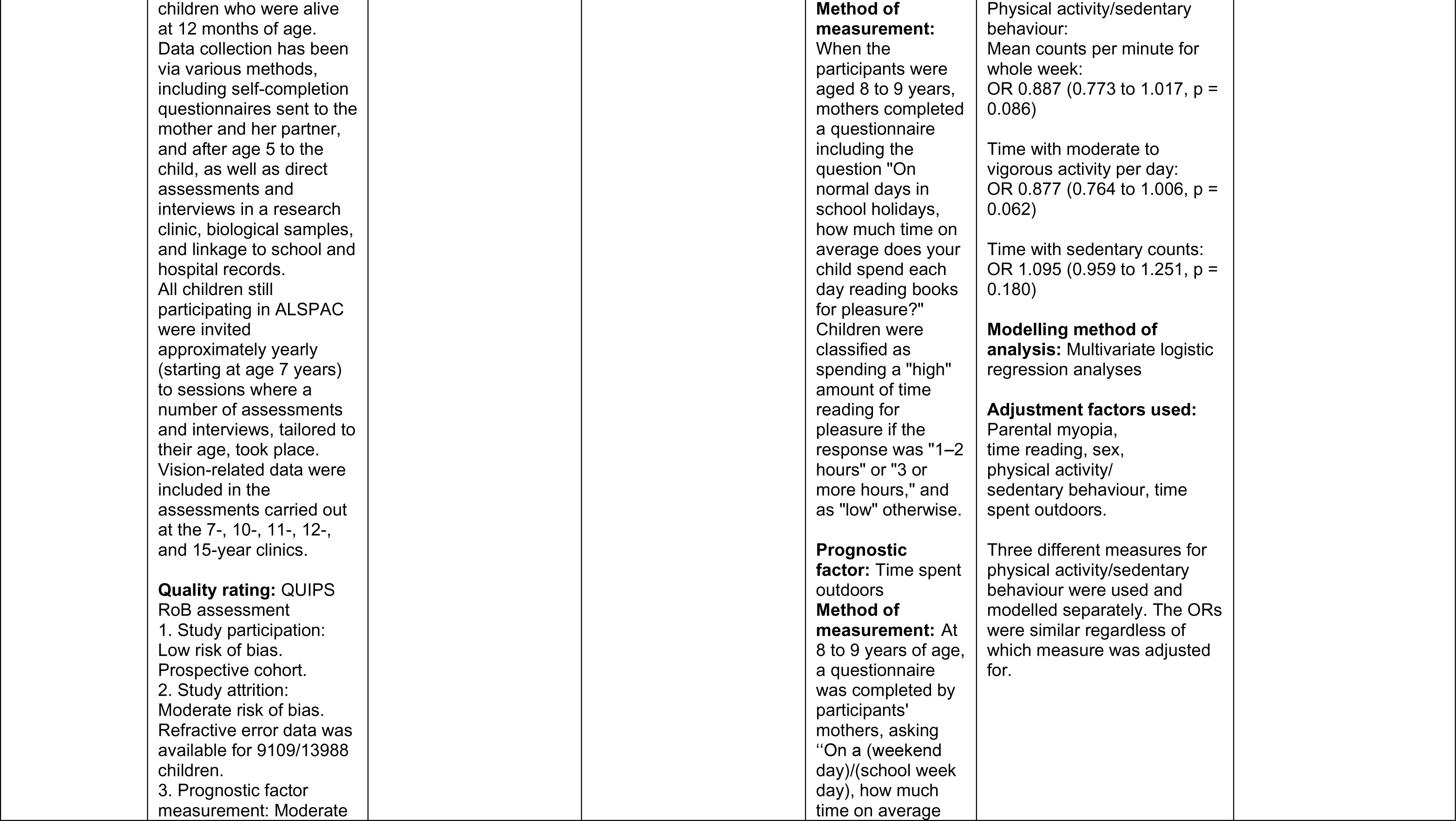
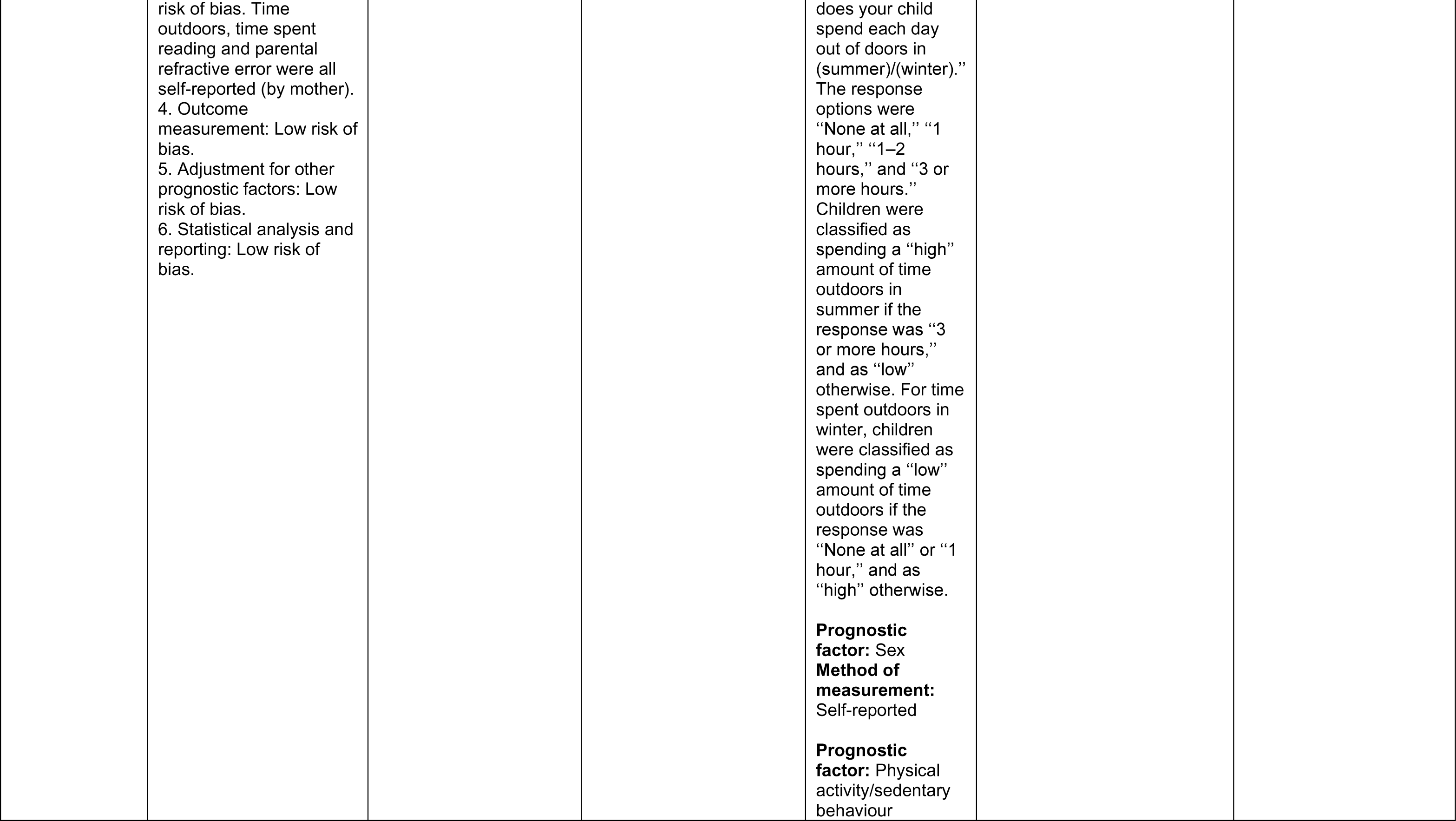
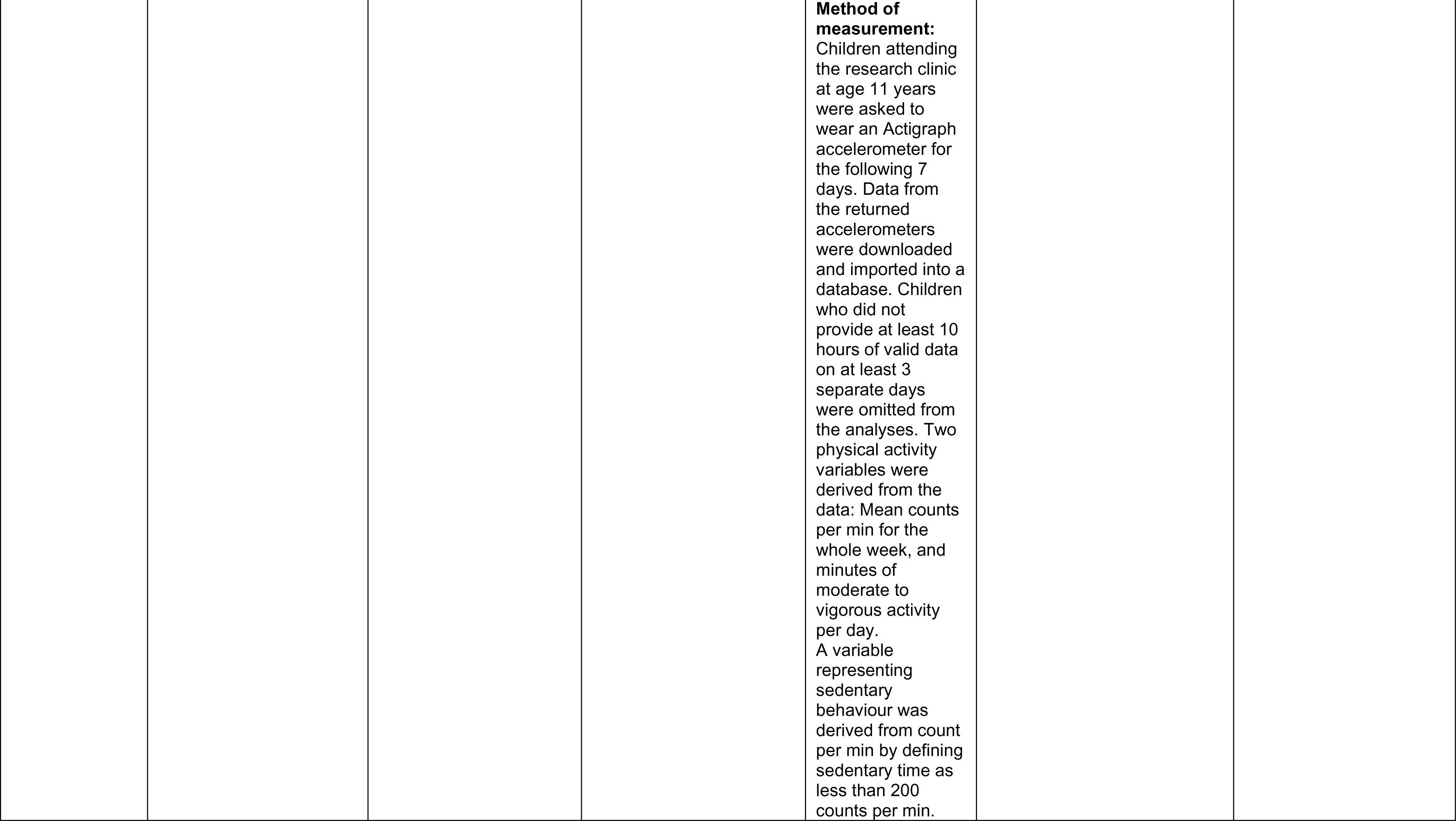
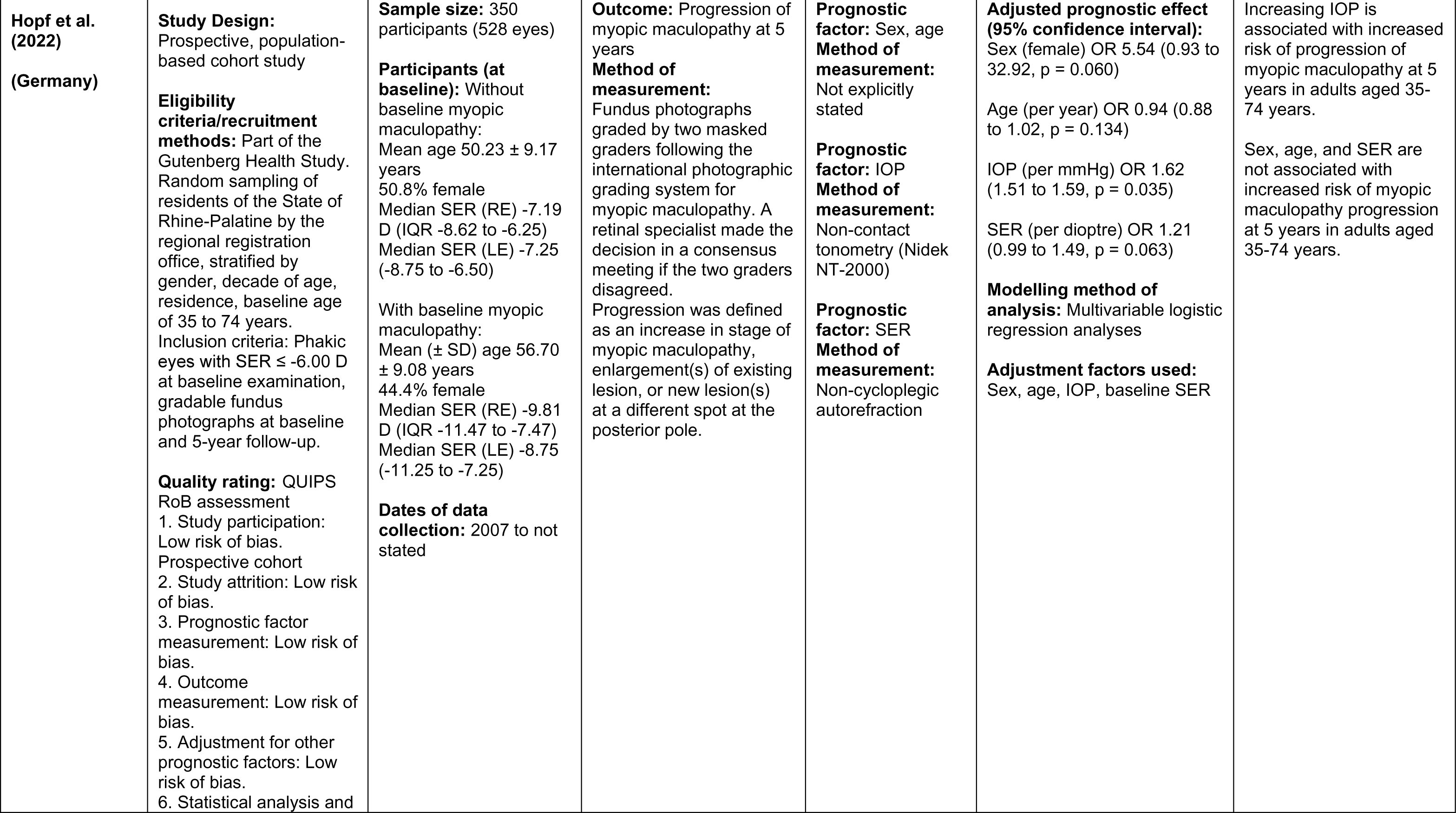
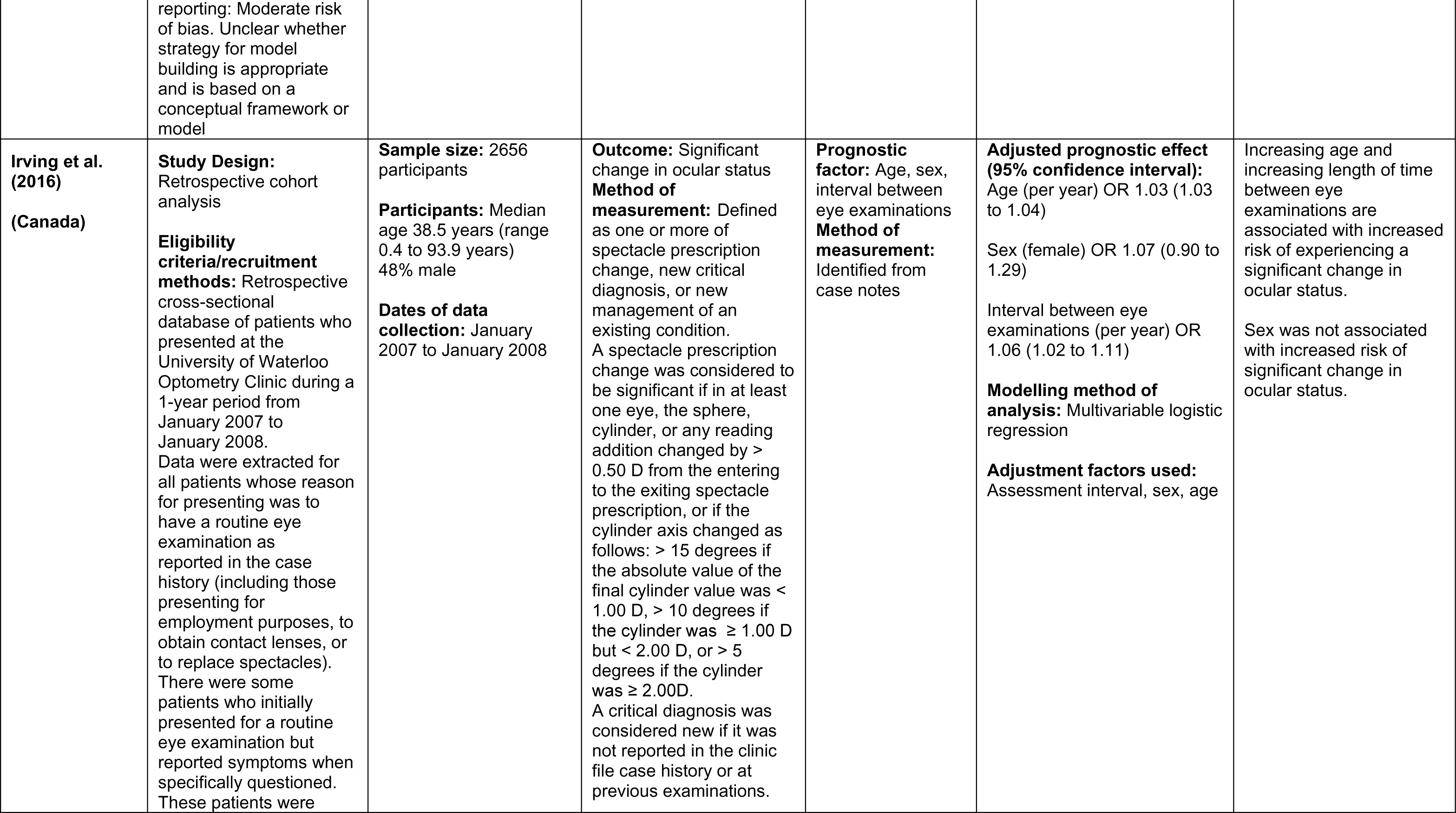
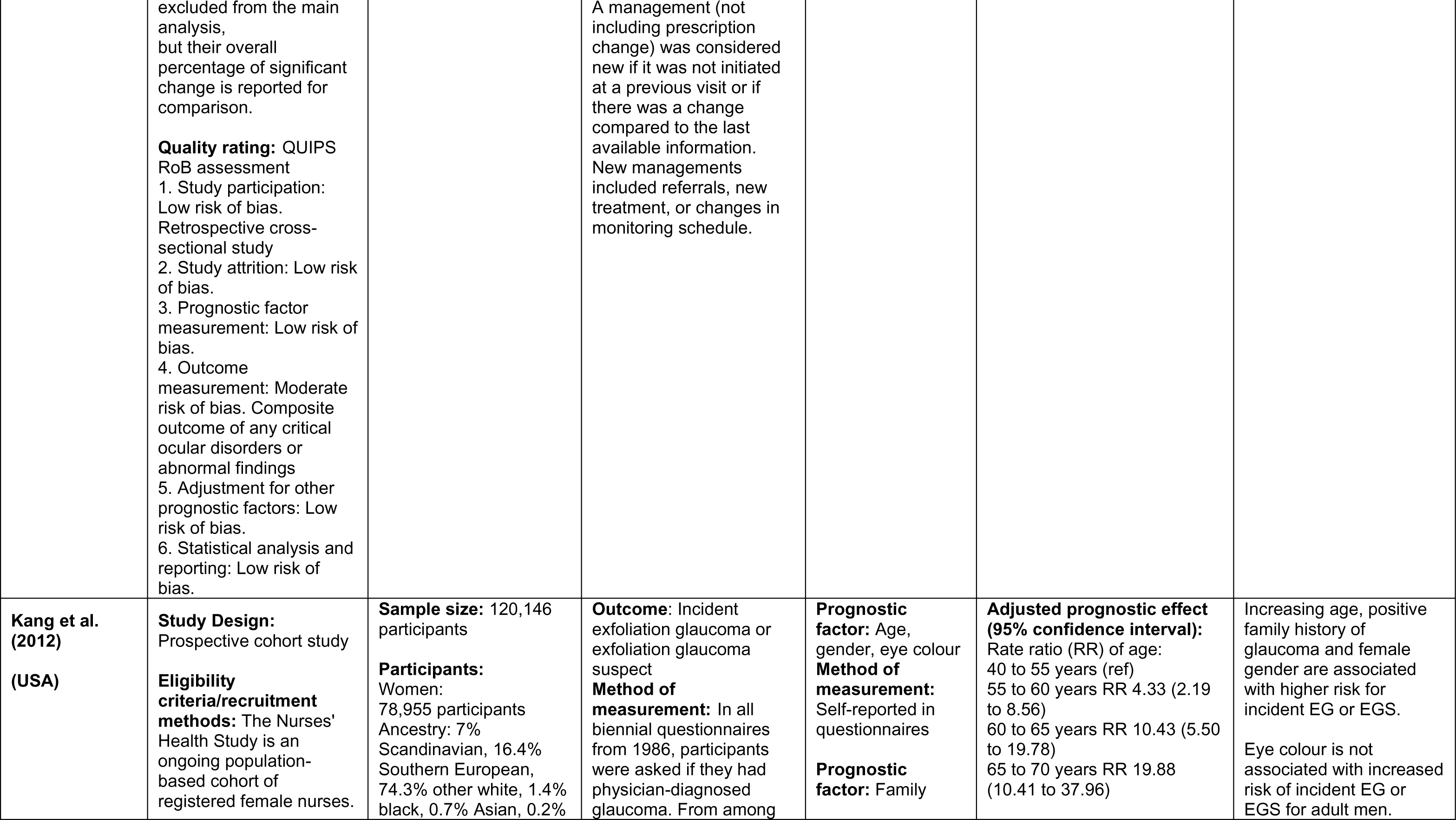
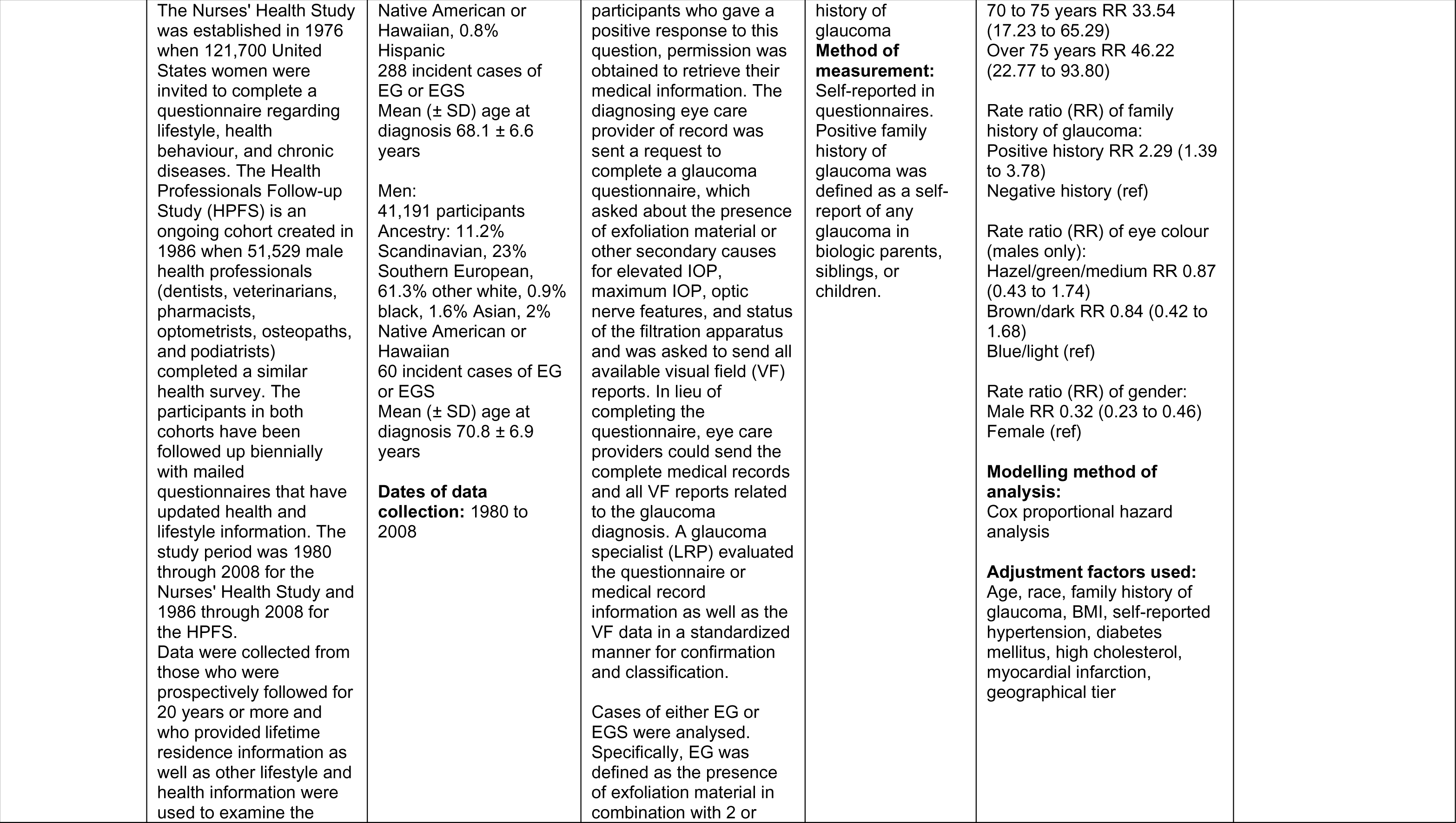
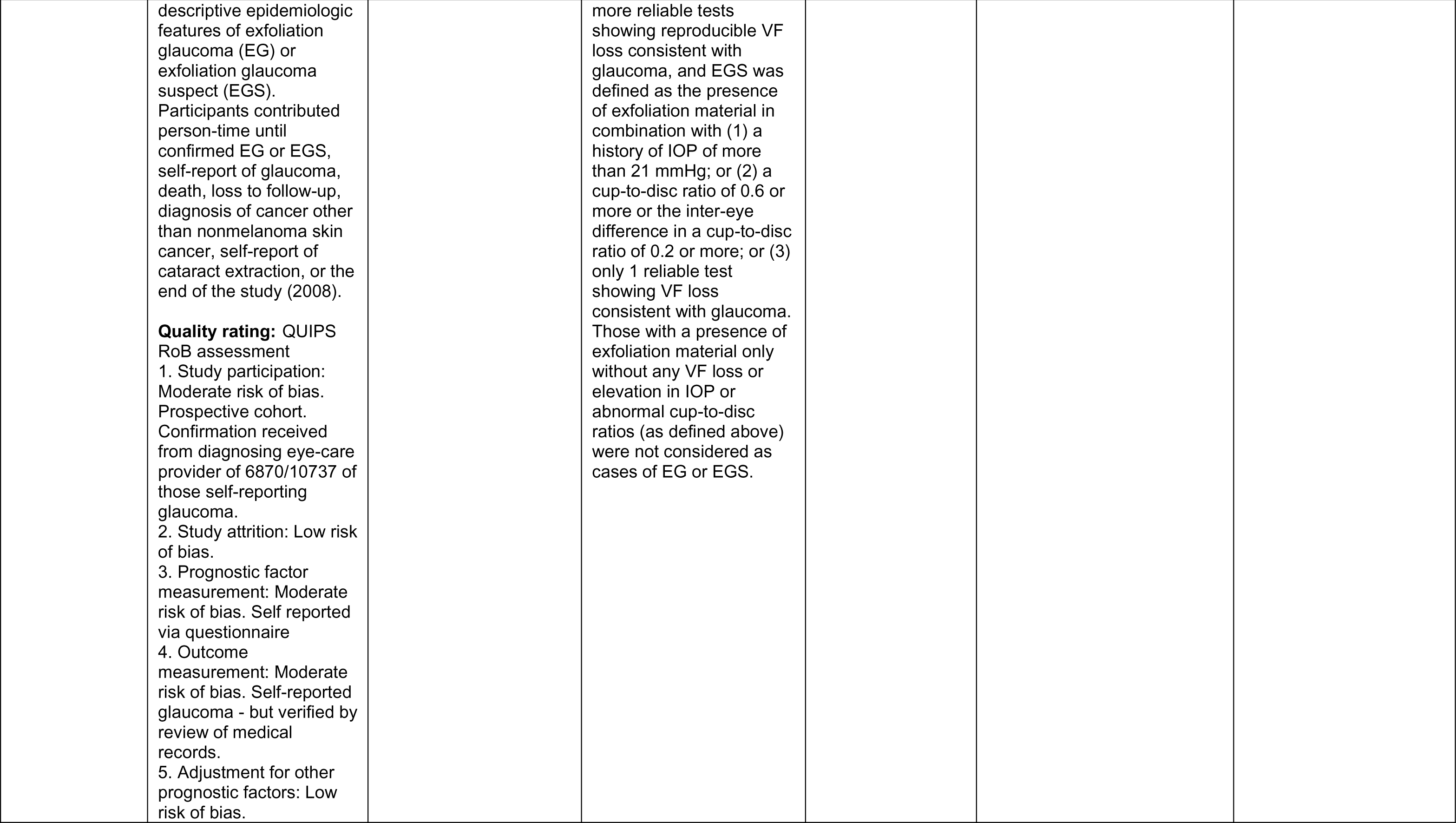
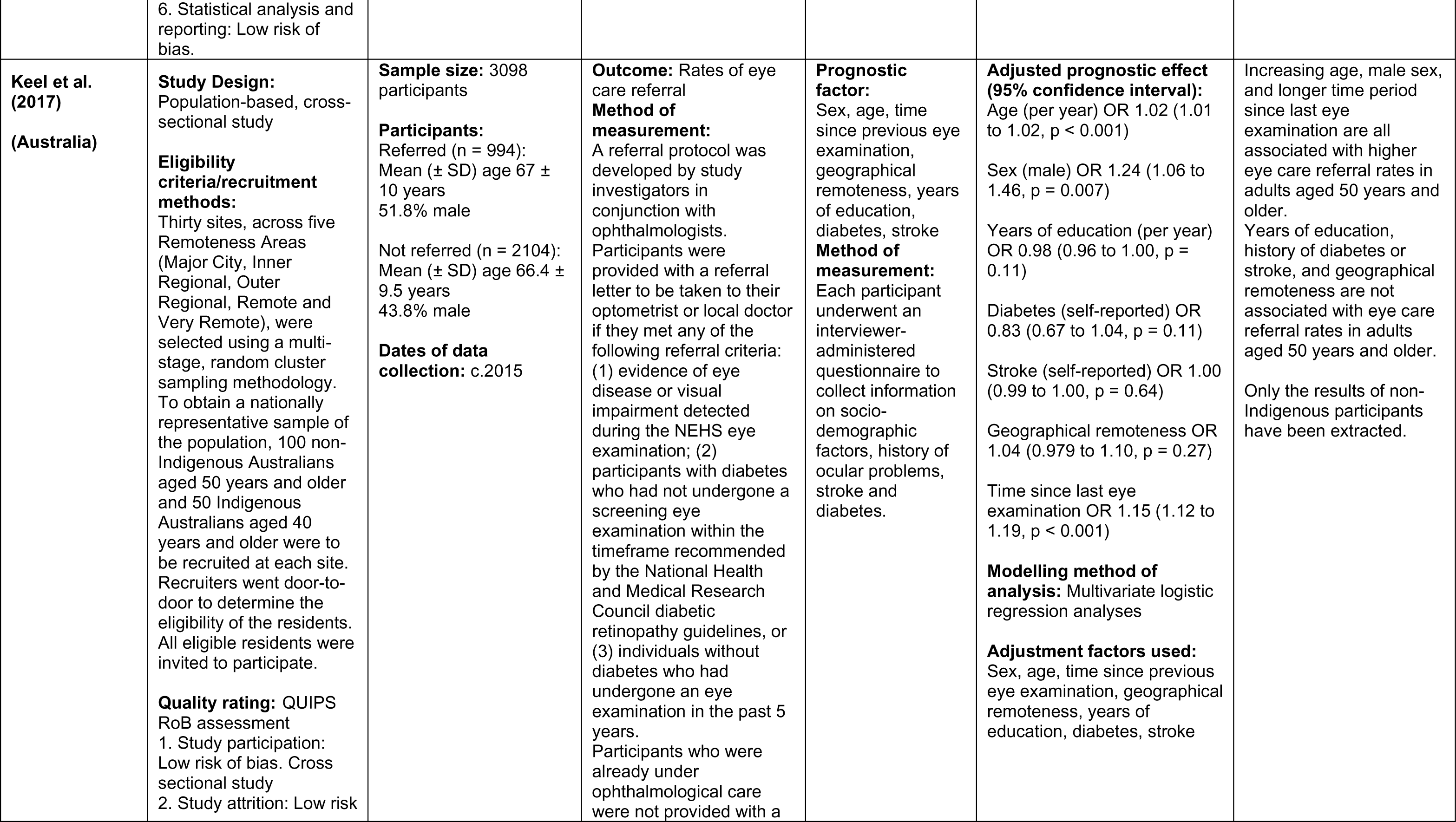
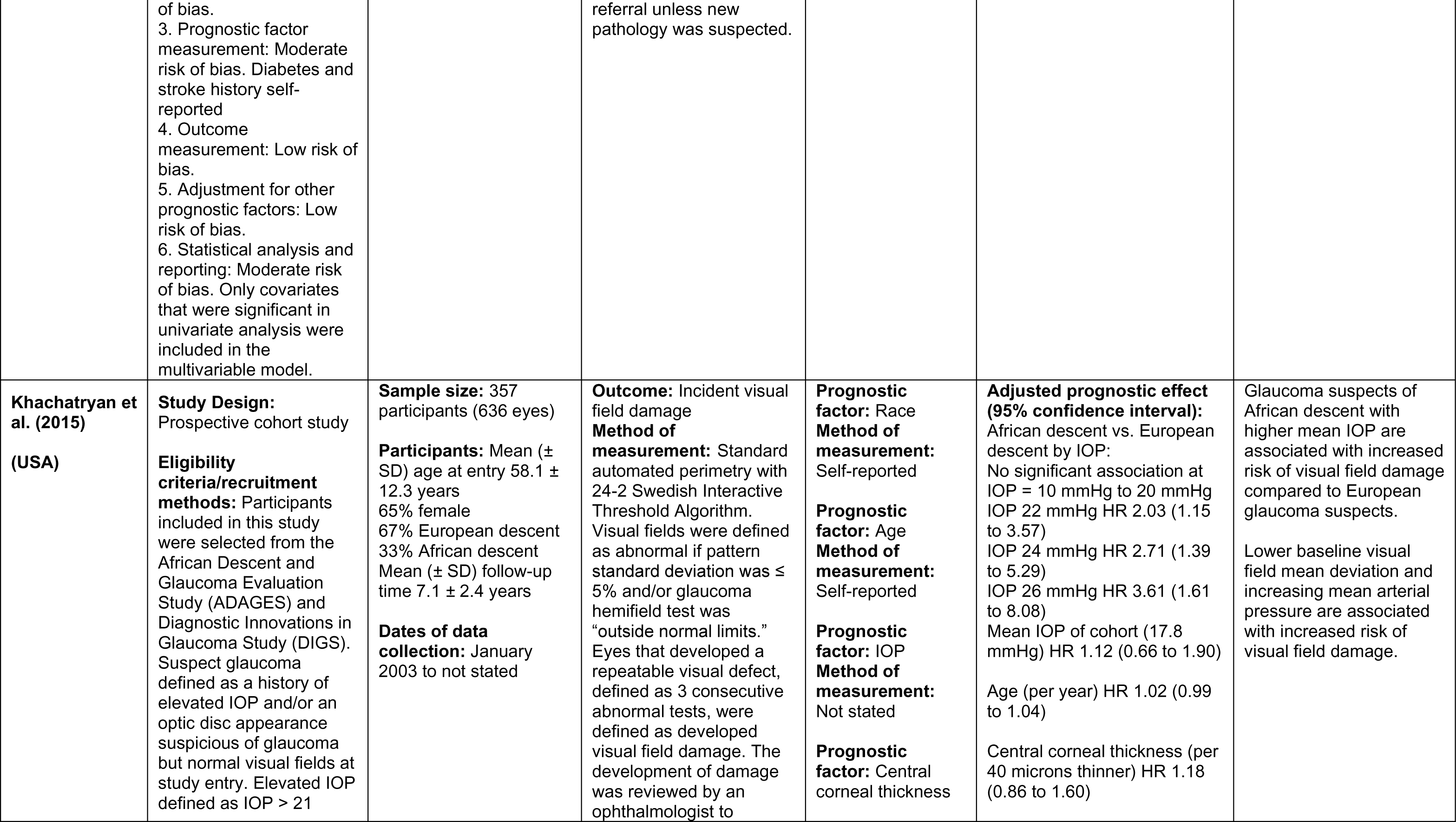
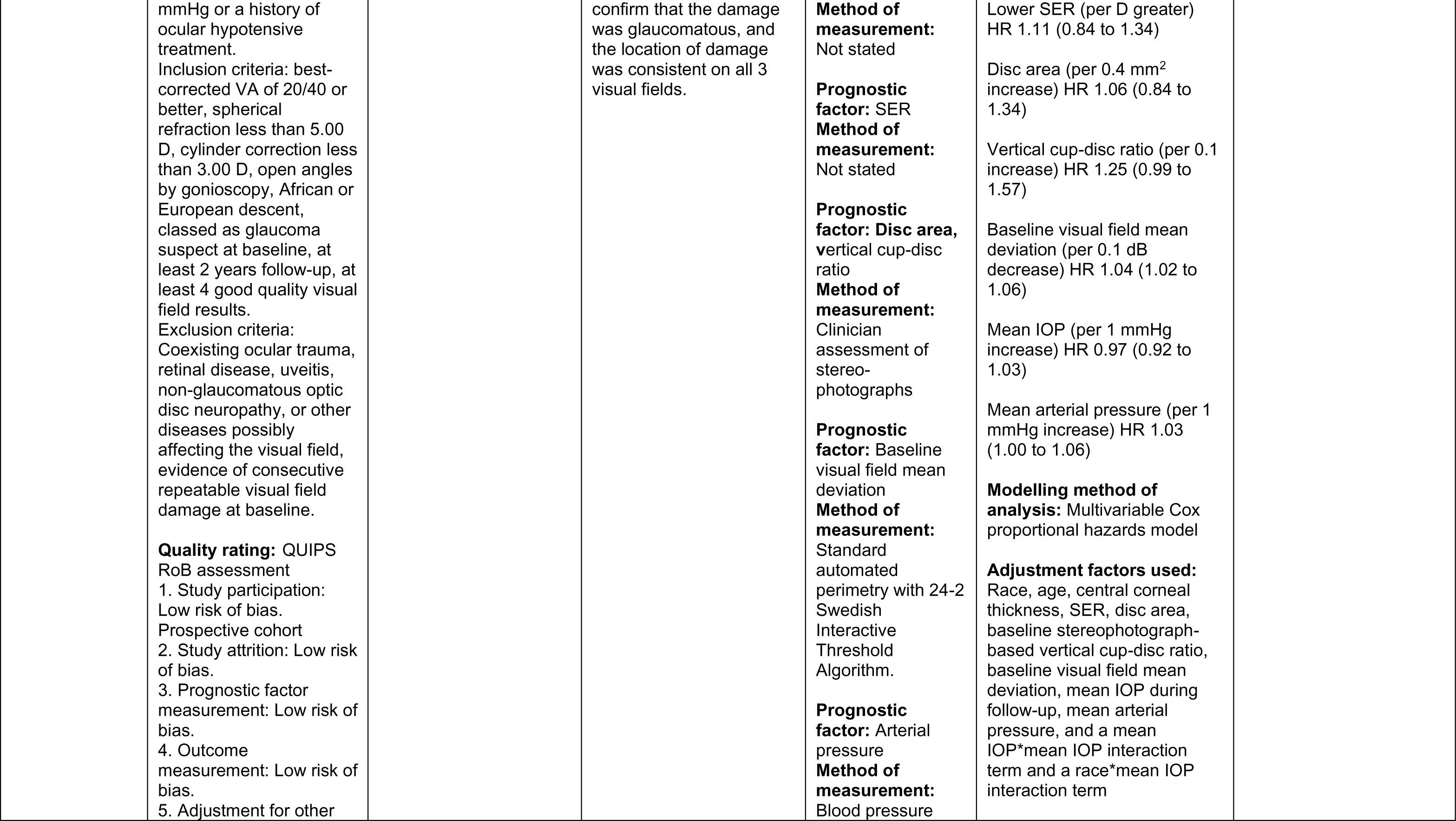
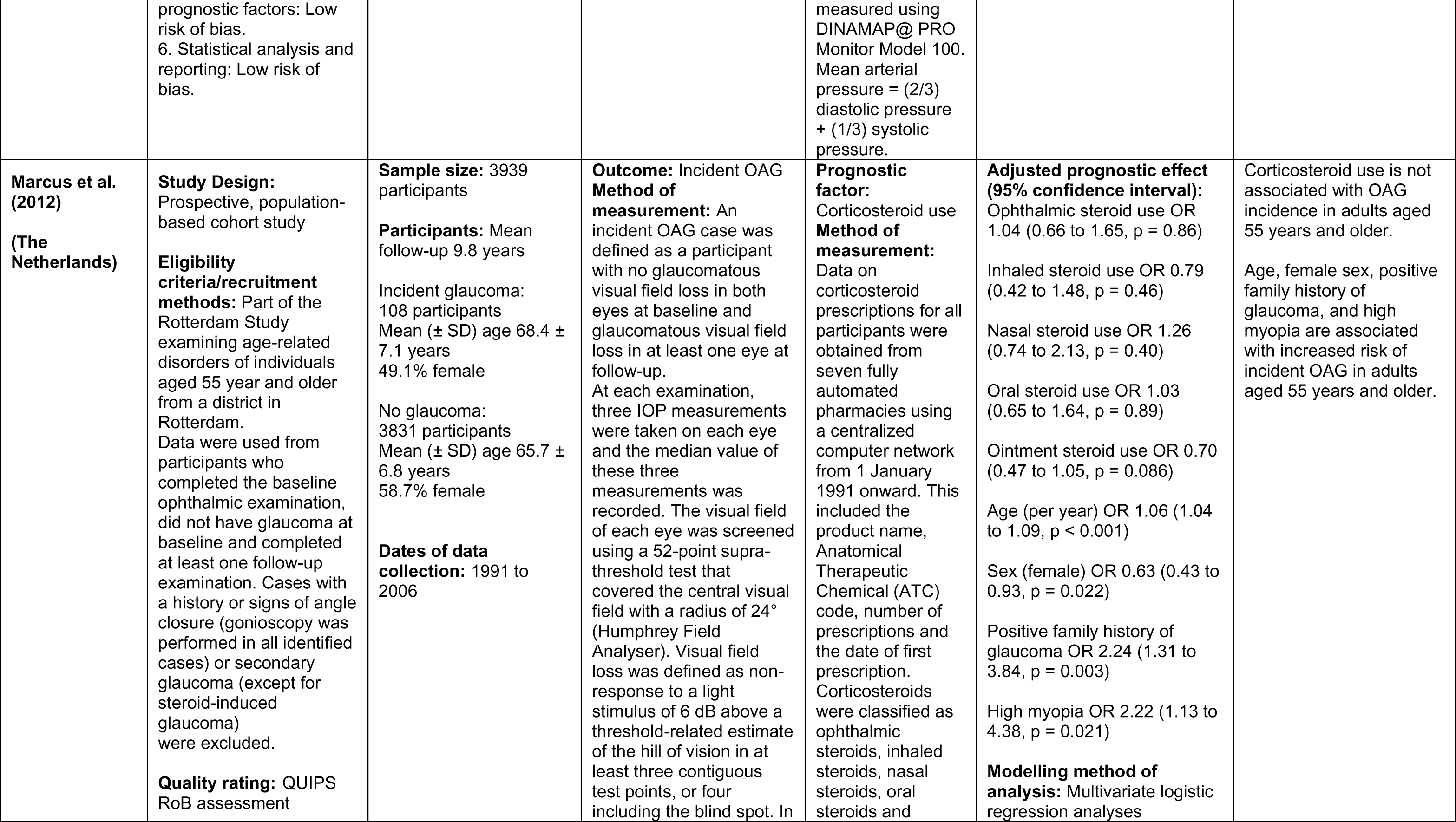
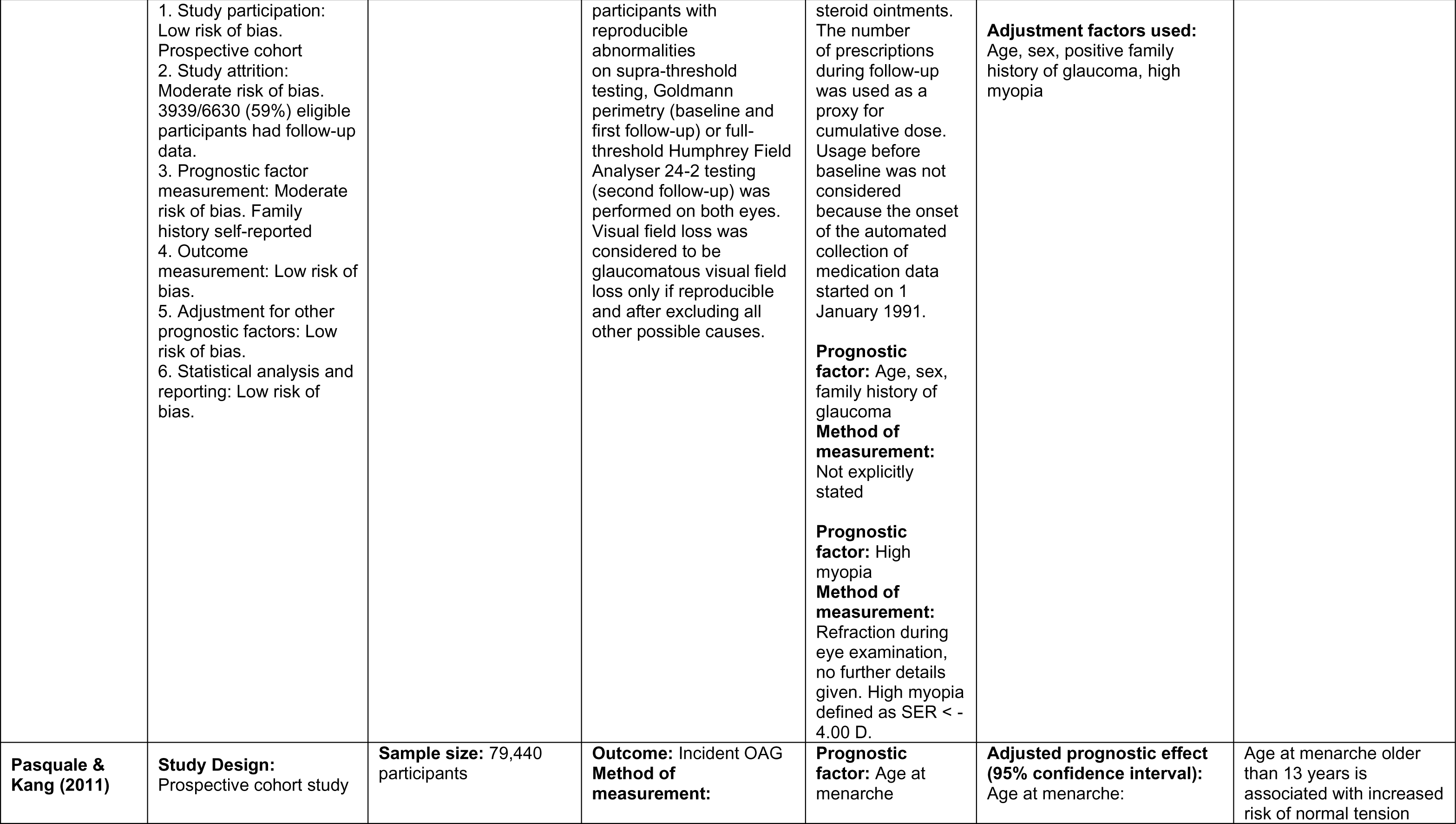
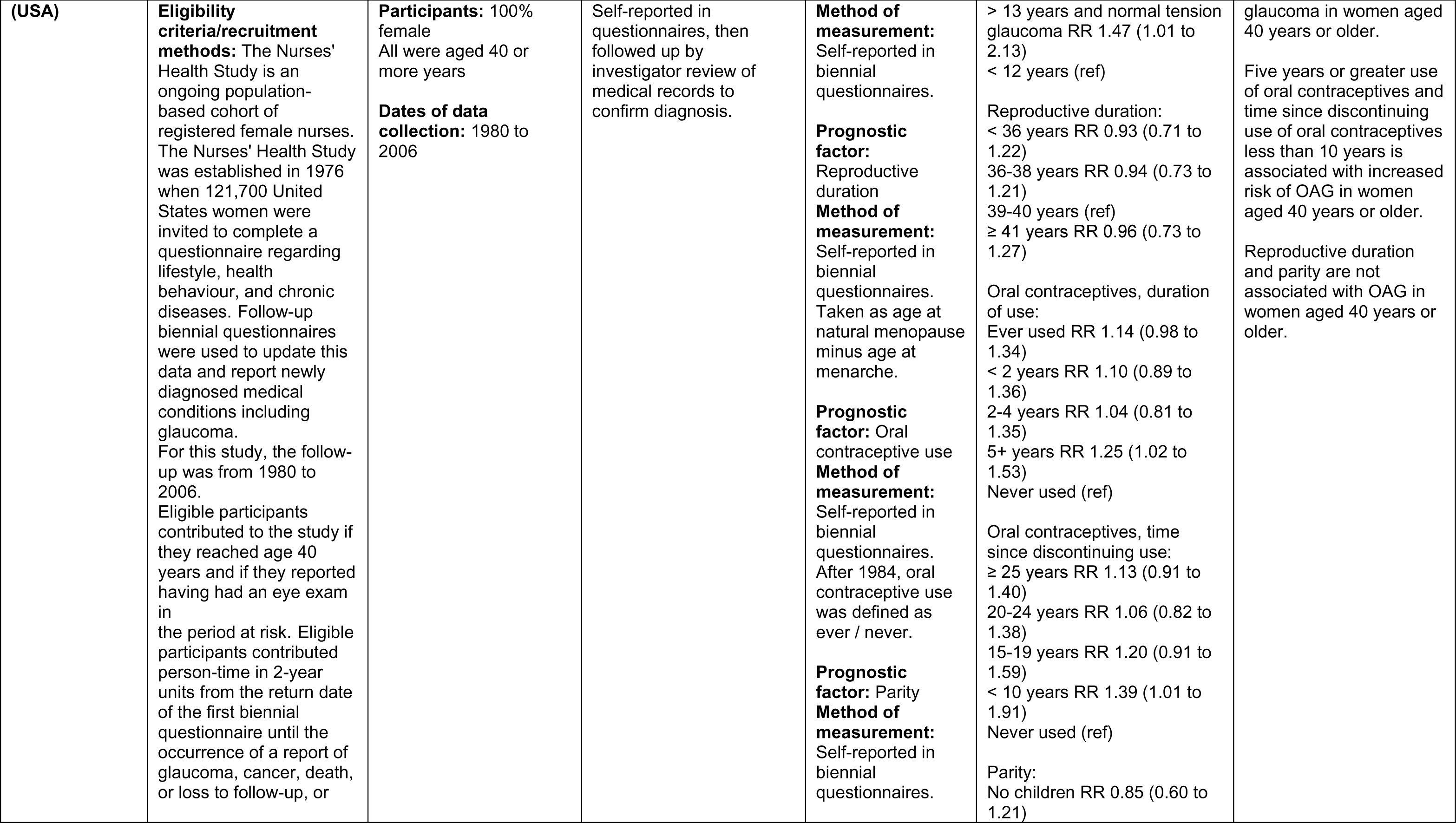
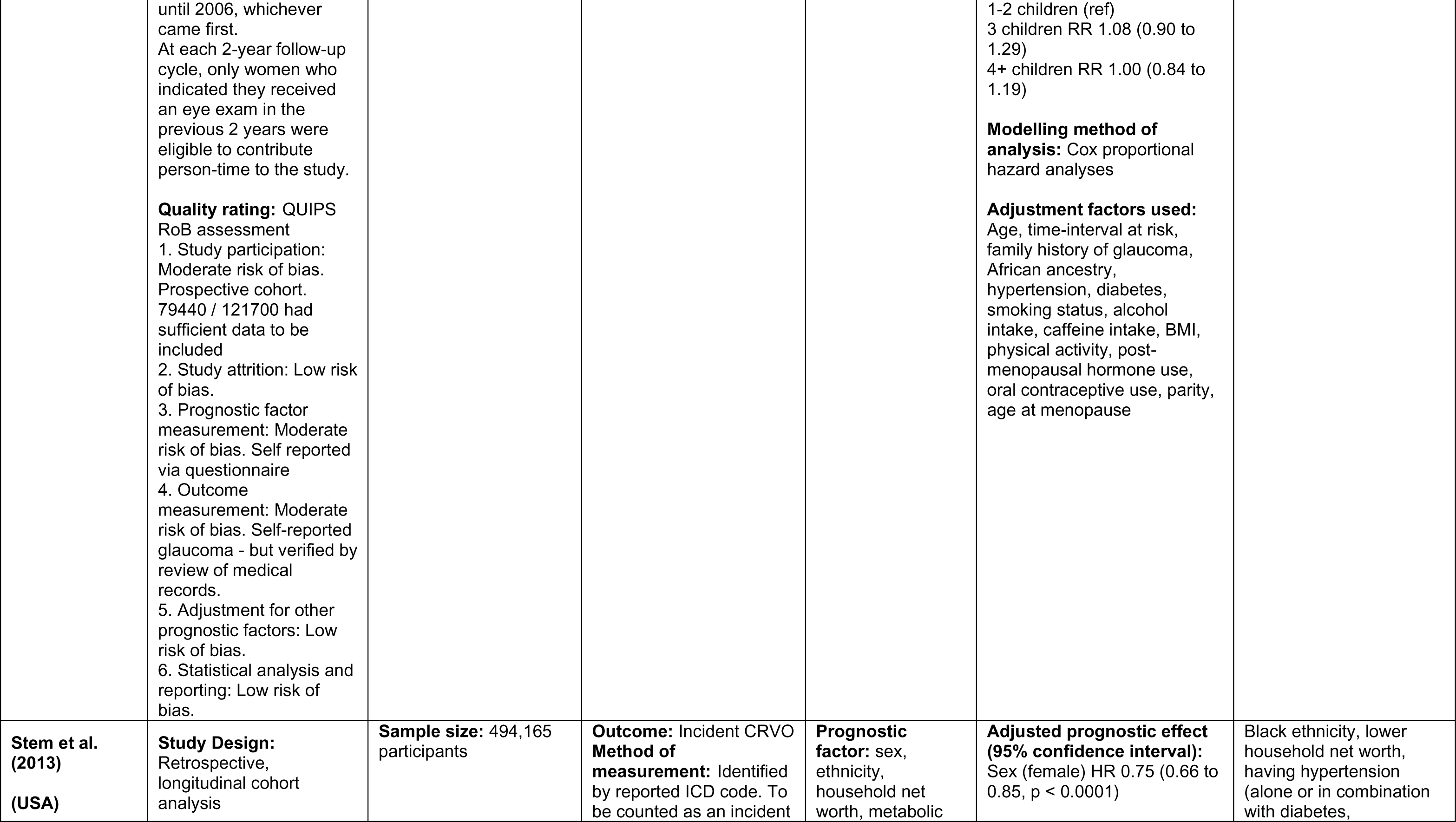
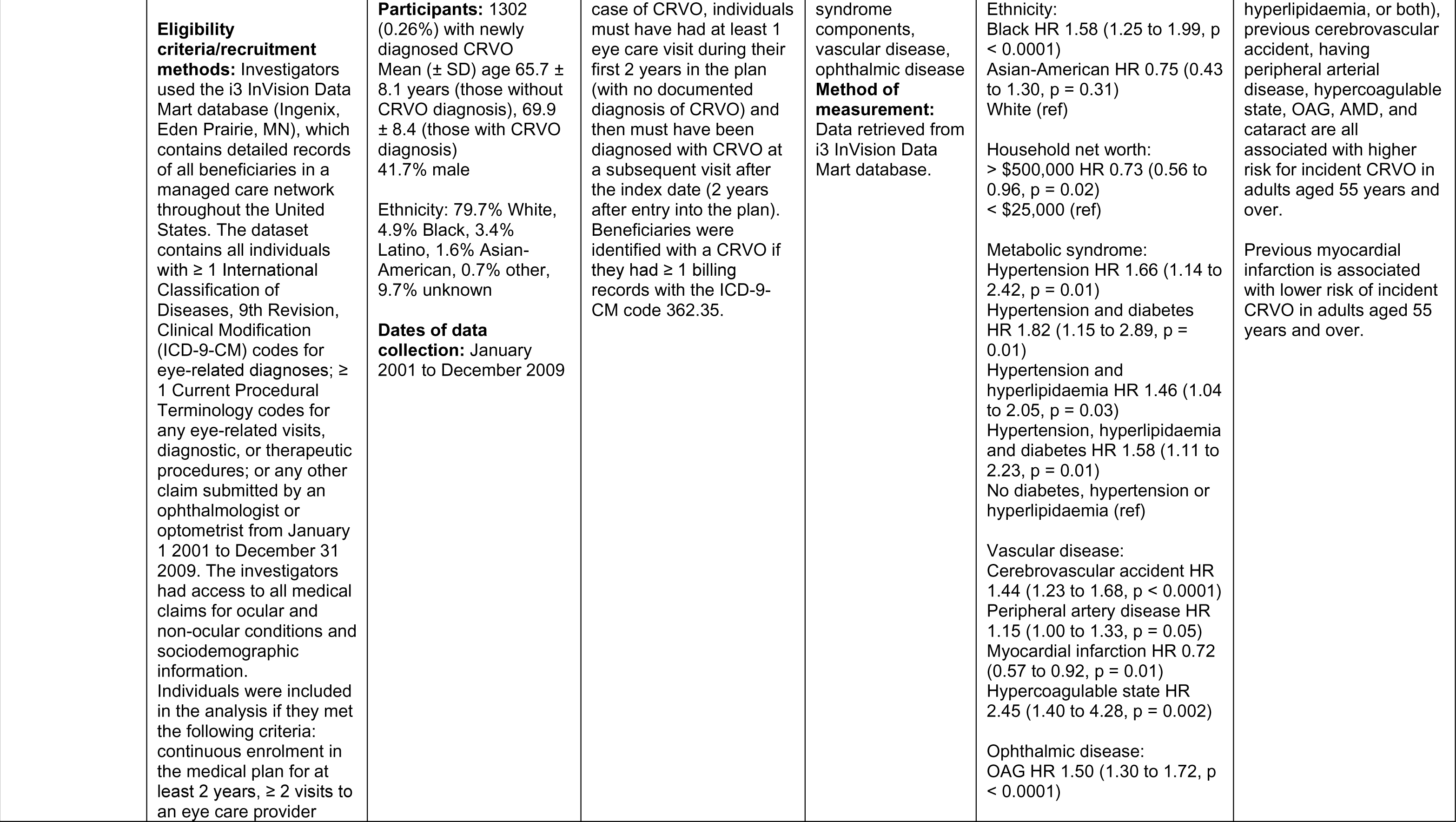
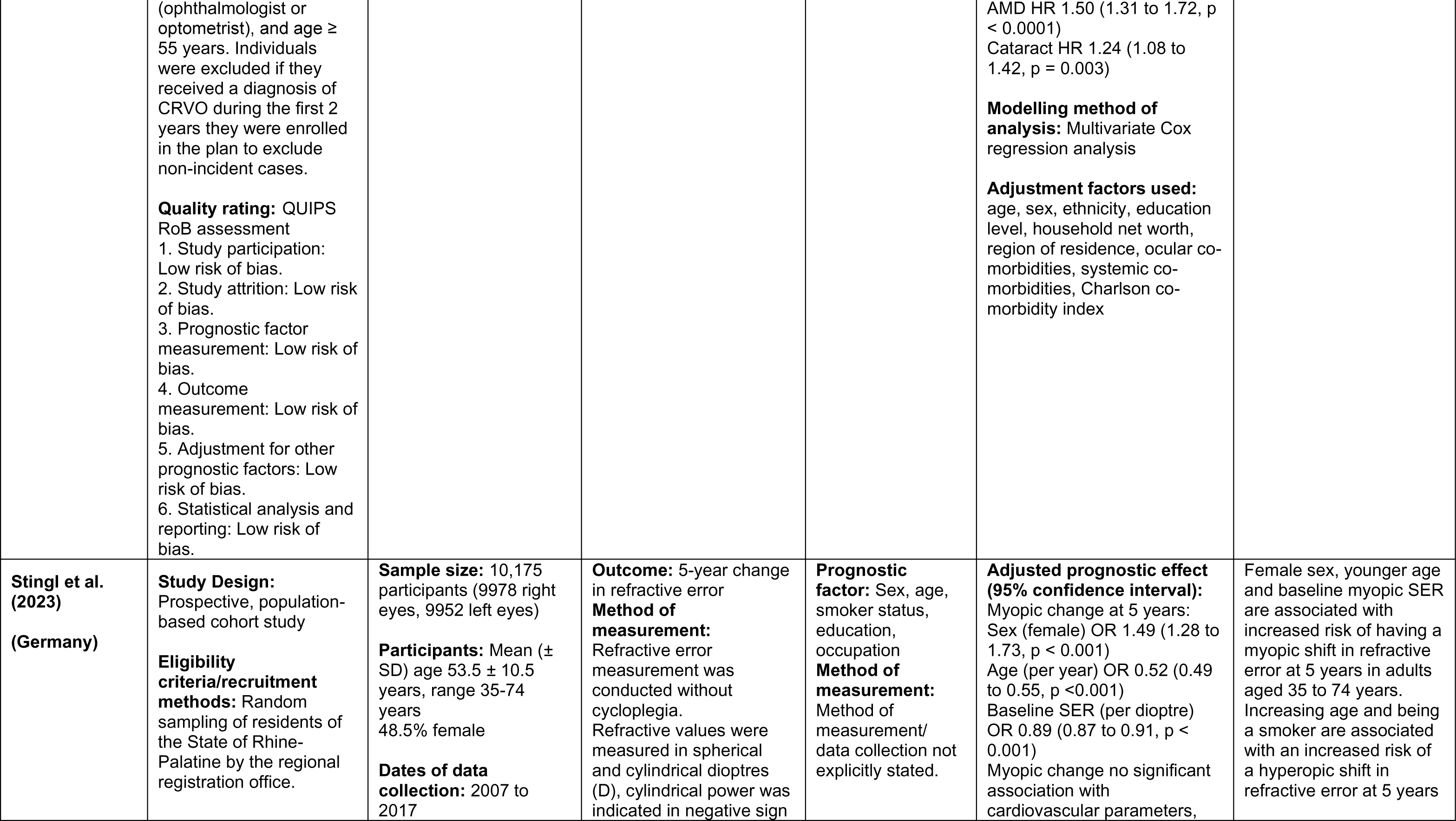
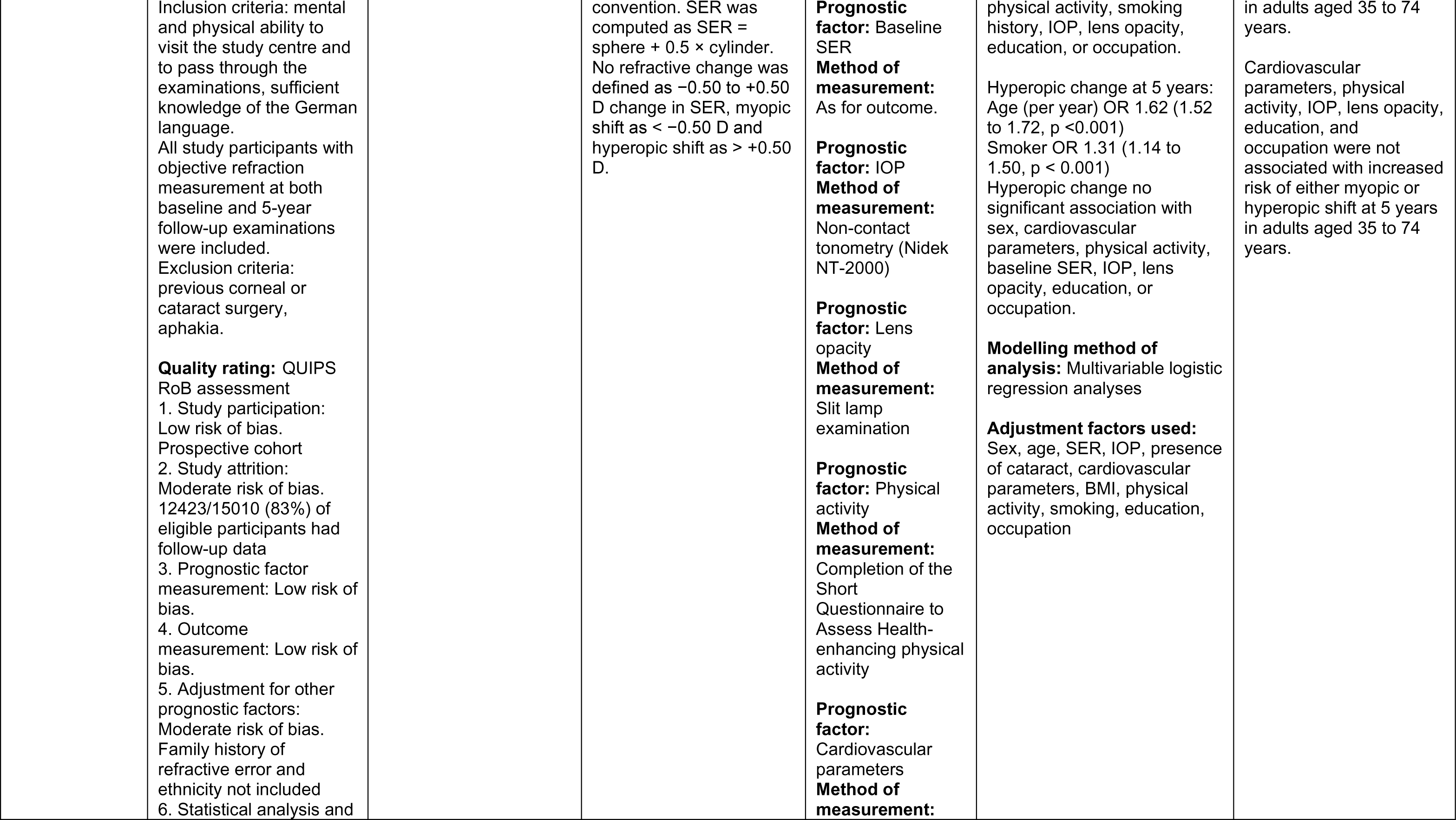
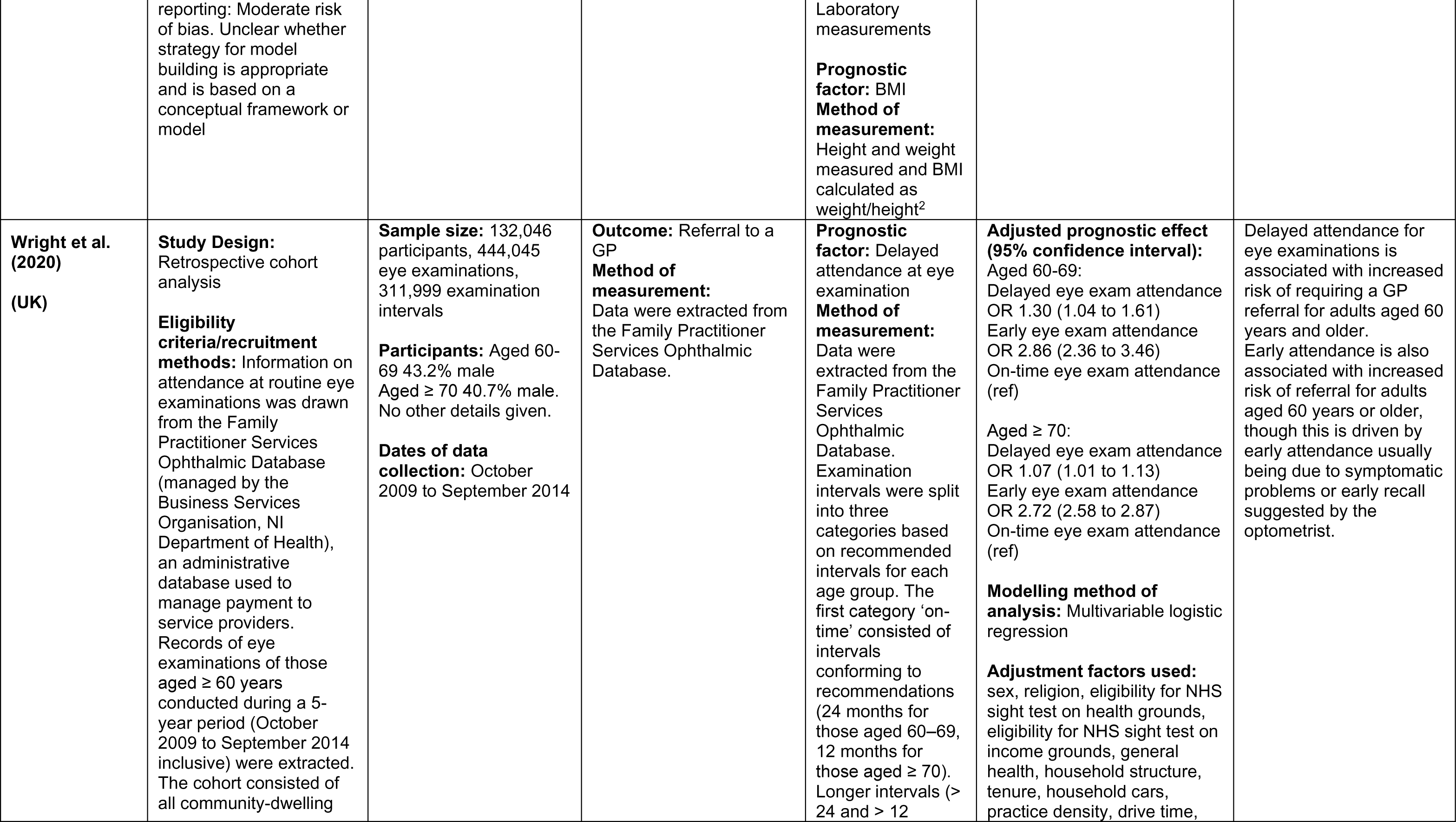
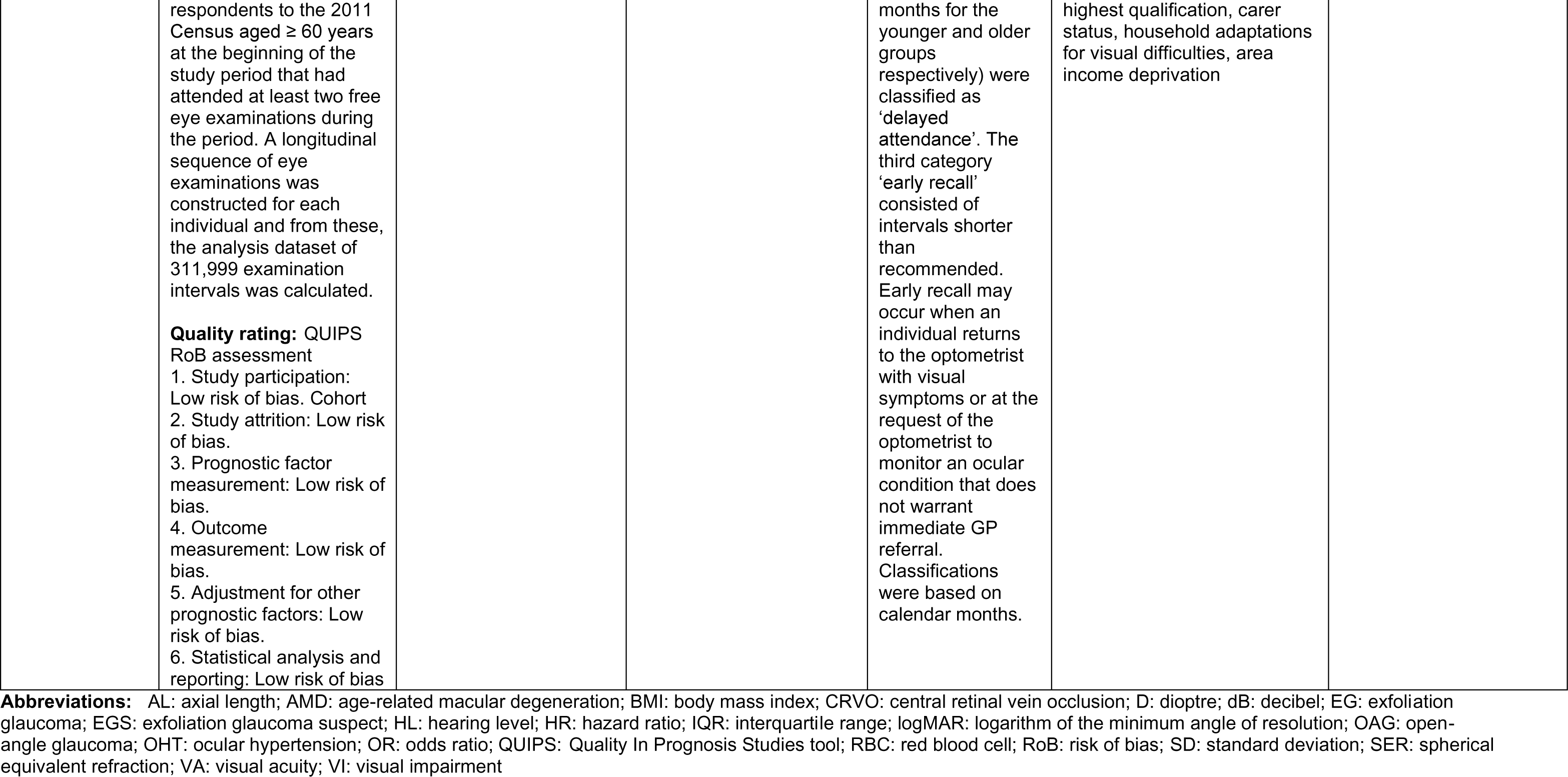
Summary of included primary studies.

| Citation (Country) | Study Details | Participants & setting | Outcomes | Prognostic factors | Key findings | Interpretation and observations |
| --- | --- | --- | --- | --- | --- | --- |
| Barsam et al. (2017)<br>(UK) | **Study Design:** Case-control study | **Sample size:** 64 cases, 1794 controls<br>**Participants:** Cases: | **Outcome:** Acute corneal hydrops<br>**Method of measurement:** | **Prognostic factor:** Vernal keratoconjunctivitis<br>**Method of measurement:** | **Adjusted prognostic effect (95% confidence interval):**<br>Vernal conjunctivitis OR 15.00 (1.30 to 173.70, p = 0.026) | History of vernal conjunctivitis, asthma, and having worse visual acuity were associated with higher odds of |
|  | **Eligibility criteria/recruitment methods:** The British Ophthalmological Surveillance Unit was used to identify new cases of acute corneal hydrops that occurred between November 2009 and December 2010. Clinicians who reported a case were sent an initial questionnaire that requested information on patient demographics, the best-corrected visual acuity before the onset of hydrops, previous keratometry values, if available, and prior ophthalmic and medical history. Patients with a completed questionnaire were defined as cases and included for further analysis. Controls with keratoconus who did not have a prior history of an acute corneal hydrops were identified from the public care hospital system from nine ophthalmic centres in the UK selected by a clustered, stratified random sampling procedure. The UK was divided into nine regions and selected hospitals within each region using | Mean (±SD) age 33.3 ± 12.9 years<br>75% male<br>Ethnicity: 65.1% White, 22.2% South Asian, 11.1% Black, 1.6% other<br>Controls:<br>Mean (±SD) age 36.4 ± 12.1 years<br>66.1% male<br>Ethnicity: 74.4% White, 17.1% South Asian, 4.3% Black, 4.3% other<br>**Dates of data collection:** November 2009 to December 2010 | Acute corneal hydrops was defined as the acute onset of bullous corneal oedema with an identifiable break in the Descemet's layer in the presence of keratoconus. | Clinician-completed questionnaire, based on clinical assessment.<br>**Prognostic factor:** Asthma<br>**Method of measurement:** Clinician-completed questionnaire, based on self-reported symptoms.<br>**Prognostic factor:** VA in worse eye<br>**Method of measurement:** Clinician-completed questionnaire. | Asthma OR 4.92 (1.22 to 19.78, $p = 0.025$)<br>VA in worse eye OR 4.11 (1.18 to 14.32, $p = 0.026$)<br>**Modelling method of analysis:** Backward stepwise multiple variable logistic regression<br>**Adjustment factors used:** Not stated | developing acute corneal hydrops in people with keratoconus.<br>The multivariable model only included 15 cases and 144 controls. |

| Citation<br>(Country) | Study Details | Participants &<br>setting | Outcomes | Prognostic<br>factors | Key findings | Interpretation and<br>observations |
| --- | --- | --- | --- | --- | --- | --- |
|  | computer-generated random numbers with the probability of selection proportional to the number of ophthalmic consultants who worked in each hospital. A local investigator at each centre then retrieved the case notes of 20 consecutive patients with keratoconus who had not had an acute corneal hydrops in the order they attended clinic. The same demographic and clinical data were collected for both cases and controls.<br>**Quality rating:** QUIPS<br>RoB assessment<br>1. Study participation: High risk of bias. Case control study design. Prognostic data was collected after the outcome was known.<br>2. Study attrition: High risk of bias. 88% of eligible cases returned questionnaire. 21% of cases included in analysis<br>3. Prognostic factor measurement: Low risk of bias.<br>4. Outcome measurement: Low risk of bias.<br>5. Adjustment for other prognostic factors: Low risk of bias. Relevant factors identified from the literature<br>6. Statistical analysis and reporting: Moderate risk of bias. Unclear whether strategy for model building is appropriate and is based on a conceptual framework or model |  |  |  |  |  |
| Ekström (2012) (Sweden) | **Study Design:** Population-based, longitudinal study<br>**Eligibility criteria/recruitment methods:** In 1984–1986, a population survey was conducted in the municipality of Tierp, south central Sweden. Its target population comprised 2429 residents 65–74 years of age. The size of the sample was limited to about one-third of the target population. Participants with normal and reliable visual fields, who completed the population survey, were invited to the follow-up study. To increase the cohort, 14 patients diagnosed with ocular hypertension at the Eye Department in Tierp in 1984–1986 were included. A further | **Sample size:** 679 participants<br>**Participants:** 61.9% age 65–69 years 38.1% age 70–74 years 59.6% female Mean (± SD) follow-up time 9.0 ± 4.3 years<br>**Dates of data collection:** 1984 to 2006 | **Outcome:** Incident open-angle glaucoma (OAG)<br>**Method of measurement:** Patients diagnosed with OAG via supra-threshold visual field testing underwent manual Goldmann perimetry and repeated visual field testing using Computer threshold test logic. Threshold fields were sent for grading by an ophthalmologist otherwise unconnected with the study. Patients deemed to have progressive disease were classed as definite OAG cases. The glaucoma case records of patients with non-progressive disease, or missing threshold fields, were reviewed by an ophthalmologist, including optic disc characteristics. Patients were then classified as either definite OAG or | **Prognostic factor:** Age<br>**Method of measurement:** Self-reported<br>**Prognostic factor:** IOP<br>**Method of measurement:** Goldmann applanation tonometry<br>**Prognostic factor:** Pseudoexfoliation<br>**Method of measurement:** Not stated | **Adjusted prognostic effect (95% confidence interval):** Age (per year) HR 1.15 (1.05 to 1.26)<br>Mean IOP ≥ 25 mmHg and pseudoexfoliation HR 2.38 (1.87 to 3.03)<br>Time-dependent (per 10 years): Mean IOP ≥ 25 mmHg HR 15.4 (4.52 to 52.1)<br>Mean IOP 20–24.99 mmHg HR 3.92 (2.13 to 7.22)<br>Mean IOP < 20 mmHg (ref)<br>**Modelling method of analysis:** Cox proportional hazard models<br>**Adjustment factors used:** Not clearly stated | Increasing age and increasing IOP are associated with increased risk of incident OAG in adults aged 65–74 years. Mean IOP ≥ 25 mmHg concurrent with pseudoexfoliation is associated with an increased risk of incident OAG in adults aged 65–74 years.<br>There is a risk of some double reporting between this study and Ekström & Hårleman (2023). |

| Citation<br>(Country) | Study Details | Participants &<br>setting | Outcomes | Prognostic<br>factors | Key findings | Interpretation and<br>observations |
| --- | --- | --- | --- | --- | --- | --- |
|  | 259 people, participating in a case-control study in 1988–1995, were also recruited. Those enrolled were in the age range of 65–74 years and underwent the same baseline examination as those in the population survey.<br>Exclusion criteria: previous treatment for OAG, previous cataract surgery<br>**Quality rating:** QUIPS<br>RoB assessment<br>1. Study participation: Low risk of bias.<br>Prospective cohort.<br>2. Study attrition: Low risk of bias.<br>3. Prognostic factor measurement: Moderate risk of bias. Unclear whether prognostic factor measures were updated during follow-up.<br>4. Outcome measurement: Low risk of bias.<br>5. Adjustment for other prognostic factors: Moderate risk of bias. Unclear how factors were chosen. It appears family history of glaucoma was not included in the multivariable model. |  | not. |  |  |  |
|  | 6. Statistical analysis and reporting: Low risk of bias. |  |  |  |  |  |
| **Ekström & Hårleman (2023)**<br>**(Sweden)** | **Study Design:** Nested case-control study<br>**Eligibility criteria/recruitment methods:** Eligibility criteria for entry into the study included being a resident in one of the two rural districts of Tierp or Älvkarleby in the north of Uppsala County, south central Sweden, and being 55–84 years of age at the first consultation for eye-related problems at the Tierp Health Centre during the two recruitment periods, January 1988 to December 1995, or June 2003 to December 2003. In addition, the participants had to fulfil the IOP criteria for the study. The vast majority were referred from opticians or general practitioners. People who had seen an eye care provider in the last 3 years, were using pressure-reducing therapy, or had a history of intraocular surgery were not eligible. | **Sample size:** 481 participants<br>**Participants:** Cases: n = 99<br>12 cases age 55-64 years, 38 cases age 65-74 years, 49 cases age 75-84 years<br>50 cases male<br>Controls: n = 382<br>80 controls age 55-64 years, 183 controls age 65-74 years, 119 controls age 75-84 years<br>139 controls male<br>**Dates of data collection:** 1988 to 2003 | **Outcome:** Incident OAG<br>**Method of measurement:** OAG was classified by a repeatable visual field defect in either eye, consistent with glaucoma and not explained by other causes.<br>Participants with normal screening fields, who developed an abnormal test point within 2 years, were counted as incident OAG.<br>Patients with a totally excavated optic disc and visual acuity < 0.05, unable to undergo automated perimetry, were also included in the OAG cases. | **Prognostic factor:** Age, sex, family history of glaucoma<br>**Method of measurement:** Self-reported<br>**Prognostic factor:** IOP<br>**Method of measurement:** Goldmann applanation tonometry<br>**Prognostic factor:** Hypertension, ischaemic heart disease<br>**Method of measurement:** Self-reported, obtained from medical records (medical records prioritised if a discrepancy) | **Adjusted prognostic effect (95% confidence interval):**<br>Age:<br>75-84 years OR 3.02 (1.13 to 8.08)<br>65-74 years OR 1.15 (0.44 to 3.00)<br>55-64 years (ref)<br>Sex:<br>Male OR 1.77 (0.91 to 3.43)<br>Female (ref)<br>Positive family history of OAG OR 3.21 (1.38 to 7.45)<br>IOP (per 5 mmHg) OR 4.04 (2.91 to 5.62)<br>Pseudoexfoliation OR 1.27 (0.63 to 2.57)<br>Treated hypertension OR 0.58 (0.29 to 1.15)<br>Ischaemic heart disease OR 2.41 (1.15 to 5.06)<br>**Modelling method of analysis:** Multiple logistic regression analyses<br>**Adjustment factors used:** Not stated | Increasing age, increasing IOP, positive family history of OAG, and ischaemic heart disease are associated with increased risk of incident OAG in adults aged 55-84 years.<br>There is no evidence that sex and pseudoexfoliation are associated with increased risk of OAG in adults aged 55-84 years. The effect of pseudoexfoliation on glaucoma risk is mediated by elevated IOP.<br>There is a risk of some double reporting between this study and Ekström (2012). |

| Citation<br>(Country) | Study Details | Participants &<br>setting | Outcomes | Prognostic<br>factors | Key findings | Interpretation and<br>observations |
| --- | --- | --- | --- | --- | --- | --- |
|  | With the intention of reaching an equal distribution of IOP readings around 22 mmHg, the pressure limit for inclusion in the study was changed once or twice every year, with a lower level of 18 mmHg, depending on the results of those already included. From January 1995 to November 1995, people with an IOP < 18 mmHg were recruited, while all pressures were accepted in December 1995. From June to December 2003, the IOP had to be < 17 or ≥ 35 mmHg.<br>**Quality rating:** QUIPS<br>RoB assessment<br>1. Study participation: Low risk of bias. Nested case control study design<br>2. Study attrition: Low risk of bias.<br>3. Prognostic factor measurement: Moderate risk of bias. Unclear whether prognostic factor measures were updated during follow-up.<br>4. Outcome measurement: Low risk of bias.<br>5. Adjustment for other prognostic factors: Low risk of bias. Unclear how factors were chosen. |  |  |  |  |  |
|  | 6. Statistical analysis and reporting: Moderate risk of bias. Unclear whether strategy for model building is appropriate and is based on a conceptual framework or model |  |  |  |  |  |
| Elmore et al. (2022)<br>(USA) | **Study Design:** Prospective cohort analysis<br>**Eligibility criteria/recruitment methods:** Post-menopausal women. Participants had to be enrolled in both the Women's Health Initiative Memory Study and the Women's Health Initiative Sight Exam Study (WHI-SE); two ancillary studies conducted in the Women's Health Initiative Hormone Therapy (WHI HT) trial. Exclusion criteria: missing or ungradable fundus photos, unreliable red blood cell fatty acid measures, self-reported energy intake above 5000 kcals or below 600 kcals, missing covariate data.<br>**Quality rating:** QUIPS RoB assessment | **Sample size:** 1076 participants<br>**Participants:** No AMD, n = 938 (follow-up): 59.0% < 70 years, 32.5% 70-74 years, 8.5% 75+ years<br>Ethnicity 89.8% White, 6.5% Black, 1.8% Hispanic, 0.9% Asian or Pacific Islander, 0.1% American Indian or Alaskan Native, 1.0% other<br>Incident AMD, n = 138 (follow-up): 48.6% < 70 years, 39.1% 70-74 years, 12.3% 75+ years<br>Ethnicity 92.0% White, 4.3% Black, 0.0% Hispanic, 0.7% Asian or Pacific Islander, 0.0% American Indian or Alaskan Native, 2.9% other<br>**Dates of data collection:** 2000 to 2015 | **Outcome:** Incident AMD<br>**Method of measurement:** Self-reported. Prevalent AMD status was determined from stereoscopic 30° colour fundus photographs taken as part of the WHI-SE Study (2000–2002) and graded using the Wisconsin Age-Related Maculopathy Grading Scheme. This identified 240 prevalent AMD cases and 1216 cases without AMD. Participants enrolled in the WHI Extension Study 1 (2005–2010) and Extension Study 2 (2010–2015) received a mailed Medical History Update survey annually. This survey asked, "since the date on the front of this form, has a doctor told you for the first time that you have macular degeneration?" | **Prognostic factor:** RBC fatty acid<br>**Method of measurement:** Fasting blood samples were collected at WHI HT baseline (1993–1998). RBC fatty acid composition was analysed using gas chromatography with flame ionization detection and then expressed as percent weight of total fatty acids.<br>**Prognostic factor:** Dietary intake of fatty acids and fish<br>**Method of measurement:** A subset of the study sample were additionally enrolled in the WHI | **Adjusted prognostic effect (95% confidence interval):**<br>RBC polyunsaturated fatty acid levels:<br>No significant association between any RBC polyunsaturated fatty acid levels and incident AMD<br>Dietary intake of fatty acids:<br>No significant association between dietary intake of any fatty acids and incident AMD<br>Dietary intake of fish:<br>≥ 1 serving per week HR 0.91 (0.53 to 1.58)<br>≥ 1 serving per month and < 1 serving per week HR 0.86 (0.48 to 1.54)<br>None or < 1 serving per month (ref)<br>Dietary intake of dark fish:<br>≥ 1 serving per week HR 1.20 (0.79 to 1.81)<br>≥ 1 serving per month and < 1 serving per week HR 0.60 (0.34 to 1.04)<br>None or < 1 serving per month (ref) | There is no association between fatty acid intake or fish intake and incident AMD in post-menopausal women.<br>RBC polyunsaturated fatty acid levels are a longer-term biomarker of fatty acid intake. |
|  | 1. Study participation: Low risk of bias. Prospective cohort<br>2. Study attrition: Moderate risk of bias. Only participants enrolled in the WHI extension study (1076/1216) provided incidence data.<br>3. Prognostic factor measurement: Moderate risk of bias. Self-reported via questionnaire<br>4. Outcome measurement: Moderate risk of bias. Self-reported AMD<br>5. Adjustment for other prognostic factors: Low risk of bias.<br>6. Statistical analysis and reporting: Low risk of bias |  | Self-reported AMD was categorized as “yes” or “no.”<br>Participants not followed up into the extension studies were excluded. 1076 of the 1216 women without prevalent AMD at WHI-SE baseline were followed up, and 138 women were identified as developing incident AMD. | Dietary Modification Trial. Dietary variables were collected via a modified Block food frequency questionnaire assessing usual dietary intake during the previous three months. | **Modelling method of analysis:** Cox proportional hazards regression modelling<br>**Adjustment factors used:** Age, race/ethnicity, pack-years of smoking, assignment to clinical trial (the hormone therapy trial, the dietary modification trial, or the calcium and vitamin D trial), hypertension, BMI, recreational physical activity, diabetes status |  |
| **Fernández-Montero et al. (2017)**<br>**(Spain)** | **Study Design:** Longitudinal, prospective cohort study<br>**Eligibility criteria/recruitment methods:** The SUN project is a multipurpose, prospective, dynamic cohort of young adult university graduates conducted in Spain. The recruitment of participants started in 1999 and is permanently open. Mailed questionnaires are used to gather baseline | **Sample size:** 10,401 participants<br>**Participants:** 3180 reported pregnancies<br>Pregnancy mean (± SD) age $28.4 \pm 4.2$ years<br>No pregnancy mean (± SD) age $35.3 \pm 8.5$ years<br>**Dates of data collection:** 1999 to 2013 | **Outcome:** Incident myopia or progression of myopia<br>**Method of measurement:** All follow-up questionnaires included the following question: “Have you been diagnosed by a medical doctor of new-onset myopia or a progression of 0.5 or more dioptres in myopia, since the last questionnaire you filled in?”<br>Participants who responded yes to this question were considered incident cases. | **Prognostic factor:** Pregnancy<br>**Method of measurement:** Pregnancies were assessed in each biennial follow-up questionnaire. All questionnaires, except for the first 2-year follow-up, included the following question: “Have you been diagnosed by a doctor of a pregnancy since the last questionnaire? If | **Adjusted prognostic effect (95% confidence interval):** Pregnancy HR 0.58 (0.49 to 0.69, $p < 0.001$)<br>**Modelling method of analysis:** Multivariable Cox regression analysis<br>**Adjustment factors used:** Age, BMI, total energy intake, Mediterranean Diet, smoking habits, computer use, educational level, sleeping behaviour, time of television watching, physical activity | Pregnancy is associated with a decreased risk of developing myopia or progression of existing myopia in women aged 20-50 years. |
|  | characteristics and information on diet, lifestyles and new medical diagnoses of disease every 2 years. This study only included women with a minimum of 2-years follow-up between the ages of 20 to 50 years.<br>**Quality rating:** QUIPS RoB assessment<br>1. Study participation: Low risk of bias. Prospective cohort<br>2. Study attrition: Moderate risk of bias. 14% lost to follow-up<br>3. Prognostic factor measurement: Moderate risk of bias. Self-reported via questionnaire. Time outdoors was estimated & only at baseline.<br>4. Outcome measurement: Moderate risk of bias. Self-reported myopia<br>5. Adjustment for other prognostic factors: Low risk of bias.<br>6. Statistical analysis and reporting: Low risk of bias. |  | Participants were considered a case of myopia progression when they reported a new diagnosis of myopia or an increase in myopia of at least -0.50 D in one eye. | so, please report estimated due data (month/year)". |  |  |
| **Gopinath et al. (2014)**<br>**(Australia)** | **Study Design:** Longitudinal, population-based cohort study | **Sample size:** 2443 participants<br>**Participants:** 1<sup>st</sup> quintile of total diet score: | **Outcome:** 5-year incidence of dual sensory impairment<br>**Method of measurement:** | **Prognostic factor:** Diet<br>**Method of measurement:** At baseline, dietary data were | **Adjusted prognostic effect (95% confidence interval):** Total diet score 5<sup>th</sup> quintile vs. 1<sup>st</sup> quintile and dual sensory impairment OR 1.03 (0.30 to 3.50) | Diet is not associated with 5-year incidence of dual sensory impairment in adults aged over 49 years. |
|  | **Eligibility criteria/recruitment methods:** Part of the Blue Mountains Eye Study (BMES). Following a door-to-door census of the region, baseline examinations of 3654 residents aged > 49 years were conducted during 1992-4 (BMES-1). Surviving baseline participants were invited to attend 5-year follow-up examinations (1997-9, BMES-2), at which 2334 (75.1% of survivors) and an additional 1174 newly eligible residents were examined. At BMES-2, 2956 participants had audiometric testing performed. At BMES-3 (2002-4), 1952 participants were re-examined. Visual acuity data were collected at all three BMES examinations.<br>**Quality rating:** QUIPS RoB assessment<br>1. Study participation: Moderate risk of bias. Prospective cohort. 513/2956 (17%) were ineligible due to missing data at baseline.<br>2. Study attrition: Moderate risk of bias. Unclear how many | Mean (± SD) age 67.3 ± 9.5 years, 53.9% male<br>5<sup>th</sup> quintile of total diet score:<br>Mean (± SD) age 66.8 ± 8.4 years, 31.9% male<br>**Dates of data collection:** 1992 to 2004 | Pure-tone audiometry was performed by audiologists in sound-treated booths, using TDH-39 earphones and Madsen OB822 audiometers. Sound-proof rooms were set-up according to International Standards Organization protocol 8253-2. Bilateral hearing impairment was determined as the pure-tone average of audiometric hearing thresholds at 500, 1000, 2000, and 4000 Hz (PTA0.5 to 4 kHz) in the better ear, defining any hearing loss as PTA0.5 to 4kHz > 25 dB HL; mild hearing loss as PTA0.5 to 4kHz > 25 to 40 dB HL; and moderate to severe hearing loss as PTA0.5 to 4kHz > 40 dB HL. Monocular distance logMAR VA was measured with forced choice procedures using a retro-illuminated chart according to the early treatment diabetic retinopathy Study protocol. Both presenting VA (with current eyeglasses, if worn) and after subjective refraction (best-corrected VA) were measured. | collected using a 145-item self-administered food frequency questionnaire. A total diet score was established by allocating scores for intakes of selected food groups and nutrients for each participant as described in the Dietary Guidelines for Australian Adults. The total diet score is divided into ten components, and each component has a possible score ranging from 0 to 2. A maximum score of 2 was given to subjects who met the recommendations with pro-rated scores for lower intakes. These were then summated providing a final score ranging between 0 and 20 with higher scores indicating closer adherence to the dietary guidelines. | **Modelling method of analysis:** Discrete linear logistic models<br>**Adjustment factors used:** age, sex, education, current smoking, noise exposure, type 2 diabetes |  |
|  | participants completed the 5 year follow-up.<br>3. Prognostic factor measurement: Low risk of bias. Some factors based on self-report others measured.<br>4. Outcome measurement: Low risk of bias.<br>5. Adjustment for other prognostic factors: Moderate risk of bias. Unclear how factors were chosen.<br>6. Statistical analysis and reporting: Low risk of bias. |  | Any visual impairment was defined as presenting VA of the better eye < 20/40). Dual sensory impairment was defined as concurrent visual (either presenting or best-corrected) and hearing impairment, as determined using the above definitions. |  |  |  |
| **Guggenheim et al. (2012)**<br>**(UK)** | **Study Design:** Opportunistic, longitudinal study<br>**Eligibility criteria/recruitment methods:** Avon Longitudinal Study of Parents and Children (ALSPAC) cohort: Pregnant women with an expected date of delivery between April 1, 1991 and December 31, 1992, resident in the former Avon health authority area in Southwest England, were eligible to participate in the study. A cohort of 14,541 pregnant women was established, resulting in 13,988 | **Sample size:** 2005 participants with complete information on predictor variables and either were seen at the age 15-year clinic or who were already known to have become myopic when they attended the 12-year clinic.<br>**Participants:** Mean (± SD) age 11.7 ± 0.2 years<br>49.1% male<br>**Dates of data collection:** April 1991 to c.2008 | **Outcome:** Incident myopia after age 11 years<br>**Method of measurement:** Non-cycloplegic autorefraction. Participants were classified as myopic if the average of the SERs in their right and left eyes was ≤ -1.00 D. Subjects were classified as emmetropic or hyperopic if the averaged SER in their right and left eyes was ≥ -0.25 D. | **Prognostic factor:** Parental myopia<br>**Method of measurement:** Participants' parents completed a questionnaire that included the question "How would you rate your sight without glasses?" and were classed as myopic if they answered "can't see clearly at distance" for both eyes.<br>**Prognostic factor:** Time spent reading | **Adjusted prognostic effect (95% confidence interval):**<br>Parental myopia:<br>1 myopic parent OR 1.175 (0.900 to 1.533, $p = 0.236$)<br>2 myopic parent OR 1.143 (0.718 to 1.818, $p = 0.574$)<br>No myopic parents (ref)<br>Time spent reading (low vs. high):<br>OR 1.323 (1.023 to 1.712, $p = 0.033$)<br>Time spent outdoors:<br>OR 0.65 (0.45 to 0.96, $p = 0.029$)<br>Sex (male vs. female):<br>OR 1.058 (0.810 to 1.382, $p = 0.679$) | Increased time spent reading is associated with increased risk of developing myopia after age 11.<br>Increased time spent outdoors is associated with lower risk of developing myopia after age 11.<br>Parental myopia, sex and physical activity/sedentary behaviour are not associated with incident myopia after age 11. |
|  | children who were alive at 12 months of age. Data collection has been via various methods, including self-completion questionnaires sent to the mother and her partner, and after age 5 to the child, as well as direct assessments and interviews in a research clinic, biological samples, and linkage to school and hospital records. All children still participating in ALSPAC were invited approximately yearly (starting at age 7 years) to sessions where a number of assessments and interviews, tailored to their age, took place. Vision-related data were included in the assessments carried out at the 7-, 10-, 11-, 12-, and 15-year clinics.<br>**Quality rating:** QUIPS RoB assessment<br>1. Study participation: Low risk of bias. Prospective cohort.<br>2. Study attrition: Moderate risk of bias. Refractive error data was available for 9109/13988 children.<br>3. Prognostic factor measurement: Moderate |  |  | **Method of measurement:** When the participants were aged 8 to 9 years, mothers completed a questionnaire including the question "On normal days in school holidays, how much time on average does your child spend each day reading books for pleasure?" Children were classified as spending a "high" amount of time reading for pleasure if the response was "1–2 hours" or "3 or more hours," and as "low" otherwise.<br>**Prognostic factor:** Time spent outdoors<br>**Method of measurement:** At 8 to 9 years of age, a questionnaire was completed by participants' mothers, asking "On a (weekend day)/(school week day), how much time on average | Physical activity/sedentary behaviour:<br>Mean counts per minute for whole week:<br>OR 0.887 (0.773 to 1.017, p = 0.086)<br>Time with moderate to vigorous activity per day:<br>OR 0.877 (0.764 to 1.006, p = 0.062)<br>Time with sedentary counts:<br>OR 1.095 (0.959 to 1.251, p = 0.180)<br>**Modelling method of analysis:** Multivariate logistic regression analyses<br>**Adjustment factors used:** Parental myopia, time reading, sex, physical activity/sedentary behaviour, time spent outdoors.<br>Three different measures for physical activity/sedentary behaviour were used and modelled separately. The ORs were similar regardless of which measure was adjusted for. |  |

| Citation<br>(Country) | Study Details | Participants &<br>setting | Outcomes | Prognostic<br>factors | Key findings | Interpretation and<br>observations |
| --- | --- | --- | --- | --- | --- | --- |
|  | risk of bias. Time outdoors, time spent reading and parental refractive error were all self-reported (by mother).<br>4. Outcome measurement: Low risk of bias.<br>5. Adjustment for other prognostic factors: Low risk of bias.<br>6. Statistical analysis and reporting: Low risk of bias. |  |  | does your child spend each day out of doors in (summer)/(winter).”<br>The response options were “None at all,” “1 hour,” “1–2 hours,” and “3 or more hours.”<br>Children were classified as spending a “high” amount of time outdoors in summer if the response was “3 or more hours,” and as “low” otherwise. For time spent outdoors in winter, children were classified as spending a “low” amount of time outdoors if the response was “None at all” or “1 hour,” and as “high” otherwise.<br>**Prognostic factor:** Sex<br>**Method of measurement:** Self-reported<br>**Prognostic factor:** Physical activity/sedentary behaviour |  |  |
|  |  |  |  | **Method of measurement:**<br>Children attending the research clinic at age 11 years were asked to wear an Actigraph accelerometer for the following 7 days. Data from the returned accelerometers were downloaded and imported into a database. Children who did not provide at least 10 hours of valid data on at least 3 separate days were omitted from the analyses. Two physical activity variables were derived from the data: Mean counts per min for the whole week, and minutes of moderate to vigorous activity per day. A variable representing sedentary behaviour was derived from count per min by defining sedentary time as less than 200 counts per min. |  |  |
| **Hopf et al. (2022)**<br>**(Germany)** | **Study Design:** Prospective, population-based cohort study<br>**Eligibility criteria/recruitment methods:** Part of the Gutenberg Health Study. Random sampling of residents of the State of Rhine-Palatine by the regional registration office, stratified by gender, decade of age, residence, baseline age of 35 to 74 years. Inclusion criteria: Phakic eyes with SER ≤ -6.00 D at baseline examination, gradable fundus photographs at baseline and 5-year follow-up.<br>**Quality rating:** QUIPS RoB assessment<br>1. Study participation: Low risk of bias.<br>2. Study attrition: Low risk of bias.<br>3. Prognostic factor measurement: Low risk of bias.<br>4. Outcome measurement: Low risk of bias.<br>5. Adjustment for other prognostic factors: Low risk of bias.<br>6. Statistical analysis and | **Sample size:** 350 participants (528 eyes)<br>**Participants (at baseline):** Without baseline myopic maculopathy: Mean age 50.23 ± 9.17 years<br>50.8% female<br>Median SER (RE) -7.19 D (IQR -8.62 to -6.25)<br>Median SER (LE) -7.25 (-8.75 to -6.50)<br>With baseline myopic maculopathy: Mean (± SD) age 56.70 ± 9.08 years<br>44.4% female<br>Median SER (RE) -9.81 D (IQR -11.47 to -7.47)<br>Median SER (LE) -8.75 (-11.25 to -7.25)<br>**Dates of data collection:** 2007 to not stated | **Outcome:** Progression of myopic maculopathy at 5 years<br>**Method of measurement:** Fundus photographs graded by two masked graders following the international photographic grading system for myopic maculopathy. A retinal specialist made the decision in a consensus meeting if the two graders disagreed.<br>Progression was defined as an increase in stage of myopic maculopathy, enlargement(s) of existing lesion, or new lesion(s) at a different spot at the posterior pole. | **Prognostic factor:** Sex, age<br>**Method of measurement:** Not explicitly stated<br>**Prognostic factor:** IOP<br>**Method of measurement:** Non-contact tonometry (Nidek NT-2000)<br>**Prognostic factor:** SER<br>**Method of measurement:** Non-cycloplegic autorefraction | **Adjusted prognostic effect (95% confidence interval):**<br>Sex (female) OR 5.54 (0.93 to 32.92, $p = 0.060$)<br>Age (per year) OR 0.94 (0.88 to 1.02, $p = 0.134$)<br>IOP (per mmHg) OR 1.62 (1.51 to 1.59, $p = 0.035$)<br>SER (per dioptre) OR 1.21 (0.99 to 1.49, $p = 0.063$)<br>**Modelling method of analysis:** Multivariable logistic regression analyses<br>**Adjustment factors used:** Sex, age, IOP, baseline SER | Increasing IOP is associated with increased risk of progression of myopic maculopathy at 5 years in adults aged 35-74 years.<br>Sex, age, and SER are not associated with increased risk of myopic maculopathy progression at 5 years in adults aged 35-74 years. |

| Citation<br>(Country) | Study Details | Participants &<br>setting | Outcomes | Prognostic<br>factors | Key findings | Interpretation and<br>observations |
| --- | --- | --- | --- | --- | --- | --- |
|  | reporting: Moderate risk of bias. Unclear whether strategy for model building is appropriate and is based on a conceptual framework or model |  |  |  |  |  |
| Irving et al.<br>(2016)<br>(Canada) | **Study Design:** Retrospective cohort analysis<br>**Eligibility criteria/recruitment methods:** Retrospective cross-sectional database of patients who presented at the University of Waterloo Optometry Clinic during a 1-year period from January 2007 to January 2008. Data were extracted for all patients whose reason for presenting was to have a routine eye examination as reported in the case history (including those presenting for employment purposes, to obtain contact lenses, or to replace spectacles). There were some patients who initially presented for a routine eye examination but reported symptoms when specifically questioned. These patients were | **Sample size:** 2656 participants<br>**Participants:** Median age 38.5 years (range 0.4 to 93.9 years) 48% male<br>**Dates of data collection:** January 2007 to January 2008 | **Outcome:** Significant change in ocular status<br>**Method of measurement:** Defined as one or more of spectacle prescription change, new critical diagnosis, or new management of an existing condition. A spectacle prescription change was considered to be significant if in at least one eye, the sphere, cylinder, or any reading addition changed by > 0.50 D from the entering to the exiting spectacle prescription, or if the cylinder axis changed as follows: > 15 degrees if the absolute value of the final cylinder value was < 1.00 D, > 10 degrees if the cylinder was $\geq 1.00$ D but < 2.00 D, or > 5 degrees if the cylinder was $\geq 2.00$ D. A critical diagnosis was considered new if it was not reported in the clinic file case history or at previous examinations. | **Prognostic factor:** Age, sex, interval between eye examinations<br>**Method of measurement:** Identified from case notes | **Adjusted prognostic effect (95% confidence interval):**<br>Age (per year) OR 1.03 (1.03 to 1.04)<br>Sex (female) OR 1.07 (0.90 to 1.29)<br>Interval between eye examinations (per year) OR 1.06 (1.02 to 1.11)<br>**Modelling method of analysis:** Multivariable logistic regression<br>**Adjustment factors used:** Assessment interval, sex, age | Increasing age and increasing length of time between eye examinations are associated with increased risk of experiencing a significant change in ocular status.<br>Sex was not associated with increased risk of significant change in ocular status. |
|  | excluded from the main analysis, but their overall percentage of significant change is reported for comparison.<br>**Quality rating:** QUIPS RoB assessment<br>1. Study participation: Low risk of bias.<br>2. Study attrition: Low risk of bias.<br>3. Prognostic factor measurement: Low risk of bias.<br>4. Outcome measurement: Moderate risk of bias. Composite outcome of any critical ocular disorders or abnormal findings<br>5. Adjustment for other prognostic factors: Low risk of bias.<br>6. Statistical analysis and reporting: Low risk of bias. |  | A management (not including prescription change) was considered new if it was not initiated at a previous visit or if there was a change compared to the last available information. New managements included referrals, new treatment, or changes in monitoring schedule. |  |  |  |
| **Kang et al. (2012)**<br>**(USA)** | **Study Design:** Prospective cohort study<br>**Eligibility criteria/recruitment methods:** The Nurses' Health Study is an ongoing population-based cohort of registered female nurses. | **Sample size:** 120,146 participants<br>**Participants:**<br>Women: 78,955 participants<br>Ancestry: 7% Scandinavian, 16.4% Southern European, 74.3% other white, 1.4% black, 0.7% Asian, 0.2% | **Outcome:** Incident exfoliation glaucoma or exfoliation glaucoma suspect<br>**Method of measurement:** In all biennial questionnaires from 1986, participants were asked if they had physician-diagnosed glaucoma. From among | **Prognostic factor:** Age, gender, eye colour<br>**Method of measurement:** Self-reported in questionnaires<br>**Prognostic factor:** Family | **Adjusted prognostic effect (95% confidence interval):**<br>Rate ratio (RR) of age:<br>40 to 55 years (ref)<br>55 to 60 years RR 4.33 (2.19 to 8.56)<br>60 to 65 years RR 10.43 (5.50 to 19.78)<br>65 to 70 years RR 19.88 (10.41 to 37.96) | Increasing age, positive family history of glaucoma and female gender are associated with higher risk for incident EG or EGS.<br>Eye colour is not associated with increased risk of incident EG or EGS for adult men. |
|  | The Nurses' Health Study was established in 1976 when 121,700 United States women were invited to complete a questionnaire regarding lifestyle, health behaviour, and chronic diseases. The Health Professionals Follow-up Study (HPFS) is an ongoing cohort created in 1986 when 51,529 male health professionals (dentists, veterinarians, pharmacists, optometrists, osteopaths, and podiatrists) completed a similar health survey. The participants in both cohorts have been followed up biennially with mailed questionnaires that have updated health and lifestyle information. The study period was 1980 through 2008 for the Nurses' Health Study and 1986 through 2008 for the HPFS. Data were collected from those who were prospectively followed for 20 years or more and who provided lifetime residence information as well as other lifestyle and health information were used to examine the | Native American or Hawaiian, 0.8% Hispanic<br>288 incident cases of EG or EGS<br>Mean (± SD) age at diagnosis 68.1 ± 6.6 years<br>Men:<br>41,191 participants<br>Ancestry: 11.2% Scandinavian, 23% Southern European, 61.3% other white, 0.9% black, 1.6% Asian, 2% Native American or Hawaiian<br>60 incident cases of EG or EGS<br>Mean (± SD) age at diagnosis 70.8 ± 6.9 years<br>**Dates of data collection:** 1980 to 2008 | participants who gave a positive response to this question, permission was obtained to retrieve their medical information. The diagnosing eye care provider of record was sent a request to complete a glaucoma questionnaire, which asked about the presence of exfoliation material or other secondary causes for elevated IOP, maximum IOP, optic nerve features, and status of the filtration apparatus and was asked to send all available visual field (VF) reports. In lieu of completing the questionnaire, eye care providers could send the complete medical records and all VF reports related to the glaucoma diagnosis. A glaucoma specialist (LRP) evaluated the questionnaire or medical record information as well as the VF data in a standardized manner for confirmation and classification.<br>Cases of either EG or EGS were analysed. Specifically, EG was defined as the presence of exfoliation material in combination with 2 or | history of glaucoma<br>**Method of measurement:** Self-reported in questionnaires. Positive family history of glaucoma was defined as a self-report of any glaucoma in biologic parents, siblings, or children. | 70 to 75 years RR 33.54 (17.23 to 65.29)<br>Over 75 years RR 46.22 (22.77 to 93.80)<br>Rate ratio (RR) of family history of glaucoma:<br>Positive history RR 2.29 (1.39 to 3.78)<br>Negative history (ref)<br>Rate ratio (RR) of eye colour (males only):<br>Hazel/green/medium RR 0.87 (0.43 to 1.74)<br>Brown/dark RR 0.84 (0.42 to 1.68)<br>Blue/light (ref)<br>Rate ratio (RR) of gender:<br>Male RR 0.32 (0.23 to 0.46)<br>Female (ref)<br>**Modelling method of analysis:**<br>Cox proportional hazard analysis<br>**Adjustment factors used:**<br>Age, race, family history of glaucoma, BMI, self-reported hypertension, diabetes mellitus, high cholesterol, myocardial infarction, geographical tier |  |
|  | descriptive epidemiologic features of exfoliation glaucoma (EG) or exfoliation glaucoma suspect (EGS). Participants contributed person-time until confirmed EG or EGS, self-report of glaucoma, death, loss to follow-up, diagnosis of cancer other than nonmelanoma skin cancer, self-report of cataract extraction, or the end of the study (2008).<br>**Quality rating:** QUIPS RoB assessment<br>1. Study participation: Moderate risk of bias. Prospective cohort. Confirmation received from diagnosing eye-care provider of 6870/10737 of those self-reporting glaucoma.<br>2. Study attrition: Low risk of bias.<br>3. Prognostic factor measurement: Moderate risk of bias. Self reported via questionnaire<br>4. Outcome measurement: Moderate risk of bias. Self-reported glaucoma - but verified by review of medical records.<br>5. Adjustment for other prognostic factors: Low risk of bias. |  | more reliable tests showing reproducible VF loss consistent with glaucoma, and EGS was defined as the presence of exfoliation material in combination with (1) a history of IOP of more than 21 mmHg; or (2) a cup-to-disc ratio of 0.6 or more or the inter-eye difference in a cup-to-disc ratio of 0.2 or more; or (3) only 1 reliable test showing VF loss consistent with glaucoma. Those with a presence of exfoliation material only without any VF loss or elevation in IOP or abnormal cup-to-disc ratios (as defined above) were not considered as cases of EG or EGS. |  |  |  |
|  | 6. Statistical analysis and reporting: Low risk of bias. |  |  |  |  |  |
| Keel et al. (2017)<br>(Australia) | **Study Design:** Population-based, cross-sectional study<br>**Eligibility criteria/recruitment methods:** Thirty sites, across five Remoteness Areas (Major City, Inner Regional, Outer Regional, Remote and Very Remote), were selected using a multi-stage, random cluster sampling methodology. To obtain a nationally representative sample of the population, 100 non-Indigenous Australians aged 50 years and older and 50 Indigenous Australians aged 40 years and older were to be recruited at each site. Recruiters went door-to-door to determine the eligibility of the residents. All eligible residents were invited to participate.<br>**Quality rating:** QUIPS RoB assessment<br>1. Study participation: Low risk of bias. Cross sectional study<br>2. Study attrition: Low risk | **Sample size:** 3098 participants<br>**Participants:** Referred (n = 994): Mean (± SD) age 67 ± 10 years<br>51.8% male<br>Not referred (n = 2104): Mean (± SD) age 66.4 ± 9.5 years<br>43.8% male<br>**Dates of data collection:** c.2015 | **Outcome:** Rates of eye care referral<br>**Method of measurement:** A referral protocol was developed by study investigators in conjunction with ophthalmologists. Participants were provided with a referral letter to be taken to their optometrist or local doctor if they met any of the following referral criteria: (1) evidence of eye disease or visual impairment detected during the NEHS eye examination; (2) participants with diabetes who had not undergone a screening eye examination within the timeframe recommended by the National Health and Medical Research Council diabetic retinopathy guidelines, or (3) individuals without diabetes who had undergone an eye examination in the past 5 years. Participants who were already under ophthalmological care were not provided with a | **Prognostic factor:** Sex, age, time since previous eye examination, geographical remoteness, years of education, diabetes, stroke<br>**Method of measurement:** Each participant underwent an interviewer-administered questionnaire to collect information on socio-demographic factors, history of ocular problems, stroke and diabetes. | **Adjusted prognostic effect (95% confidence interval):**<br>Age (per year) OR 1.02 (1.01 to 1.02, p < 0.001)<br>Sex (male) OR 1.24 (1.06 to 1.46, p = 0.007)<br>Years of education (per year) OR 0.98 (0.96 to 1.00, p = 0.11)<br>Diabetes (self-reported) OR 0.83 (0.67 to 1.04, p = 0.11)<br>Stroke (self-reported) OR 1.00 (0.99 to 1.00, p = 0.64)<br>Geographical remoteness OR 1.04 (0.979 to 1.10, p = 0.27)<br>Time since last eye examination OR 1.15 (1.12 to 1.19, p < 0.001)<br>**Modelling method of analysis:** Multivariate logistic regression analyses<br>**Adjustment factors used:** Sex, age, time since previous eye examination, geographical remoteness, years of education, diabetes, stroke | Increasing age, male sex, and longer time period since last eye examination are all associated with higher eye care referral rates in adults aged 50 years and older.<br>Years of education, history of diabetes or stroke, and geographical remoteness are not associated with eye care referral rates in adults aged 50 years and older.<br>Only the results of non-Indigenous participants have been extracted. |
|  | of bias.<br>3. Prognostic factor measurement: Moderate risk of bias. Diabetes and stroke history self-reported<br>4. Outcome measurement: Low risk of bias.<br>5. Adjustment for other prognostic factors: Low risk of bias.<br>6. Statistical analysis and reporting: Moderate risk of bias. Only covariates that were significant in univariate analysis were included in the multivariable model. |  | referral unless new pathology was suspected. |  |  |  |
| **Khachatryan et al. (2015)**<br>**(USA)** | **Study Design:** Prospective cohort study<br>**Eligibility criteria/recruitment methods:** Participants included in this study were selected from the African Descent and Glaucoma Evaluation Study (ADAGES) and Diagnostic Innovations in Glaucoma Study (DIGS). Suspect glaucoma defined as a history of elevated IOP and/or an optic disc appearance suspicious of glaucoma but normal visual fields at study entry. Elevated IOP defined as IOP > 21 | **Sample size:** 357 participants (636 eyes)<br>**Participants:** Mean (± SD) age at entry 58.1 ± 12.3 years<br>65% female<br>67% European descent<br>33% African descent<br>Mean (± SD) follow-up time 7.1 ± 2.4 years<br>**Dates of data collection:** January 2003 to not stated | **Outcome:** Incident visual field damage<br>**Method of measurement:** Standard automated perimetry with 24-2 Swedish Interactive Threshold Algorithm. Visual fields were defined as abnormal if pattern standard deviation was ≤ 5% and/or glaucoma hemifield test was "outside normal limits." Eyes that developed a repeatable visual defect, defined as 3 consecutive abnormal tests, were defined as developed visual field damage. The development of damage was reviewed by an ophthalmologist to | **Prognostic factor:** Race<br>**Method of measurement:** Self-reported<br>**Prognostic factor:** Age<br>**Method of measurement:** Self-reported<br>**Prognostic factor:** IOP<br>**Method of measurement:** Not stated<br>**Prognostic factor:** Central corneal thickness | **Adjusted prognostic effect (95% confidence interval):**<br>African descent vs. European descent by IOP:<br>No significant association at IOP = 10 mmHg to 20 mmHg IOP 22 mmHg HR 2.03 (1.15 to 3.57)<br>IOP 24 mmHg HR 2.71 (1.39 to 5.29)<br>IOP 26 mmHg HR 3.61 (1.61 to 8.08)<br>Mean IOP of cohort (17.8 mmHg) HR 1.12 (0.66 to 1.90)<br>Age (per year) HR 1.02 (0.99 to 1.04)<br>Central corneal thickness (per 40 microns thinner) HR 1.18 (0.86 to 1.60) | Glaucoma suspects of African descent with higher mean IOP are associated with increased risk of visual field damage compared to European glaucoma suspects.<br>Lower baseline visual field mean deviation and increasing mean arterial pressure are associated with increased risk of visual field damage. |
|  | mmHg or a history of ocular hypotensive treatment.<br>Inclusion criteria: best-corrected VA of 20/40 or better, spherical refraction less than 5.00 D, cylinder correction less than 3.00 D, open angles by gonioscopy, African or European descent, classed as glaucoma suspect at baseline, at least 2 years follow-up, at least 4 good quality visual field results.<br>Exclusion criteria: Coexisting ocular trauma, retinal disease, uveitis, non-glaucomatous optic disc neuropathy, or other diseases possibly affecting the visual field, evidence of consecutive repeatable visual field damage at baseline.<br>**Quality rating:** QUIPS RoB assessment<br>1. Study participation: Low risk of bias.<br>2. Study attrition: Low risk of bias.<br>3. Prognostic factor measurement: Low risk of bias.<br>4. Outcome measurement: Low risk of bias.<br>5. Adjustment for other |  | confirm that the damage was glaucomatous, and the location of damage was consistent on all 3 visual fields. | **Method of measurement:**<br>Not stated<br>**Prognostic factor:** SER<br>**Method of measurement:**<br>Not stated<br>**Prognostic factor:** Disc area, vertical cup-disc ratio<br>**Method of measurement:**<br>Clinician assessment of stereo-photographs<br>**Prognostic factor:** Baseline visual field mean deviation<br>**Method of measurement:**<br>Standard automated perimetry with 24-2 Swedish Interactive Threshold Algorithm.<br>**Prognostic factor:** Arterial pressure<br>**Method of measurement:**<br>Blood pressure | Lower SER (per D greater) HR 1.11 (0.84 to 1.34)<br>Disc area (per 0.4 mm<sup>2</sup> increase) HR 1.06 (0.84 to 1.34)<br>Vertical cup-disc ratio (per 0.1 increase) HR 1.25 (0.99 to 1.57)<br>Baseline visual field mean deviation (per 0.1 dB decrease) HR 1.04 (1.02 to 1.06)<br>Mean IOP (per 1 mmHg increase) HR 0.97 (0.92 to 1.03)<br>Mean arterial pressure (per 1 mmHg increase) HR 1.03 (1.00 to 1.06)<br>**Modelling method of analysis:** Multivariable Cox proportional hazards model<br>**Adjustment factors used:** Race, age, central corneal thickness, SER, disc area, baseline stereophotograph-based vertical cup-disc ratio, baseline visual field mean deviation, mean IOP during follow-up, mean arterial pressure, and a mean IOP*mean IOP interaction term and a race*mean IOP interaction term |  |
|  | prognostic factors: Low risk of bias.<br>6. Statistical analysis and reporting: Low risk of bias. |  |  | measured using DINAMAP@ PRO Monitor Model 100. Mean arterial pressure = (2/3) diastolic pressure + (1/3) systolic pressure. |  |  |
| **Marcus et al. (2012)**<br>**(The Netherlands)** | **Study Design:** Prospective, population-based cohort study<br>**Eligibility criteria/recruitment methods:** Part of the Rotterdam Study examining age-related disorders of individuals aged 55 year and older from a district in Rotterdam. Data were used from participants who completed the baseline ophthalmic examination, did not have glaucoma at baseline and completed at least one follow-up examination. Cases with a history or signs of angle closure (gonioscopy was performed in all identified cases) or secondary glaucoma (except for steroid-induced glaucoma) were excluded.<br>**Quality rating:** QUIPS RoB assessment | **Sample size:** 3939 participants<br>**Participants:** Mean follow-up 9.8 years<br>Incident glaucoma: 108 participants<br>Mean (± SD) age 68.4 ± 7.1 years<br>49.1% female<br>No glaucoma: 3831 participants<br>Mean (± SD) age 65.7 ± 6.8 years<br>58.7% female<br>**Dates of data collection:** 1991 to 2006 | **Outcome:** Incident OAG<br>**Method of measurement:** An incident OAG case was defined as a participant with no glaucomatous visual field loss in both eyes at baseline and glaucomatous visual field loss in at least one eye at follow-up.<br>At each examination, three IOP measurements were taken on each eye and the median value of these three measurements was recorded. The visual field of each eye was screened using a 52-point supra-threshold test that covered the central visual field with a radius of 24° (Humphrey Field Analyser). Visual field loss was defined as non-response to a light stimulus of 6 dB above a threshold-related estimate of the hill of vision in at least three contiguous test points, or four including the blind spot. In | **Prognostic factor:** Corticosteroid use<br>**Method of measurement:** Data on corticosteroid prescriptions for all participants were obtained from seven fully automated pharmacies using a centralized computer network from 1 January 1991 onward. This included the product name, Anatomical Therapeutic Chemical (ATC) code, number of prescriptions and the date of first prescription. Corticosteroids were classified as ophthalmic steroids, inhaled steroids, nasal steroids, oral steroids and | **Adjusted prognostic effect (95% confidence interval):**<br>Ophthalmic steroid use OR 1.04 (0.66 to 1.65, p = 0.86)<br>Inhaled steroid use OR 0.79 (0.42 to 1.48, p = 0.46)<br>Nasal steroid use OR 1.26 (0.74 to 2.13, p = 0.40)<br>Oral steroid use OR 1.03 (0.65 to 1.64, p = 0.89)<br>Ointment steroid use OR 0.70 (0.47 to 1.05, p = 0.086)<br>Age (per year) OR 1.06 (1.04 to 1.09, p < 0.001)<br>Sex (female) OR 0.63 (0.43 to 0.93, p = 0.022)<br>Positive family history of glaucoma OR 2.24 (1.31 to 3.84, p = 0.003)<br>High myopia OR 2.22 (1.13 to 4.38, p = 0.021)<br>**Modelling method of analysis:** Multivariate logistic regression analyses | Corticosteroid use is not associated with OAG incidence in adults aged 55 years and older.<br>Age, female sex, positive family history of glaucoma, and high myopia are associated with increased risk of incident OAG in adults aged 55 years and older. |
|  | 1. Study participation: Low risk of bias. Prospective cohort<br>2. Study attrition: Moderate risk of bias. 3939/6630 (59%) eligible participants had follow-up data.<br>3. Prognostic factor measurement: Moderate risk of bias. Family history self-reported<br>4. Outcome measurement: Low risk of bias.<br>5. Adjustment for other prognostic factors: Low risk of bias.<br>6. Statistical analysis and reporting: Low risk of bias. |  | participants with reproducible abnormalities on supra-threshold testing, Goldmann perimetry (baseline and first follow-up) or full-threshold Humphrey Field Analyser 24-2 testing (second follow-up) was performed on both eyes. Visual field loss was considered to be glaucomatous visual field loss only if reproducible and after excluding all other possible causes. | steroid ointments. The number of prescriptions during follow-up was used as a proxy for cumulative dose. Usage before baseline was not considered because the onset of the automated collection of medication data started on 1 January 1991.<br>**Prognostic factor:** Age, sex, family history of glaucoma<br>**Method of measurement:** Not explicitly stated<br>**Prognostic factor:** High myopia<br>**Method of measurement:** Refraction during eye examination, no further details given. High myopia defined as SER < -4.00 D. | **Adjustment factors used:** Age, sex, positive family history of glaucoma, high myopia |  |
| Pasquale & Kang (2011) | **Study Design:** Prospective cohort study | **Sample size:** 79,440 participants | **Outcome:** Incident OAG<br>**Method of measurement:** | **Prognostic factor:** Age at menarche | **Adjusted prognostic effect (95% confidence interval):** Age at menarche: | Age at menarche older than 13 years is associated with increased risk of normal tension |
| (USA) | **Eligibility criteria/recruitment methods:** The Nurses' Health Study is an ongoing population-based cohort of registered female nurses. The Nurses' Health Study was established in 1976 when 121,700 United States women were invited to complete a questionnaire regarding lifestyle, health behaviour, and chronic diseases. Follow-up biennial questionnaires were used to update this data and report newly diagnosed medical conditions including glaucoma. For this study, the follow-up was from 1980 to 2006. Eligible participants contributed to the study if they reached age 40 years and if they reported having had an eye exam in the period at risk. Eligible participants contributed person-time in 2-year units from the return date of the first biennial questionnaire until the occurrence of a report of glaucoma, cancer, death, or loss to follow-up, or | **Participants:** 100% female<br>All were aged 40 or more years<br>**Dates of data collection:** 1980 to 2006 | Self-reported in questionnaires, then followed up by investigator review of medical records to confirm diagnosis. | **Method of measurement:** Self-reported in biennial questionnaires.<br>**Prognostic factor:** Reproductive duration<br>**Method of measurement:** Self-reported in biennial questionnaires. Taken as age at natural menopause minus age at menarche.<br>**Prognostic factor:** Oral contraceptive use<br>**Method of measurement:** Self-reported in biennial questionnaires. After 1984, oral contraceptive use was defined as ever / never.<br>**Prognostic factor:** Parity<br>**Method of measurement:** Self-reported in biennial questionnaires. | > 13 years and normal tension glaucoma RR 1.47 (1.01 to 2.13)<br>< 12 years (ref)<br>Reproductive duration:<br>< 36 years RR 0.93 (0.71 to 1.22)<br>36-38 years RR 0.94 (0.73 to 1.21)<br>39-40 years (ref)<br>≥ 41 years RR 0.96 (0.73 to 1.27)<br>Oral contraceptives, duration of use:<br>Ever used RR 1.14 (0.98 to 1.34)<br>< 2 years RR 1.10 (0.89 to 1.36)<br>2-4 years RR 1.04 (0.81 to 1.35)<br>5+ years RR 1.25 (1.02 to 1.53)<br>Never used (ref)<br>Oral contraceptives, time since discontinuing use:<br>≥ 25 years RR 1.13 (0.91 to 1.40)<br>20-24 years RR 1.06 (0.82 to 1.38)<br>15-19 years RR 1.20 (0.91 to 1.59)<br>< 10 years RR 1.39 (1.01 to 1.91)<br>Never used (ref)<br>Parity:<br>No children RR 0.85 (0.60 to 1.21) | glaucoma in women aged 40 years or older.<br>Five years or greater use of oral contraceptives and time since discontinuing use of oral contraceptives less than 10 years is associated with increased risk of OAG in women aged 40 years or older.<br>Reproductive duration and parity are not associated with OAG in women aged 40 years or older. |
|  | until 2006, whichever came first.<br>At each 2-year follow-up cycle, only women who indicated they received an eye exam in the previous 2 years were eligible to contribute person-time to the study.<br>**Quality rating:** QUIPS RoB assessment<br>1. Study participation: Moderate risk of bias. Prospective cohort. 79440 / 121700 had sufficient data to be included<br>2. Study attrition: Low risk of bias.<br>3. Prognostic factor measurement: Moderate risk of bias. Self reported via questionnaire<br>4. Outcome measurement: Moderate risk of bias. Self-reported glaucoma - but verified by review of medical records.<br>5. Adjustment for other prognostic factors: Low risk of bias.<br>6. Statistical analysis and reporting: Low risk of bias. |  |  |  | 1-2 children (ref)<br>3 children RR 1.08 (0.90 to 1.29)<br>4+ children RR 1.00 (0.84 to 1.19)<br>**Modelling method of analysis:** Cox proportional hazard analyses<br>**Adjustment factors used:** Age, time-interval at risk, family history of glaucoma, African ancestry, hypertension, diabetes, smoking status, alcohol intake, caffeine intake, BMI, physical activity, post-menopausal hormone use, oral contraceptive use, parity, age at menopause |  |
| **Stem et al. (2013)**<br>**(USA)** | **Study Design:** Retrospective, longitudinal cohort analysis | **Sample size:** 494,165 participants | **Outcome:** Incident CRVO<br>**Method of measurement:** Identified by reported ICD code. To be counted as an incident | **Prognostic factor:** sex, ethnicity, household net worth, metabolic | **Adjusted prognostic effect (95% confidence interval):**<br>Sex (female) HR 0.75 (0.66 to 0.85, p < 0.0001) | Black ethnicity, lower household net worth, having hypertension (alone or in combination with diabetes, |
| | **Eligibility criteria/recruitment methods:** Investigators used the i3 InVision Data Mart database (Ingenix, Eden Prairie, MN), which contains detailed records of all beneficiaries in a managed care network throughout the United States. The dataset contains all individuals with $\geq 1$ International Classification of Diseases, 9th Revision, Clinical Modification (ICD-9-CM) codes for eye-related diagnoses; $\geq 1$ Current Procedural Terminology codes for any eye-related visits, diagnostic, or therapeutic procedures; or any other claim submitted by an ophthalmologist or optometrist from January 1 2001 to December 31 2009. The investigators had access to all medical claims for ocular and non-ocular conditions and sociodemographic information. Individuals were included in the analysis if they met the following criteria: continuous enrolment in the medical plan for at least 2 years, $\geq 2$ visits to an eye care provider | **Participants:** 1302 (0.26%) with newly diagnosed CRVO<br>Mean (± SD) age 65.7 ± 8.1 years (those without CRVO diagnosis), 69.9 ± 8.4 (those with CRVO diagnosis)<br>41.7% male<br>Ethnicity: 79.7% White, 4.9% Black, 3.4% Latino, 1.6% Asian-American, 0.7% other, 9.7% unknown<br>**Dates of data collection:** January 2001 to December 2009 | case of CRVO, individuals must have had at least 1 eye care visit during their first 2 years in the plan (with no documented diagnosis of CRVO) and then must have been diagnosed with CRVO at a subsequent visit after the index date (2 years after entry into the plan). Beneficiaries were identified with a CRVO if they had $\geq 1$ billing records with the ICD-9-CM code 362.35. | syndrome components, vascular disease, ophthalmic disease<br>**Method of measurement:** Data retrieved from i3 InVision Data Mart database. | Ethnicity:<br>Black HR 1.58 (1.25 to 1.99, $p < 0.0001$)<br>Asian-American HR 0.75 (0.43 to 1.30, $p = 0.31$)<br>White (ref)<br>Household net worth:<br>> \$500,000 HR 0.73 (0.56 to 0.96, $p = 0.02$)<br>< \$25,000 (ref)<br>Metabolic syndrome:<br>Hypertension HR 1.66 (1.14 to 2.42, $p = 0.01$)<br>Hypertension and diabetes HR 1.82 (1.15 to 2.89, $p = 0.01$)<br>Hypertension and hyperlipidaemia HR 1.46 (1.04 to 2.05, $p = 0.03$)<br>Hypertension, hyperlipidaemia and diabetes HR 1.58 (1.11 to 2.23, $p = 0.01$)<br>No diabetes, hypertension or hyperlipidaemia (ref)<br>Vascular disease:<br>Cerebrovascular accident HR 1.44 (1.23 to 1.68, $p < 0.0001$)<br>Peripheral artery disease HR 1.15 (1.00 to 1.33, $p = 0.05$)<br>Myocardial infarction HR 0.72 (0.57 to 0.92, $p = 0.01$)<br>Hypercoagulable state HR 2.45 (1.40 to 4.28, $p = 0.002$)<br>Ophthalmic disease:<br>OAG HR 1.50 (1.30 to 1.72, $p < 0.0001$) | hyperlipidaemia, or both), previous cerebrovascular accident, having peripheral arterial disease, hypercoagulable state, OAG, AMD, and cataract are all associated with higher risk for incident CRVO in adults aged 55 years and over.<br>Previous myocardial infarction is associated with lower risk of incident CRVO in adults aged 55 years and over. |
|  | (ophthalmologist or optometrist), and age ≥ 55 years. Individuals were excluded if they received a diagnosis of CRVO during the first 2 years they were enrolled in the plan to exclude non-incident cases.<br>**Quality rating:** QUIPS RoB assessment<br>1. Study participation: Low risk of bias.<br>2. Study attrition: Low risk of bias.<br>3. Prognostic factor measurement: Low risk of bias.<br>4. Outcome measurement: Low risk of bias.<br>5. Adjustment for other prognostic factors: Low risk of bias.<br>6. Statistical analysis and reporting: Low risk of bias. |  |  |  | AMD HR 1.50 (1.31 to 1.72, $p < 0.0001$)<br>Cataract HR 1.24 (1.08 to 1.42, $p = 0.003$)<br>**Modelling method of analysis:** Multivariate Cox regression analysis<br>**Adjustment factors used:** age, sex, ethnicity, education level, household net worth, region of residence, ocular co-morbidities, systemic co-morbidities, Charlson co-morbidity index |  |
| **Stingl et al. (2023)**<br>**(Germany)** | **Study Design:** Prospective, population-based cohort study<br>**Eligibility criteria/recruitment methods:** Random sampling of residents of the State of Rhine-Palatine by the regional registration office. | **Sample size:** 10,175 participants (9978 right eyes, 9952 left eyes)<br>**Participants:** Mean (± SD) age $53.5 \pm 10.5$ years, range 35-74 years<br>48.5% female<br>**Dates of data collection:** 2007 to 2017 | **Outcome:** 5-year change in refractive error<br>**Method of measurement:** Refractive error measurement was conducted without cycloplegia. Refractive values were measured in spherical and cylindrical dioptres (D), cylindrical power was indicated in negative sign | **Prognostic factor:** Sex, age, smoker status, education, occupation<br>**Method of measurement:** Method of measurement/data collection not explicitly stated. | **Adjusted prognostic effect (95% confidence interval):**<br>Myopic change at 5 years:<br>Sex (female) OR 1.49 (1.28 to 1.73, $p < 0.001$)<br>Age (per year) OR 0.52 (0.49 to 0.55, $p < 0.001$)<br>Baseline SER (per dioptre) OR 0.89 (0.87 to 0.91, $p < 0.001$)<br>Myopic change no significant association with cardiovascular parameters, | Female sex, younger age and baseline myopic SER are associated with increased risk of having a myopic shift in refractive error at 5 years in adults aged 35 to 74 years. Increasing age and being a smoker are associated with an increased risk of a hyperopic shift in refractive error at 5 years |
|  | Inclusion criteria: mental and physical ability to visit the study centre and to pass through the examinations, sufficient knowledge of the German language.<br>All study participants with objective refraction measurement at both baseline and 5-year follow-up examinations were included.<br>Exclusion criteria: previous corneal or cataract surgery, aphakia.<br>**Quality rating:** QUIPS RoB assessment<br>1. Study participation: Low risk of bias.<br>Prospective cohort<br>2. Study attrition: Moderate risk of bias.<br>12423/15010 (83%) of eligible participants had follow-up data<br>3. Prognostic factor measurement: Low risk of bias.<br>4. Outcome measurement: Low risk of bias.<br>5. Adjustment for other prognostic factors: Moderate risk of bias.<br>Family history of refractive error and ethnicity not included<br>6. Statistical analysis and |  | convention. SER was computed as $SER = \text{sphere} + 0.5 \times \text{cylinder}$.<br>No refractive change was defined as $-0.50$ to $+0.50$ D change in SER, myopic shift as $< -0.50$ D and hyperopic shift as $> +0.50$ D. | **Prognostic factor:** Baseline SER<br>**Method of measurement:** As for outcome.<br>**Prognostic factor:** IOP<br>**Method of measurement:** Non-contact tonometry (Nidek NT-2000)<br>**Prognostic factor:** Lens opacity<br>**Method of measurement:** Slit lamp examination<br>**Prognostic factor:** Physical activity<br>**Method of measurement:** Completion of the Short Questionnaire to Assess Health-enhancing physical activity<br>**Prognostic factor:** Cardiovascular parameters<br>**Method of measurement:** | physical activity, smoking history, IOP, lens opacity, education, or occupation.<br>Hyperopic change at 5 years: Age (per year) OR 1.62 (1.52 to 1.72, $p < 0.001$)<br>Smoker OR 1.31 (1.14 to 1.50, $p < 0.001$)<br>Hyperopic change no significant association with sex, cardiovascular parameters, physical activity, baseline SER, IOP, lens opacity, education, or occupation.<br>**Modelling method of analysis:** Multivariable logistic regression analyses<br>**Adjustment factors used:** Sex, age, SER, IOP, presence of cataract, cardiovascular parameters, BMI, physical activity, smoking, education, occupation | in adults aged 35 to 74 years.<br>Cardiovascular parameters, physical activity, IOP, lens opacity, education, and occupation were not associated with increased risk of either myopic or hyperopic shift at 5 years in adults aged 35 to 74 years. |
|  | reporting: Moderate risk of bias. Unclear whether strategy for model building is appropriate and is based on a conceptual framework or model |  |  | Laboratory measurements<br>**Prognostic factor:** BMI<br>**Method of measurement:** Height and weight measured and BMI calculated as weight/height<sup>2</sup> |  |  |
| **Wright et al. (2020)**<br>(UK) | **Study Design:** Retrospective cohort analysis<br>**Eligibility criteria/recruitment methods:** Information on attendance at routine eye examinations was drawn from the Family Practitioner Services Ophthalmic Database (managed by the Business Services Organisation, NI Department of Health), an administrative database used to manage payment to service providers. Records of eye examinations of those aged ≥ 60 years conducted during a 5-year period (October 2009 to September 2014 inclusive) were extracted. The cohort consisted of all community-dwelling | **Sample size:** 132,046 participants, 444,045 eye examinations, 311,999 examination intervals<br>**Participants:** Aged 60-69 43.2% male<br>Aged ≥ 70 40.7% male. No other details given.<br>**Dates of data collection:** October 2009 to September 2014 | **Outcome:** Referral to a GP<br>**Method of measurement:** Data were extracted from the Family Practitioner Services Ophthalmic Database. | **Prognostic factor:** Delayed attendance at eye examination<br>**Method of measurement:** Data were extracted from the Family Practitioner Services Ophthalmic Database. Examination intervals were split into three categories based on recommended intervals for each age group. The first category 'on-time' consisted of intervals conforming to recommendations (24 months for those aged 60–69, 12 months for those aged ≥ 70). Longer intervals (> 24 and > 12 | **Adjusted prognostic effect (95% confidence interval):**<br>Aged 60-69:<br>Delayed eye exam attendance OR 1.30 (1.04 to 1.61)<br>Early eye exam attendance OR 2.86 (2.36 to 3.46)<br>On-time eye exam attendance (ref)<br>Aged ≥ 70:<br>Delayed eye exam attendance OR 1.07 (1.01 to 1.13)<br>Early eye exam attendance OR 2.72 (2.58 to 2.87)<br>On-time eye exam attendance (ref)<br>**Modelling method of analysis:** Multivariable logistic regression<br>**Adjustment factors used:** sex, religion, eligibility for NHS sight test on health grounds, eligibility for NHS sight test on income grounds, general health, household structure, tenure, household cars, practice density, drive time, | Delayed attendance for eye examinations is associated with increased risk of requiring a GP referral for adults aged 60 years and older. Early attendance is also associated with increased risk of referral for adults aged 60 years or older, though this is driven by early attendance usually being due to symptomatic problems or early recall suggested by the optometrist. |
|  | respondents to the 2011 Census aged $\geq 60$ years at the beginning of the study period that had attended at least two free eye examinations during the period. A longitudinal sequence of eye examinations was constructed for each individual and from these, the analysis dataset of 311,999 examination intervals was calculated.<br>**Quality rating:** QUIPS RoB assessment<br>1. Study participation: Low risk of bias. Cohort<br>2. Study attrition: Low risk of bias.<br>3. Prognostic factor measurement: Low risk of bias.<br>4. Outcome measurement: Low risk of bias.<br>5. Adjustment for other prognostic factors: Low risk of bias.<br>6. Statistical analysis and reporting: Low risk of bias |  |  | months for the younger and older groups respectively) were classified as 'delayed attendance'. The third category 'early recall' consisted of intervals shorter than recommended. Early recall may occur when an individual returns to the optometrist with visual symptoms or at the request of the optometrist to monitor an ocular condition that does not warrant immediate GP referral. Classifications were based on calendar months. | highest qualification, carer status, household adaptations for visual difficulties, area income deprivation |  |

**Abbreviations:** AL: axial length; AMD: age-related macular degeneration; BMI: body mass index; CRVO: central retinal vein occlusion; D: dioptre; dB: decibel; EG: exfoliation glaucoma; EGS: exfoliation glaucoma suspect; HL: hearing level; HR: hazard ratio; IQR: interquartile range; logMAR: logarithm of the minimum angle of resolution; OAG: open-angle glaucoma; OHT: ocular hypertension; OR: odds ratio; QUIPS: Quality In Prognosis Studies tool; RBC: red blood cell; RoB: risk of bias; SD: standard deviation; SER: spherical equivalent refraction; VA: visual acuity; VI: visual impairment

### 6.3 Quality appraisal

**Table 10:**
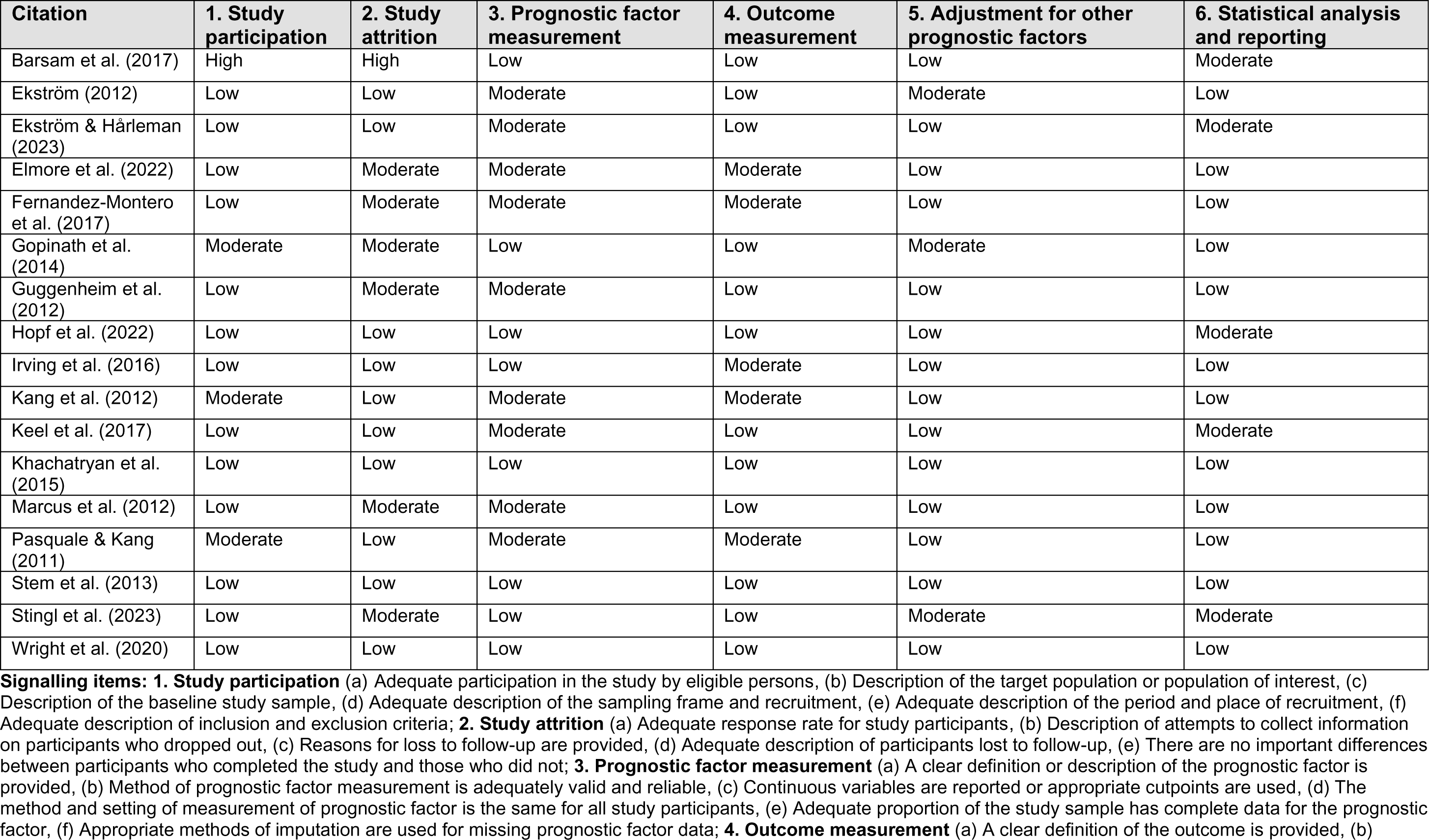
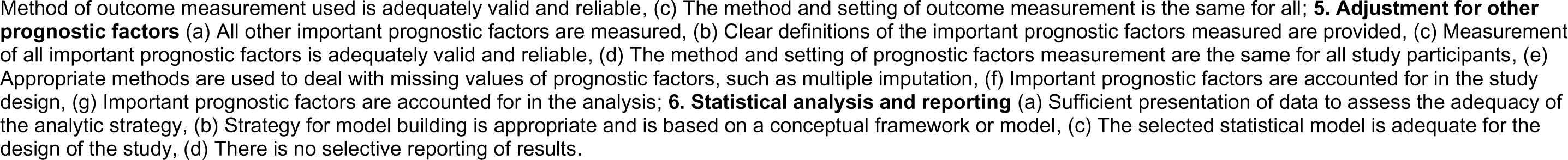
Quality in Prognostic factor Studies (QUIPS) tool for included primary studies.

| Citation | 1. Study participation | 2. Study attrition | 3. Prognostic factor measurement | 4. Outcome measurement | 5. Adjustment for other prognostic factors | 6. Statistical analysis and reporting |
| --- | --- | --- | --- | --- | --- | --- |
| Barsam et al. (2017) | High | High | Low | Low | Low | Moderate |
| Ekström (2012) | Low | Low | Moderate | Low | Moderate | Low |
| Ekström & Hårleman (2023) | Low | Low | Moderate | Low | Low | Moderate |
| Elmore et al. (2022) | Low | Moderate | Moderate | Moderate | Low | Low |
| Fernandez-Montero et al. (2017) | Low | Moderate | Moderate | Moderate | Low | Low |
| Gopinath et al. (2014) | Moderate | Moderate | Low | Low | Moderate | Low |
| Guggenheim et al. (2012) | Low | Moderate | Moderate | Low | Low | Low |
| Hopf et al. (2022) | Low | Low | Low | Low | Low | Moderate |
| Irving et al. (2016) | Low | Low | Low | Moderate | Low | Low |
| Kang et al. (2012) | Moderate | Low | Moderate | Moderate | Low | Low |
| Keel et al. (2017) | Low | Low | Moderate | Low | Low | Moderate |
| Khachatryan et al. (2015) | Low | Low | Low | Low | Low | Low |
| Marcus et al. (2012) | Low | Moderate | Moderate | Low | Low | Low |
| Pasquale & Kang (2011) | Moderate | Low | Moderate | Moderate | Low | Low |
| Stem et al. (2013) | Low | Low | Low | Low | Low | Low |
| Stingl et al. (2023) | Low | Moderate | Low | Low | Moderate | Moderate |
| Wright et al. (2020) | Low | Low | Low | Low | Low | Low |

**Signalling items:** **1. Study participation** (a) Adequate participation in the study by eligible persons, (b) Description of the target population or population of interest, (c) Description of the baseline study sample, (d) Adequate description of the sampling frame and recruitment, (e) Adequate description of the period and place of recruitment, (f) Adequate description of inclusion and exclusion criteria; **2. Study attrition** (a) Adequate response rate for study participants, (b) Description of attempts to collect information on participants who dropped out, (c) Reasons for loss to follow-up are provided, (d) Adequate description of participants lost to follow-up, (e) There are no important differences between participants who completed the study and those who did not; **3. Prognostic factor measurement** (a) A clear definition or description of the prognostic factor is provided, (b) Method of prognostic factor measurement is adequately valid and reliable, (c) Continuous variables are reported or appropriate cutpoints are used, (d) The method and setting of measurement of prognostic factor is the same for all study participants, (e) Adequate proportion of the study sample has complete data for the prognostic factor, (f) Appropriate methods of imputation are used for missing prognostic factor data; **4. Outcome measurement** (a) A clear definition of the outcome is provided, (b)
Method of outcome measurement used is adequately valid and reliable, (c) The method and setting of outcome measurement is the same for all; **5. Adjustment for other prognostic factors** (a) All other important prognostic factors are measured, (b) Clear definitions of the important prognostic factors measured are provided, (c) Measurement of all important prognostic factors is adequately valid and reliable, (d) The method and setting of prognostic factors measurement are the same for all study participants, (e) Appropriate methods are used to deal with missing values of prognostic factors, such as multiple imputation, (f) Important prognostic factors are accounted for in the study design, (g) Important prognostic factors are accounted for in the analysis; **6. Statistical analysis and reporting** (a) Sufficient presentation of data to assess the adequacy of the analytic strategy, (b) Strategy for model building is appropriate and is based on a conceptual framework or model, (c) The selected statistical model is adequate for the design of the study, (d) There is no selective reporting of results.

**Table 11:**
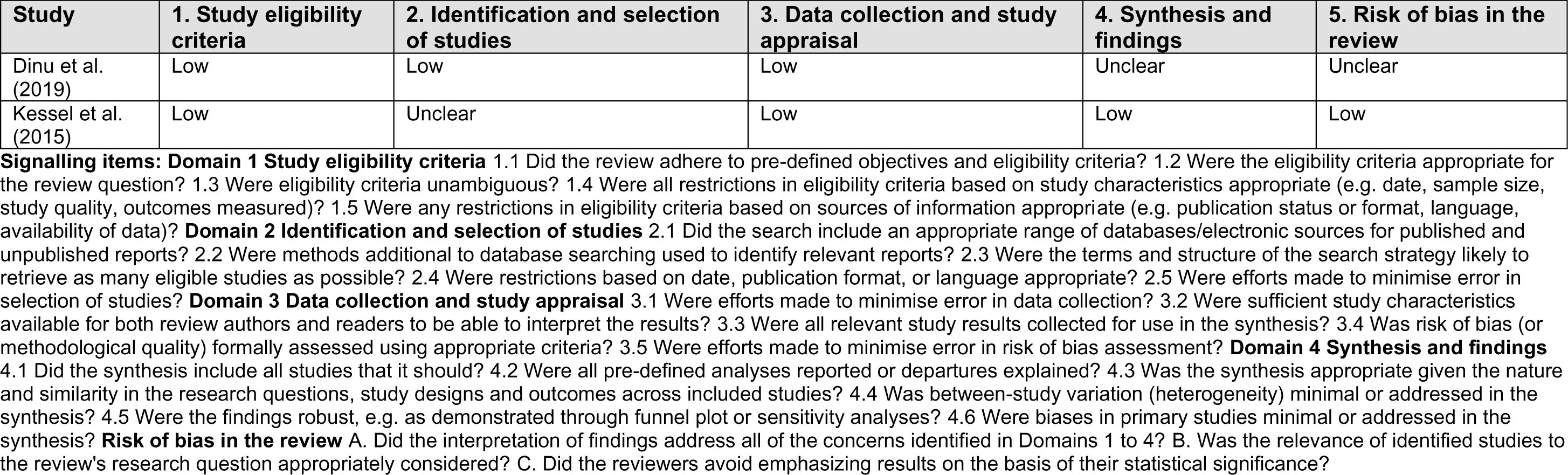
Risk of Bias in Systematic reviews (ROBIS) tool for included secondary studies.

### 6.4 Information available on request

The protocol, search strategies, and excluded studies for this rapid review are available on request.

## 7. ADDITIONAL INFORMATION

### 7.1 Conflicts of interest

The authors declare they have no conflicts of interest to report.

## 7.2 Acknowledgements

The authors would like to thank Rebecca Bartlett, Mike George, David O’Sullivan, Sarah O’Sullivan-Adams, Tim Morgan, Robert Hall and Rashmi Kumar for their contributions during stakeholder meetings in guiding the focus of the review and interpretation of findings.

## 8. APPENDIX

## APPENDIX 1: Search Strategies

**Searches for Secondary Research Medline Search Strategy**

Ovid MEDLINE(R) ALL <1946 to August 03, 2023>

Conducted 03.08.2023

| # | Search Query | Results |
| --- | --- | --- |
| 1 | exp Prognosis/ | 1922657 |
| 2 | exp Incidence/ | 301423 |
| 3 | exp Risk Assessment/ | 310857 |
| 4 | exp Decision Support Techniques/ | 82250 |
| 5 | (prognos* or prevention or progress* or diagnos* or detect* or prevalence or incidence or rate*).tw. | 10451982 |
| 6 | (risk* adj2 (assess* or factor*)).tw. | 878756 |
| 7 | ((first or initial) adj (episode* or detection)).tw. | 21793 |
| 8 | 1 or 2 or 3 or 4 or 5 or 6 or 7 | 11718920 |
| 9 | exp Vision Screening/ | 2467 |
| 10 | exp Vision Tests/ | 117699 |
| 11 | exp Mass Screening/ | 144061 |
| 12 | ((eye* or sight or vision or visual) adj1 (test* or exam* or screen* or follow-up)).tw | 17962 |
| 13 | (asymptomatic adj2 (test* or exam* or screen*)).tw. | 4556 |
| 14 | (routine adj2 (test* or exam* or screen*)).tw. | 43911 |
| 15 | ((eye* or sight or vision or visual or screening) adj3 (frequenc* or interval* or recall*)).tw. | 8138 |
| 16 | 9 or 10 or 11 or 12 or 13 or 14 or 15 | 318610 |
| 17 | exp Eye Diseases/ | 637918 |
| 18 | exp Vision, Ocular/ | 29938 |
| 19 | exp Visual Acuity/ | 92659 |
| 20 | (ametrop* or emmetrop* or glaucoma or diabetic retinopathy or refractive error or macular degeneration or cataract* or presbyop* or amblyop* or myop* or hyperop* or hypermetrop* or astigmat* or anisometrop* or vision impairment* or vision loss* or vis* acuity).tw. | 255933 |
| 21 | exp Cataract/di, dg, pc [Diagnosis, Diagnostic Imaging, Prevention & Control] | 4774 |
| 22 | exp Glaucoma/di, dg, pc [Diagnosis, Diagnostic Imaging, Prevention & Control] | 15607 |
| 23 | exp Macular Degeneration/di, dg, pc [Diagnosis, Diagnostic Imaging, Prevention & Control] | 9999 |
| 24 | exp Diabetic Retinopathy/di, dg, pc [Diagnosis, Diagnostic Imaging, Prevention & Control] | 9211 |
| 25 | exp Refractive Errors/di, dg, pc [Diagnosis, Diagnostic Imaging, Prevention & Control] | 7806 |
| 26 | 17 or 18 or 19 or 20 or 21 or 22 or 23 or 24 or 25 | 757960 |
| 27 | (UK or United kingdom or England or Wales or Scotland or Northern Ireland or Ireland or Australia or Canada or New Zealand or USA or United States or Austria or Finland or Germany or Malta or Netherlands or Norway or Spain or Sweden or Switzerland).tw,cp. | 29980939 |
| 28 | exp "Systematic Review"/ | 234551 |
| 29 | exp Meta-Analysis/ | 184949 |
| 30 | exp Systematic Reviews as Topic/ | 10951 |
| 31 | exp Meta-Analysis as Topic/ | 27324 |
| 32 | (systematic review* or meta analys#s or review*).pt. | 3356402 |
| 33 | (systematic adj2 (review* or overview*)).ti,ab,kf. | 310829 |
| 34 | (quantitative adj2 (review* or overview* or synthes*)).ti,ab,kf. | 6829 |
| 35 | (meta-analy* or metaanaly* or meta-synthes#s or metasynthes#s).tw. | 277814 |
| 36 | rapid review*.tw. | 1967 |
| 37 | 28 or 29 or 30 or 31 or 32 or 33 or 34 or 35 or 36 | 3455183 |
| 38 | 8 and 16 and 26 and 27 and 37 | 3178 |
| 39 | limit 38 to (english language and yr="2009-Current") | **1967** |

**EMBASE Search Strategy**

Conducted 03.08.2023

EMBASE Search Strategy Conducted 03.08.2023
| # | Search Query | Results |
| --- | --- | --- |
| 1 | exp Prognosis/ | 912238 |
| 2 | exp Incidence/ | 670315 |
| 3 | exp Risk Assessment/ | 737723 |
| 4 | exp Decision Support Techniques/ | 34339 |
| 5 | (prognos* or prevention or progress* or diagnos* or detect* or prevalence or incidence or rate*).tw. | 13884935 |
| 6 | (risk* adj2 (assess* or factor*)).tw. | 1278642 |
| 7 | ((first or initial) adj (episode* or detection)).tw. | 33689 |
| 8 | 1 or 2 or 3 or 4 or 5 or 6 or 7 | 14827598 |
| 9 | exp Vision Screening/ | 43654 |
| 10 | exp Vision Tests/ | 43654 |
| 11 | exp Mass Screening/ | 307655 |
| 12 | ((eye* or sight or vision or visual) adj1 (test* or exam* or screen* or follow-up)).tw | 24336 |
| 13 | (asymptomatic adj2 (test* or exam* or screen*)).tw. | 6538 |
| 14 | (routine adj2 (test* or exam* or screen*)).tw. | 66065 |
| 15 | ((eye* or sight or vision or visual or screening) adj3 (frequenc* or interval* or recall*)).tw. | 11434 |
| 16 | 9 or 10 or 11 or 12 or 13 or 14 or 15 | 442099 |
| 17 | exp Eye Diseases/ | 1095511 |
| 18 | exp Vision, Ocular/ | 316640 |
| 19 | exp Visual Acuity/ | 154663 |
| 20 | (ametrop* or emmetrop* or glaucoma or diabetic retinopathy or refractive error or macular degeneration or cataract* or presbyop* or amblyop* or myop* or hyperop* or hypermetrop* or astigmat* or anisometrop* or vision impairment* or vision loss* or vis* acuity).tw. | 327995 |
| 21 | exp Cataract/di, dg, pc [Diagnosis, Diagnostic Imaging, Prevention & Control] | 5974 |
| 22 | exp Glaucoma/di, dg, pc [Diagnosis, Diagnostic Imaging, Prevention & Control] | 14182 |
| 23 | exp Macular Degeneration/di, dg, pc [Diagnosis, Diagnostic Imaging, Prevention & Control] | 2396 |
| 24 | exp Diabetic Retinopathy/di, dg, pc [Diagnosis, Diagnostic Imaging, Prevention & Control] | 8275 |
| 25 | exp Refractive Errors/di, dg, pc [Diagnosis, Diagnostic Imaging, Prevention & Control] | 6537 |
| 26 | 17 or 18 or 19 or 20 or 21 or 22 or 23 or 24 or 25 | 1297500 |
| 27 | (UK or United kingdom or England or Wales or Scotland or Northern Ireland or Ireland or Australia or Canada or New Zealand or USA or United States or Austria or Finland or Germany or Malta or Netherlands or Norway or Spain or Sweden or Switzerland).tw,cp. | 30453772 |
| 28 | exp "Systematic Review"/ | 424400 |
| 29 | exp Meta-Analysis/ | 288688 |
| 30 | exp Systematic Reviews as Topic/ | 32284 |
| 31 | exp Meta-Analysis as Topic/ | 53131 |
| 32 | (systematic review* or meta analys#s or review*).pt. | 3104591 |
| 33 | (systematic adj2 (review* or overview*)).ti,ab,kf. | 379724 |
| 34 | (quantitative adj2 (review* or overview* or synthes*)).ti,ab,kf. | 7763 |
| 35 | (meta-analy* or metaanaly* or meta-synthes#s or metasynthes#s).tw. | 353655 |
| 36 | rapid review*.tw. | 2326 |
| 37 | 28 or 29 or 30 or 31 or 32 or 33 or 34 or 35 or 36 | 3551601 |
| 38 | 8 and 16 and 26 and 27 and 37 | 3030 |
| 39 | limit 38 to (english language and yr="2009 -Current") | **2113** |

**CINAHL Search Strategy**

Conducted 03.08.2023

CINAHL Search Strategy Conducted 03.08.2023
| # | Search Query | Results |
| --- | --- | --- |
| 1 | (MH "Prognosis+") | 550866 |
| 2 | (MM "Incidence") | 1707 |
| 3 | (MM "Risk Assessment") | 62988 |
| 4 | TX (prognos* or prevention or progress* or diagnos* or detect* or prevalence or incidence or rate*) | 3479872 |
| 5 | AB (risk* N2 (assess* or factor*)) | 239041 |
| 6 | TX ((first or initial) N1 (episode* or detection)) | 12,663 |
| 7 | S1 OR S2 OR S3 OR S4 OR S5 OR S6 | 3727296 |
| 8 | (MM "Vision Screening") | 822 |
| 9 | (MH "Vision Tests+") | 7549 |
| 10 | TX ((eye* or sight or vision or visual) N1 (test* or exam* or screen* or follow-up)) | 19149 |
| 11 | TX (asymptomatic N2 (test* or exam* or screen*)) | 3751 |
| 12 | TX (routine N2 (test* or exam* or screen*)) | 33516 |
| 13 | AB ((eye* or sight or vision or visual or screening) N3 (frequenc* or interval* or recall*)) | 2647 |
| 14 | S8 OR S9 OR S10 OR S11 OR S12 OR S13 | 61140 |
| 15 | (MH "Eye Diseases+") | 103998 |
| 16 | (MH "Diagnosis, Eye+") | 23008 |
| 17 | (MM "Vision, Subnormal") | 1133 |
| 18 | (MM "Visual Acuity") | 3891 |
| 19 | AB (ametrop* or emmetrop* or glaucoma or diabetic retinopathy or refractive error or macular degeneration or cataract* or presbyop* or amblyop* or myop* or hyperop* or hypermetrop* or astigmat* or anisometrop* or vision impairment* or vision loss* or vis* acuity) | 33038 |
| 20 | (MM "Cataract/DI/PC/PR") | 459 |
| 21 | (MH "Glaucoma+/DI/PC/PR") | 2292 |
| 22 | (MH "Macular Degeneration+/DI/PC/PR") | 2167 |
| 23 | (MM "Diabetic Retinopathy/DI/PC/PR") | 1427 |
| 24 | (MH "Refractive Errors+/DI/PC/PR") | 1472 |
| 25 | S15 OR S16 OR S17 OR S18 OR S19 OR S20 OR S21 OR S22 OR S23 OR S24 | 124965 |
| 26 | TX (UK or United kingdom or England or Wales or Scotland or Northern Ireland or Ireland or Australia or Canada or New Zealand or USA or United States or Austria or Finland or Germany or Malta or Netherlands or Norway or Spain or Sweden or Switzerland) | 7581371 |
| 27 | (MM "Systematic Review") | 1514 |
| 28 | (MM "Meta Analysis") | 1779 |
| 29 | PT (systematic review* or meta analys#s or review*) | 479918 |
| 30 | AB (systematic N2 (review* or overview*)) | 84071 |
| 31 | AB (quantitative N2 (review* or overview* or synthes*)) | 2434 |
| 32 | AB (meta-analy* or metaanaly* or meta-synthes#s or metasynthes#s) | 72993 |
| 33 | AB rapid review* | 1295 |
| 34 | S27 OR S28 OR S29 OR S30 OR S31 OR S32 OR S33 | 523656 |
| 35 | S7 AND S14 AND S25 AND S26 AND S34<br>Limiters - Published Date: 20090101-20230831; Language: English | 408 |

**Epistemonikos**

| Search Query | Results |
| --- | --- |
| (advanced_title_en:(advanced_title_en:((advanced_title_en:(vision screening) OR advanced_abstract_en:(vision screening))) OR advanced_title_en:(vision test) OR advanced_title_en:(eye test) OR advanced_title_en:(eye sight) OR advanced_title_en:(visual acuity)) OR advanced_abstract_en:(advanced_title_en:((advanced_title_en:(vision screening) OR advanced_abstract_en:(vision screening))) OR advanced_title_en:(vision test) OR advanced_title_en:(eye test) OR advanced_title_en:(eye sight) OR advanced_title_en:(visual acuity))) AND (advanced_title_en:((prognos* OR prevention OR progress* OR diagnos* OR detect* OR prevalence OR incidence OR rate*)) OR advanced_abstract_en:((prognos* OR prevention OR progress* OR diagnos* OR detect* OR prevalence OR incidence OR rate*))) [Filters: classification=systematic-review, protocol=no, min_year=2009, max_year=2023] | 148 |

| Database | Results |
| --- | --- |
| Medline (Ovid) | 1967 |
| EMBASE (Ovid) | 2113 |
| CINAHL (EBSCO) | 408 |
| Epistemonikos | 148 |
| Cochrane | 5 |
| TOTAL | 4641 |

**Searches for Primary Research Medline Search Strategy**

Ovid MEDLINE(R) ALL <1946 to August 21, 2023>

Conducted 21.08.2023

| # | Search Query | Results |
| --- | --- | --- |
| 1 | exp Prognosis/ | 1925629 |
| 2 | Incidence/ | 301723 |
| 3 | Risk Assessment/ | 306886 |
| 4 | Decision Support Techniques/ | 22494 |
| 5 | (prognos* or prevention or progress* or diagnos* or detect* or prevalence or incidence or rate*).tw. | 10479336 |
| 6 | (risk* adj2 (assess* or factor*)).tw. | 881915 |
| 7 | ((first or initial) adj (episode* or detection)).tw. | 21854 |
| 8 | 1 or 2 or 3 or 4 or 5 or 6 or 7 | 11702337 |
| 9 | exp Vision Screening/ | 2468 |
| 10 | exp Vision Tests/ | 117787 |
| 11 | exp Mass Screening/ | 144174 |
| 12 | ((eye* or sight or vision or visual) adj1 (test* or exam* or screen* or follow-up)).ti,ab. | 18010 |
| 13 | (asymptomatic adj2 (test* or exam* or screen*)).tw. | 4567 |
| 14 | (routine adj2 (test* or exam* or screen*)).tw. | 44026 |
| 15 | ((eye* or sight or vision or visual or screening) adj3 (frequenc* or interval* or recall*)).ti,ab. | 8155 |
| 16 | 9 or 10 or 11 or 12 or 13 or 14 or 15 | 318989 |
| 17 | Eye Diseases/ | 38697 |
| 18 | Vision, Ocular/ | 27018 |
| 19 | Visual Acuity/ | 85126 |
| 20 | (ametrop* or emmetrop* or glaucoma or diabetic retinopathy or refractive error or macular degeneration or cataract* or presbyop* or amblyop* or myop* or hyperop* or hypermetrop* or astigmat* or anisometrop* or vision impairment* or vision loss* or vis* acuity).tw. | 256482 |
| 21 | Cataract/di, dg, pc [Diagnosis, Diagnostic Imaging, Prevention & Control] | 4607 |
| 22 | Glaucoma/di, dg, pc [Diagnosis, Diagnostic Imaging, Prevention & Control] | 9563 |
| 23 | Macular Degeneration/di, dg, pc [Diagnosis, Diagnostic Imaging, Prevention & Control] | 4672 |
| 24 | Diabetic Retinopathy/di, dg, pc [Diagnosis, Diagnostic Imaging, Prevention & Control] | 9241 |
| 25 | Refractive Errors/di, dg, pc [Diagnosis, Diagnostic Imaging, Prevention & Control] | 2309 |
| 26 | 17 or 18 or 19 or 20 or 21 or 22 or 23 or 24 or 25 | 348878 |
| 27 | (UK or United kingdom or England or Wales or Scotland or Northern Ireland or Ireland or Australia or Canada or New Zealand or USA or United States or Austria or Finland or Germany or Malta or Netherlands or Norway or Spain or Sweden or Switzerland).tw | 1234304 |
| 28 | 8 and 16 and 26 and 27 | 3985 |
| 29 | (comment or editorial or letter).pt. or (comment or editorial or letter).ti,ab. | 2297837 |
| 30 | 28 NOT 29 | 3912 |
| 31 | (meta analysis or "review" or "systematic review").pt. or (meta analysis or "review" or "systematic review").ti,ab. | 4074667 |
| 32 | 30 NOT 31 | 3353 |
| 33 | Limit 32 to (English Language and humans and yr= 22009-Current" and English | **2059** |

**EMBASE Search Strategy**

Conducted 21.08.2023

EMBASE Search Strategy Conducted 21.08.2023
| # | Search Query | Results |
| --- | --- | --- |
| 1 | exp Prognosis/ | 914519 |
| 2 | Incidence/ | 567003 |
| 3 | Risk Assessment/ | 736472 |
| 4 | Decision Support Techniques/ | 23770 |
| 5 | (prognos* or prevention or progress* or diagnos* or detect* or prevalence or incidence or rate*).tw. | 13916058 |
| 6 | (risk* adj2 (assess* or factor*)).tw. | 1282303 |
| 7 | ((first or initial) adj (episode* or detection)).tw. | 33734 |
| 8 | 1 or 2 or 3 or 4 or 5 or 6 or 7 | 14825285 |
| 9 | exp Vision Screening/ | 43893 |
| 10 | exp Vision Tests/ | 43893 |
| 11 | exp Mass Screening/ | 308708 |
| 12 | ((eye* or sight or vision or visual) adj1 (test* or exam* or screen* or follow-up)).ti,ab. | 24543 |
| 13 | (asymptomatic adj2 (test* or exam* or screen*)).tw. | 6547 |
| 14 | (routine adj2 (test* or exam* or screen*)).tw. | 66221 |
| 15 | ((eye* or sight or vision or visual or screening) adj3 (frequenc* or interval* or recall*)).ti,ab. | 11477 |
| 16 | 9 or 10 or 11 or 12 or 13 or 14 or 15 | 443705 |
| 17 | Eye Diseases/ | 28376 |
| 18 | Vision, Ocular/ | 97000 |
| 19 | Visual Acuity/ | 135130 |
| 20 | (ametrop* or emmetrop* or glaucoma or diabetic retinopathy or refractive error or macular degeneration or cataract* or presbyop* or amblyop* or myop* or hyperop* or hypermetrop* or astigmat* or anisometrop* or vision impairment* or vision loss* or vis* acuity).tw. | 331096 |
| 21 | Cataract/di, dg, pc [Diagnosis, Diagnostic Imaging, Prevention & Control] | 4870 |
| 22 | Glaucoma/di, dg, pc [Diagnosis, Diagnostic Imaging, Prevention & Control] | 8600 |
| 23 | Macular Degeneration/di, dg, pc [Diagnosis, Diagnostic Imaging, Prevention & Control] | 619 |
| 24 | Diabetic Retinopathy/di, dg, pc [Diagnosis, Diagnostic Imaging, Prevention & Control] | 8050 |
| 25 | Refractive Errors/di, dg, pc [Diagnosis, Diagnostic Imaging, Prevention & Control] | 1373 |
| 26 | 17 or 18 or 19 or 20 or 21 or 22 or 23 or 24 or 25 | 486510 |
| 27 | (UK or United kingdom or England or Wales or Scotland or Northern Ireland or Ireland or Australia or Canada or New Zealand or USA or United States or Austria or Finland or Germany or Malta or Netherlands or Norway or Spain or Sweden or Switzerland).tw | 2164194 |
| 28 | 8 and 16 and 26 and 27 | 2477 |
| 29 | (comment or editorial or letter).pt. or (comment or editorial or letter).ti,ab. | 2215128 |
| 30 | 28 NOT 29 | 2433 |
| 31 | (meta analysis or "review" or "systematic review").pt. or (meta analysis or "review" or "systematic review").ti,ab. | 4598180 |
| 32 | 30 NOT 31 | 2086 |
| 33 | Limit 32 to (English Language and humans and yr= 22009-Current" and English | **1571** |

**CINAHL Search Strategy**

Conducted 21.08.2023

CINAHL Search Strategy Conducted 21.08.2023
| # | Search Query | Results |
| --- | --- | --- |
| 1 | (MM "Prognosis") | 1921 |
| 2 | (MM "Incidence") | 1712 |
| 3 | (MM "Risk Assessment") | 63998 |
| 4 | TX (prognos* or prevention or progress* or diagnos* or detect* or prevalence or incidence or rate*) | 3490646 |
| 5 | AB (risk* N2 (assess* or factor*)) | 239810 |
| 6 | TX ((first or initial) N1 (episode* or detection)) | 12704 |
| 7 | S1 OR S2 OR S3 OR S4 OR S5 OR S6 | 3564033 |
| 8 | (MM "Vision Screening") | 825 |
| 9 | (MH "Vision Tests+") | 7553 |
| 10 | TX ((eye* or sight or vision or visual) N1 (test* or exam* or screen* or follow-up)) | 19207 |
| 11 | TX (asymptomatic N2 (test* or exam* or screen*)) | 3757 |
| 12 | TX (routine N2 (test* or exam* or screen*)) | 33621 |
| 13 | AB ((eye* or sight or vision or visual or screening) N3 (frequenc* or interval* or recall*)) | 2663 |
| 14 | S8 OR S9 OR S10 OR S11 OR S12 OR S13 | 61314 |
| 15 | (MM "Eye Diseases") | 6614 |
| 16 | (MM "Vision, Subnormal") | 1133 |
| 17 | (MM "Visual Acuity") | 3893 |
| 18 | AB (ametrop* or emmetrop* or glaucoma or diabetic retinopathy or refractive error or macular degeneration or cataract* or presbyop* or amblyop* or myop* or hyperop* or hypermetrop* or astigmat* or anisometrop* or vision impairment* or vision loss* or vis* acuity) | 33125 |
| 19 | (MM "Cataract/DI/PC/PR") | 461 |
| 20 | (MM "Glaucoma/DI/PC/PR") | 1458 |
| 21 | (MM "Macular Degeneration/DI/PC/PR") | 1051 |
| 22 | (MM "Diabetic Retinopathy/DI/PC/PR") | 1433 |
| 23 | (MM "Refractive Errors/DI/PC/PR") | 290 |
| 24 | S15 OR S16 OR S17 OR S18 OR S19 OR S20 OR S21 OR S22 OR S23 | 43169 |
| 25 | TX (UK or United kingdom or England or Wales or Scotland or Northern Ireland or Ireland or Australia or Canada or New Zealand or USA or United States or Austria or Finland or Germany or Malta or Netherlands or Norway or Spain or Sweden or Switzerland) | 7754118 |
| 26 | S7 AND S14 AND S24 AND S25 | 4259 |
| 27 | PT ((comment or editorial or letter)) OR AB (comment or editorial or letter)) | 759333 |
| 28 | 26 NOT 27 | 4083 |
| 29 | PT ((meta analysis or "review" or "systematic review")) OR TI ((meta analysis or "review" or "systematic review")) OR AB ((meta analysis or "review" or "systematic review")) | 853902 |
| 30 | 28 NOT 29 | 3531 |
| 31 | 30 Limiters - Published Date: 20090101-20231231 | 2926 |
| 32 | 31 Limited to English Language | **2583** |

| Database | Results |
| --- | --- |
| Medline (Ovid) | 2059 |
| EMBASE (Ovid) | 1571 |
| CINAHL (EBSCO) | 2583 |
| TOTAL | **6213** |

## APPENDIX 2: Grey Literature resources

| Websites |
| --- |
| National Eye Institute \| National Eye Institute (nih.gov) |
| American Academy of Ophthalmology: Protecting Sight. Empowering Lives - American Academy of Ophthalmology (aao.org) |
| Help & Support For People Living With Glaucoma \| Glaucoma UK |
| RNIB \| Home |
| NICE \| The National Institute for Health and Care Excellence |
| Age-related macular degeneration - Macular Society |
| AMDf - Saving Sight Through Research and Education (macular.org) |
| Fight for Sight - Stopping sight loss through pioneering research |
| Cataracts & Other Eye Conditions: What You Should Know (beyondcataracts.uk) |
| Sightsavers \| Protecting sight and fighting for disability rights |
| Best Eye Charity in UK, Sight Loss Charity London \| Mission 4 Vision NGO in UK |
| General Optical Council (GOC) |
| British and Irish Orthoptic Society |
| Royal College of Ophthalmologists |
| College of Optometrists |
| European Council of Optometry and Optics (ECOO) |
| Association of Optometrists |
| International Agency for the Prevention of Blindness (IAPB) |
| Association of British Dispensing Opticians (ABDO) |
| Search terms |
| Vision test, visual test, test frequency, test interval, prognosis, incidence, progression, vision screening |
| Review, rapid review, systematic review, meta-analysis (for identification of secondary research only) |

## Notes

### Competing Interest Statement

The authors have declared no competing interest.

## 4. REFERENCES

Barsam A, Brennan N, Petrushkin H, et al. (2017). Case-control study of risk factors for acute corneal hydrops in keratoconus. British Journal of Ophthalmology. 101(4): 499. doi: 10.1136/bjophthalmol-2015-308251

Dinu M, Pagliai G, Casini A, et al. (2019). Food groups and risk of age-related macular degeneration: a systematic review with meta-analysis. European Journal of Nutrition. 58(5): 2123–43. doi: 10.1007/s00394-018-1771-5

Ekström C. (2012). Risk factors for incident open-angle glaucoma: a population-based 20-year follow-up study. Acta Ophthalmologica. 90(4): 316–21. doi: 10.1111/j.1755-3768.2010.01943.x

Ekström C, Hårleman K. (2023). Risk factors for incident open-angle glaucoma in clinical practice in Sweden: A population-based case-control study. Acta Ophthalmol. 101(5): 530–5. doi: 10.1111/aos.15644

Elmore A, Harris WS, Mu L, et al. (2022). Red blood cell fatty acids and age-related macular degeneration in postmenopausal women. European Journal of Nutrition. 61(3): 1585–94. doi: 10.1007/s00394-021-02746-2

Fernández-Montero A, Bes-Rastrollo M, Moreno-Montañés J, et al. (2017). Effect of pregnancy in myopia progression: the SUN cohort. Eye. 31(7): 1085–92. doi: 10.1038/eye.2017.24

Fight for Sight. (2020). Time to Focus. Fight for Sight. Available at: https://www.fightforsight.org.uk/media/3302/time-to-focus-report.pdf [Accessed 14 December 2023].

Gopinath B, Schneider J, Flood VM, et al. (2014). Association between diet quality with concurrent vision and hearing impairment in older adults. The journal of nutrition, health & aging. 18(3): 251–6. doi: 10.1007/s12603-013-0408-x

Guggenheim JA, Northstone K, McMahon G, et al. (2012). Time Outdoors and Physical Activity as Predictors of Incident Myopia in Childhood: A Prospective Cohort Study. Investigative Ophthalmology & Visual Science. 53(6): 2856–65. doi: 10.1167/iovs.11-9091

Hayden JA, van der Windt DA, Cartwright JL, et al. (2013). Assessing Bias in Studies of Prognostic Factors. Annals of Internal Medicine. 158(4): 280–6. doi: 10.7326/0003-4819-158-4-201302190-00009

Hex N, Bartlett C, Wright D, et al. (2012). Estimating the current and future costs of Type 1 and Type 2 diabetes in the UK, including direct health costs and indirect societal and productivity costs. Diabetic Medicine. 29(7): 855–62. doi: 10.1111/j.1464-5491.2012.03698.x

Hopf S, Heidt F, Korb CA, et al. (2022). Five-Year Cumulative Incidence and Progression of Myopic Maculopathy in a German Population. Ophthalmology. 129(5): 562–70. doi: 10.1016/j.ophtha.2021.12.014

Irving EL, Harris JD, Machan CM, et al. (2016). Value of Routine Eye Examinations in Asymptomatic Patients. Optometry and Vision Science. 93(7).

Kang JH, Loomis S, Wiggs JL, et al. (2012). Demographic and Geographic Features of Exfoliation Glaucoma in 2 United States-Based Prospective Cohorts. Ophthalmology. 119(1): 27–35. doi: 10.1016/j.ophtha.2011.06.018

Keel S, Lee PY, Foreman J, et al. (2017). Participant referral rate in the National Eye Health Survey (NEHS). PLOS ONE. 12(4): e0174867. doi: 10.1371/journal.pone.0174867

Kessel L, Erngaard D, Flesner P, et al. (2015). Cataract surgery and age-related macular degeneration. An evidence-based update. Acta Ophthalmologica. 93(7): 593–600. doi: 10.1111/aos.12665

Khachatryan N, Medeiros FA, Sharpsten L, et al. (2015). The African Descent and Glaucoma Evaluation Study (ADAGES): predictors of visual field damage in glaucoma suspects. Am J Ophthalmol. 159(4): 777–87. doi: 10.1016/j.ajo.2015.01.011

Marcus MW, Müskens RPHM, Ramdas WD, et al. (2012). Corticosteroids and Open-Angle Glaucoma in the Elderly. Drugs & Aging. 29(12): 963–70. doi: 10.1007/s40266-012-0029-9

NHS. (2021). Blindness and vision loss. NHS. Available at: https://www.nhs.uk/conditions/vision-loss/ [Accessed 14 December 2023].

Pasquale LR, Kang JH. (2011). Female reproductive factors and primary open-angle glaucoma in the Nurses’ Health Study. Eye. 25(5): 633–41. doi: 10.1038/eye.2011.34

Pezzullo L, Streatfeild J, Simkiss P, et al. (2018). The economic impact of sight loss and blindness in the UK adult population. BMC Health Services Research. 18(1): 63. doi: 10.1186/s12913-018-2836-0

Riley RD, Moons KGM, Snell KIE, et al. (2019). A guide to systematic review and meta-analysis of prognostic factor studies. BMJ. 364: k4597. doi: 10.1136/bmj.k4597

RNIB. (2021). Key statistics about sight loss 2021. Royal National Institute of the Blind. Available at: https://media.rnib.org.uk/documents/Key_stats_about_sight_loss_2021.pdf [Accessed 14 December 2023].

Robinson BE, Mairs K, Glenny C, et al. (2012). An Evidence-Based Guideline for the Frequency of Optometric Eye Examinations. Primary Health Care: Open Access. 2(4): 121. doi: 10.4172/2167-1079.1000121

Stahl A. (2020). The Diagnosis and Treatment of Age-Related Macular Degeneration. Deutsches Arzteblatt International. 117(29-30): 513–20. doi: 10.3238/arztebl.2020.0513

Stem MS, Talwar N, Comer GM, et al. (2013). A longitudinal analysis of risk factors associated with central retinal vein occlusion. Ophthalmology. 120(2): 362–70. doi: 10.1016/j.ophtha.2012.07.080

Stingl JV, Ban SA, Nagler M, et al. (2023). Five-year change in refractive error and its risk factors: results from the Gutenberg Health Study. Br J Ophthalmol. 107(1): 140–6. doi: 10.1136/bjophthalmol-2021-318828

Whiting P, Savović J, Higgins JPT, et al. (2016). ROBIS: A new tool to assess risk of bias in systematic reviews was developed. Journal of Clinical Epidemiology. 69: 225–34. doi: 10.1016/j.jclinepi.2015.06.005

Wright DM, O’Reilly D, Azuara-Blanco A, et al. (2020). Delayed attendance at routine eye examinations is associated with increased probability of general practitioner referral: a record linkage study in Northern Ireland. Ophthalmic and Physiological Optics. 40(3): 365–75. doi: 10.1111/opo.12685

